# How information source and scale shape pandemic behavior change

**DOI:** 10.64898/2026.03.09.26347630

**Authors:** Casey M Woika, Juliana C Taube, Vittoria Colizza, Shweta Bansal

**Affiliations:** Department of Biology, Georgetown University, Washington, DC, USA; INSERM, Paris, France

**Author notes:** Shared first authorship (Casey M Woika, Juliana C Taube).

## Abstract

**Background:** The COVID-19 pandemic highlighted the importance of incorporating human behavior into infectious disease models. Doing so requires predicting how individuals will respond to outbreaks and policies. Key questions that limit the integration of dynamic behavior into disease models are: Is behavior change driven by policy, objective disease information, or subjective risk perception? Can disease data approximate risk perception? Is mitigation behavior influenced by information at broad or fine spatial scales?

**Methods:** In this ecological study, we leverage US survey data (4.3 million responses from September 2020 through January 2021) on social distancing and concern about infection, alongside data on COVID-19 cases and mitigation policies. Using a county-level spatiotemporal regression model, we evaluate whether social distancing is associated with policy, social information, or epidemiological information at broad or fine spatial scales. We use transmission models to demonstrate the epidemic consequences of different behavioral drivers.

**Results:** We find that epidemiological and social information are predictive of mitigation behavior: a one standard deviation increase in epidemiological or social information led to a decrease in non-household contacts of 0.10 or 0.47 people per day, respectively. State-level variables are more predictive of social distancing than county-level variables, especially for policies. However, conditions in socially adjacent counties are better predictors of behavior than those in spatially adjacent counties. In our transmission model, epidemiologically and socially-informed contact patterns yield similar epidemic dynamics, though broader spatial scales of information explain more behavioral variation.

**Conclusions:** The US population modified their behavior during the COVID-19 pandemic in response to case incidence and policy at broader spatial scales, potentially reflecting the lack of localized surveillance or mandates. Modeling assumptions of behavioral responses to disease incidence at larger spatial scales are reasonable.

## Introduction

Human behavior and infectious disease dynamics are inextricably linked through a feedback loop [1]. Behavior determines who comes into contact with whom and the ensuing opportunities for direct infectious disease transmission. This side of the feedback loop and its impacts on epidemiological dynamics has been more thoroughly characterized, for example through the incorporation of social contact heterogeneity using contact networks [2] and contact matrices [3]. Less empirically studied is the impact of disease on behavior. A myriad of hypotheses have been developed by theoretical models, but few have been validated [4, 5, 6]. While behavior change by infected individuals, for example via sickness behaviors or isolation [7, 8, 9], is relevant for transmission, we focus on behavior change in the susceptible population. Susceptible individuals often compose a majority of the population, meaning that their behavior can have a substantial impact on epidemic dynamics, whether driven by policy, fear, or exposure risk. Understanding the drivers of susceptible behavior change and parameterizing this feedback loop is critical for future modeling efforts. After all, behavioral changes are the first line of defense against an epidemic, as exemplified by the COVID-19 pandemic [10].

There is ample evidence that susceptible individuals alter their behavior during epidemics. Susceptible behavior change can result from top-down influences, like policies and recommendations, that affect social contact behavior. For example, influenza school closures reduce contact with classmates [11, 12] and COVID-19 stay-at-home orders resulted in a 12-15% increase in time at home, and a 25% reduction in overall mobility [10, 13, 14]. Behavioral changes can also be individually-motivated, creating a bottom-up response to information and awareness of infectious disease [15]. For example, in the US, 16-25% of people avoided public spaces and 20% reduced non-household social contacts during the 2009 H1N1 influenza pandemic [16], and time spent at home began to increase prior to COVID-19 social distancing policies, indicating voluntary behavioral change [13]. In order to be able to predict the magnitude and timing of these behavior changes, we need to understand what specifically is driving them.

There is a strong literature of theoretical behavior-disease models that evaluate the interaction of behavioral and infection dynamics [6, 4, 5, 17]. Individually-motivated behavior has been incorporated into such studies in a number of ways. The earliest coupled disease-behavior models assumed that behavior was prevalence-elastic, that is, that behavior changes as a function of local or global disease dynamics [18]. There is some evidence to support this assumption: Ahituv et al. found that condom usage increased among men upon the onset of the AIDS epidemic in the United States [19] and Philipson showed that children received their first MMR vaccine earlier in states with higher measles incidence [20]. Whether these findings generalize to other pathogens and behaviors, especially in the current information environment, remains unclear. Other models have incorporated behavior change through a “fear” class [21] in which individuals change their behavior in response to information, or through a game-theoretical tradeoff between risk and social contact [22]. However, social science research suggests that individual risk perception may diverge from reality, shaped by the information sources people consult and their personal experiences. Indeed, humans are known to misestimate their risk of infection [23, 24]. Moreover, many models assume that individuals respond to risk homogeneously, spurring recent efforts to examine the epidemiological consequences of varied risk tolerance across a population [25]. The most common feature across these models is their lack of validation, with a 2016 study estimating that about 20% of behavior-disease modeling studies use any empirical evidence [5]. Therefore, it is critical that we evaluate these common modeling assumptions using real-world data.

One of the most frequent assumptions in these models pertains to whether individuals respond to local or global information about disease [21, 26]. Evaluating this assumption is particularly relevant in the United States, a demographically heterogeneous, geographically large, and politically diverse country that did not implement national-scale policies during the COVID-19 pandemic. Instead, US public health authority and surveillance infrastructure is fragmented and distributed across states and counties [27]. In consequence, communities throughout the country were subject to different policies, had access to different information, and were exposed to different disease risks during the COVID-19 pandemic. Policies were enacted at both the state and county level, and data on cases, hospitalizations, deaths, and vaccinations had to be manually compiled from different sources [27]. Risk was also communicated at different spatial scales: some were informed by non-local social media or traditional national news broadcasts, while others relied on more community-specific sources such as county or state websites, neighbors, and trusted leaders. Information transmission can occur in several ways: homophily, where individuals that are similar to one another live geographically close to one another, and thus adjacent communities impact one another; person-to-person information transmission, where individuals who come into contact with one another are more likely to influence one another; and social media information transmission, where people can impact others perceptions and policies over social media platforms. The pathways through which information is disseminated likely shape the risk that individuals perceive and their response to it. The uncoordinated landscape of policy and information in the US provides a rich setting in which to interrogate these possible mechanisms of information spread and improve our understanding of the drivers of behavior change during an epidemic.

Other researchers have considered the impacts of policies and perceptions on social behavior in the US. For instance, Yan et al. found that both county and state case incidence, as well as policy-induced behavior change, contributed to time spent at home at the onset of the COVID-19 pandemic in the US [13]. Urmi et al. found that national COVID-19 mortality explained periods of increased risk aversion throughout the pandemic, but state mortality was not always predictive of state risk-aversion at coarse temporal scales [28]. During the 2009 H1N1 epidemic, Bayham et al. found that individuals spent more time at home as cases increased, but that time spent at home was not associated with Google search trends [29]. Despite this progress, our understanding of the relative contribution of epidemiological information, social information, and policy on epidemic behavior change remains limited.

In this study, we hypothesize that behavior change is driven proximately by perceived risk, which is informed both by epidemiological information (direct measures, like case counts) and by social information (norms, beliefs, and cues from one’s social network), and that policy also acts on behavior through its influence on this perceived risk. Our central question is whether behavior change during epidemics is driven more by epidemiological information or social information, and what role policy plays in shaping this response. We first implement a spatiotemporal regression model of counties in the US to 1) compare the effects of policies versus perceptions on mitigation behavior during the COVID-19 pandemic, 2) identify the most predictive spatial scale of epidemic information, and 3) evaluate whether counties influence one another through physical proximity, physical mobility or social connectivity. We find that epidemiological and social information are both predictive of behavior, and that mesoscale information predicts behavior more than microscale. We then translate our findings into an empirically-parameterized behavior change epidemiological model, where we compare how disease dynamics are impacted by social compared to epidemiological information, and how epidemiological dynamics change when state versus county-level factors drive local behavior. Consistent with our first analysis, we find that epidemiological information is interchangeable with social information in coupled disease-behavior dynamics models and that models using meso-or macroscale information are able to better explain variation in behavior change compared to models using microscale information. Our results have implications for identifying the most effective scale for health policy, public health communication, disease surveillance, and behavioral data collection, as well as for determining which assumptions about human behavior are reasonable for disease modelers to make.

## Methods

To compare the association of COVID-19 policies, social information, and disease trends with protective health behaviors, we use a multivariate regression approach with social distancing as the response. Based on these findings, we demonstrate the impacts of various behavior change assumptions on model predictions using an epidemiological model.

### Data

We take advantage of a number of different data sources to quantify behavior, risk, and factors that could affect the relationship between them. All data are included at the county level spatially and on the weekly scale temporally for the time period from September 2020 through January 2021.

To quantify epidemiological information informing perceived risk, we incorporated daily data on COVID-19 cases and deaths reported by the New York Times [30] and aggregated to the weekly level. The case data are likely subject to underreporting, and we examine this possibility and its effects on our results in a sensitivity analysis [31] (Figures S13, S14).

To estimate social distancing (behavior change), we used survey data collected in the COVID-19 Trends and Impact Survey (CTIS) designed by the Delphi Group at Carnegie Mellon and administered via Facebook [32]. Among other questions related to their behavior, beliefs, and choices during the pandemic, respondents were asked to report the number of non-household contacts they had in the last 24 hours. Contacts were defined as “a conversation lasting more than 5 minutes with a person who is closer than 6 feet [1.8 meters] away from you, or physical contact like hand-shaking, hugging, or kissing” and can be interpreted as a measure of the degree to which people were social distancing. We used daily data from over four million respondents reweighted to match population age and gender composition; these data were then aggregated to the county-week level (3,113 counties) and temporally smoothed with generalized additive models to address noise and low sample sizes, as described in [33]. We assess the robustness of our findings to this source of social distancing data by running sensitivity analyses using mobility data (Figures S1, S7, S8), as described below. This study was reviewed by the Institutional Review Board at Georgetown University and was determined not to be human subjects research.

To measure social information informing perceived risk, CTIS respondents were additionally asked to rate their concern about “the possibility that [they] or someone in [their] immediate family might become seriously ill from COVID-19 (coronavirus disease)” on a four-point scale. We quantified the prevalence of social information prompting high perceived risk for COVID-19 by estimating the proportion of individuals who were “somewhat worried” or “very worried” in each county-week; we call this *proportion worried* (Figure S38). This variable is included to measure the subjective perception of risk, which captures the impact of social norms and beliefs as well as influence from social networks. When aggregating to the county-week scale, we used the same weights as calculated for the social distancing responses to correct for age and gender representation.

To evaluate the effect of policy, we utilized data on COVID-19 State and County Policy orders from [34]. This is a curated dataset that standardizes state and county orders from a number of sources. For both states and counties, we classified policies as either present (1) or not present (0) for a state or county in a given week (Figures S25, S26, S27, S28, S29, S30, S31, S32, S33, S34, S35, S36, S37). The considered policies were those that restricted food and drink businesses, houses of worship, outdoor and recreational activities, non-essential businesses, daycare or childcare (combined from separate categories reported in the dataset for simplicity), entertainment businesses, and manufacturing businesses. To further characterize policies, we also included the composite state government response index from the Oxford COVID-19 Government Response Tracker [35]. This index captures the extent of policy limitations on human behavior, including school, work, and transit closures, stay-at-home requirements, travel restrictions, contact tracing, testing, masking and vaccination policies, and government economic support at the state level.

We controlled for other factors that may have influenced individuals’ social distancing behavior but were not related to COVID-19 perceptions and policies. First, we included data on weather, aggregated to the county-week. We considered the measures minimum humidity, maximum humidity, solar radiation, minimum temperature, maximum temperature, wind speed, and precipitation collected from [36]. Similarly, because the ability to interact outdoors may have influenced the number of interactions that people had, we considered the propensity for interactions to occur indoors versus outdoors during the winter months using the metric defined in [37]. Susswein et al. calculate the ratio of visits to indoor locations (e.g., grocery stores, schools, doctors’ offices) versus outdoor locations (e.g., amusement parks, cemeteries) at the county-level using mobile-phone location data from Safegraph [38, 37]. We additionally included the static metrics of population density, measured as number of people in a county (from [39]) divided by the square miles in the county (from [40]), and socioeconomic factors. The socioeconomic data are from County Health Rankings [41]; we considered income (measured as median income), education (measured as proportion completed high school), and race (proportion of the population that is Black, White, Hispanic, Asian, and American Indian/Alaskan Native). Finally, we included an indicator of whether or not there was a bank holiday each week, as a holiday may increase mobility and social contacts as people travel and participate in leisure activities, or decrease mobility and social contacts as people do not attend school or work.

### Spatial scales of information

We aimed to identify which spatial scales of information are most predictive of social distancing behavior; therefore, we incorporated data on perceptions, cases, and policies at different scales to estimate their relative effects. We consider two US administrative levels (state and county) and three intermediate spatial scales, to account for how counties impact one another. We aggregate disease, worry, and policy covariates across linked counties included in each intermediate spatial group by taking a weighted mean of each variable across the linked counties. The first intermediate scale is *neighboring counties*; here, we assumed that through homophily, similar populations live near one another, and thus neighboring counties might impact one another equally. The second scale that we consider is *commuting counties*. We assumed that counties may have influenced one another through person-to-person physical social interaction; we use county-level commuting data derived from Safegraph as a proxy for these interactions [42]. Safegraph records mobility data from approximately 10% of mobile devices in the US, and the data has been demonstrated to be relatively unbiased by income, education, race, and geography [43]. During aggregation, commuting counties are weighted by their commuting volume to a destination county. We dropped edges with weight less than 0.1% of the population (e.g., 27 individuals at the median US county population size) because ties this weak are the least reliable given how sparse the underlying observations are, and add computational cost without improving explanatory power. The third scale that we consider is *socially connected counties* where we assume that counties may influence each other through social media. Here, we used Facebook’s county-level social connectedness index (SCI) [44], which quantifies how socially connected US counties are to one another through Facebook friends. During aggregation, socially connected counties are weighted by this index; counties with weights less than 20 were dropped based on visual inspection of the weight distribution. We note that no data were included at the national level, as none of the considered policies were implemented by the national government and adding other national-level variables introduced collinearity.

### Spatiotemporal regression model

To investigate the relative association of different factors with social distancing throughout the COVID-19 pandemic, we used R-INLA to create a spatiotemporal regression model [45, 46]. R-INLA is a package for implementing Bayesian inference through integrated nested Laplace approximation (INLA) models. INLA has demonstrated computational efficiency for latent Gaussian models, produced similar estimates for fixed parameters as established implementations of Markov Chain Monte Carlo (MCMC) methods for Bayesian inference, and been applied to disease mapping and spatial ecology questions [47].

Our regression model predicts county-week mean number of non-household contacts, a measure of social distancing, using the covariates listed in Table 1. COVID-19 deaths were not included in the main model due to collinearity, but we considered them in sensitivity analyses (Figures S9, S10). A subset of weather and race/ethnicity variables were included to avoid multicollinearity. Childcare policy data was only included at the county level as there were no state-level childcare policy changes in our dataset during the study period. The variance inflation factor (VIF) was calculated to check for problematic multicollinearity, and all included covariates fell below a threshold VIF value of 10 (Table S1). County-weeks missing any of the covariates were dropped, leaving 2,187 counties with 22 weeks of data for the main model. Alaska and the District of Columbia are excluded from the analysis.

**Table 1:** Variables included in the spatiotemporal regression model.

| Variable | Spatial scale | Category |
| --- | --- | --- |
| COVID-19 case incidence | County, state | Epidemiological information |
| Prop. worried | County, state | Social information |
| Restaurant/bar closure | County, state | Policy |
| Worship restrictions | County, state | Policy |
| Outdoor recreation restrictions | County, state | Policy |
| Non-essential business restrictions | County, state | Policy |
| Entertainment restrictions | County, state | Policy |
| Manufacturing restrictions | County, state | Policy |
| Childcare restrictions | County | Policy |
| Government response stringency index | State | Policy |
| Min. temperature | County | Control |
| Max. windspeed | County | Control |
| Precipitation | County | Control |
| Population density | County | Control |
| Prop. graduated high school | County | Control |
| Median income | County | Control |
| Prop. Black | County | Control |
| Prop. Hispanic | County | Control |
| Prop. essential workers | County | Control |
| Indoor activity index | County, state | Control |
| Holiday week | National | Control |

All temporally-varying covariates were z-normalized at their respective spatial scale (i.e., county variables with county mean and standard deviation, state variables with state mean and standard deviation), while temporally stable variables were z-normalized with the national mean and standard deviation. We chose this normalization approach to limit collinearity, allow comparison across predictors, and center values at spatial unit means. We considered temporal lags and rolling averages of case incidence and proportion worried and used the values that yielded the lowest DIC (see Figures S11, S12 for sensitivity to these choices). Uninformative priors were included for all predictor variables. Independent and identically distributed random effects were included for each state to account for baseline differences in mitigation behaviors. We considered autoregressive order 1 (AR1), AR2, random walk order 1 (RW1), and RW2 temporal autocorrelation models, and selected the RW1 model since it had the lowest DIC and produced a good model fit. We did not include spatial autocorrelation as it did not improve model fit. The selected model fits well and does not leave spatially-stratified residuals (Figure S2).

We repeated the same modeling approach for each intermediate spatial scale. We predict focal county social distancing based on both focal and linked county epidemiological information, social information, and policy covariates, as well as focal county control variables. Due to collinearity among linked county variables, we ran three separate models across the intermediate scales – one model each for neighboring, commuting, and socially connected counties (Tables S2, S3, S4). These models cover 2,160 counties over 22 weeks and fit the data well (Figures S4, S5, S6).

### Social distancing sensitivity analysis

In the main model we use contacts reported via surveys as a measure of social distancing. These data are subject to recall, social desirability, and response biases that we could not entirely control for. To assess the robustness of our findings, we ran the same model using mobile-phone location data from Safegraph [42] as the response which provides a more objective measure of social activity due to its automatic collection through mobile app usage. From Safegraph, we used the rate of visitors to 5.5 million points of interest (e.g., schools, hospitals, parks, grocery stores, restaurants) normalized by the maximum number of visits to each location in 2019. This model does not fit as well as the main model (Figures S7, S8), but most qualitative findings agree with the model using subjectively-reported social contact data.

### Epidemiological model

To assess the validity of common disease modeling assumptions about local versus global information sources and prevalence-elastic behavior change, we developed an empirically-parameterized epidemiological model. Our parameterizations capture common and available assumptions of behavior-disease models where risk is measured objectively, assessed “globally”, and responded to uniformly.

These models are data-driven but parsimonious representations of behavioral and epidemiological dynamics and aim only to consider the role of risk perception and spatial scales on epidemiological dynamics; they are not intended to be predictive in nature. To implement this model, we used simple regression models to predict time-varying contact rates as a function of data on epidemiological and social information. We then incorporated these behavioral predictions into a compartmental model, composed of two susceptible populations differentiated by their contact rates (worried versus unworried).

We compare epidemiological information (cases) versus social information (worry) as predictors of social distancing behavior at the county, state, and national scale. Using simple linear regressions with each of these covariates (cases and worry) and complete pooling, we predict social distancing for two groups, *worried* and *unworried* individuals, as determined by their survey response. We run twelve different regressions: three spatial scales *×* two worry groups *×* two types of information.

Each regression model is structured as follows:

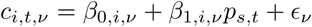

where

- *c_i,t,ν_*: mean contact in county *i*, week *t*, and worry group *ν* (worried or unworried)
- *p_s,t_*: predictor at spatial scale *s* (county, state, or national) during week *t*. The predictor is either the 3-week right-aligned rolling average of case incidence or proportion worried.
- *ɛ_i,t,ν_*: error, where *ɛ ∼ N* (0*, σ*^2^)

Contact predictions from these simple regressions are incorporated into discrete-time Susceptible-Infected-Recovered (SIR) models for each county with available data. The susceptible class is divided into two populations, worried and unworried, similar to [48, 21]; the distribution of individuals into these classes is determined by the proportion of the survey respondents in that county that report being worried for the first week of the simulation in September 2020. We use parameters that are similar to COVID-19, but could be generalized to other respiratory-transmitted infectious diseases: we assume an *R*_0_ of 2 [49], an infectious period (1*/γ*) of 10 days [50], and we assume no re-infection. Transmissibility is calculated as 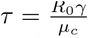 where *µ_c_* represents mean contact over all counties across the study period. The force of infection varies between the two susceptible populations due to different contact rates. The number of newly infected individuals at each time step *t* is determined by:

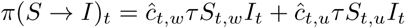

where

- *ĉ_t,w_* and *ĉ_t,u_*: predicted contact from our simple linear regressions for worried (*w*) or unworried (*u*) individuals at time *t*
- *S_t,w_* and *S_t,u_*: the number of susceptible individuals in worried (*w*) and unworried (*u*) classes at time *t*
- *I_t_*: the number of infected individuals at time *t*

We assume that infected individuals of both worry statuses have the same contact patterns and infectivity. The model was initialized with COVID-19 incidence from the previous two weeks (8/24/2020 - 9/6/2020).

We compare the predicted contact rates based on epidemiological and social information to observed patterns. Then, we compare the predicted case incidence time series arising from each set of contact predictions and observed patterns. In particular, we examine how epidemic peak time and peak size derived from contact patterns predicted using epidemiological or social information differ from those predicted based on observed contact patterns.

## Results

We investigate whether epidemiological information, social information, or policies were associated with social distancing behavior during the COVID-19 pandemic, and we evaluate the spatial scale that most predicts individual behavior using spatiotemporal regression. In addition to the administrative scales of county and state, we include three intermediate spatial scales summarizing potential between-county influence based on geographic proximity, population movement, and social media connectedness. Finally, using compartmental models, we demonstrate where traditional coupled disease-behavior modeling assumptions hold up and where they can go wrong.

We use social distancing data across 2,187 counties from 4.3 million responses to the US COVID-19 Trends and Impact Survey beginning in September 2020 through January 2021. This period encompasses a full wave of COVID-19 infections across the US, during which there was increased social distancing, concern about infection, and policies restricting movement and gatherings. National concern about becoming seriously ill from COVID peaked at 72% of responses, while a maximum of 49 counties and 26 states had any of the seven considered policies in place during each study week.

### Behavior change is predicted by both epidemiological and social information

Across epidemiological information, social information, and policy variables, state-level social information is most strongly associated with increased social distancing (Figure 1). State-level case incidence and non-essential business restrictions are the next strongest predictors of increased social distancing (fewer contacts). The strength and direction of the association between social distancing and policy varies by policy category: state-level manufacturing and outdoor recreation policies are associated with more social distancing, while state-level worship and entertainment policies are associated with less social distancing. Increased overall stringency in state-level restrictions is also associated with increased social distancing. No county-level information or policy variables are significantly associated with social distancing. These results suggest that social information, while one of the most difficult variables to measure, is the most valuable indicator of how people will alter their behavior, and it is more predictive at larger spatial scales. However, epidemiological information alone could be a sufficient predictor of behavior change – we explore this possibility below.

**Figure 1:**
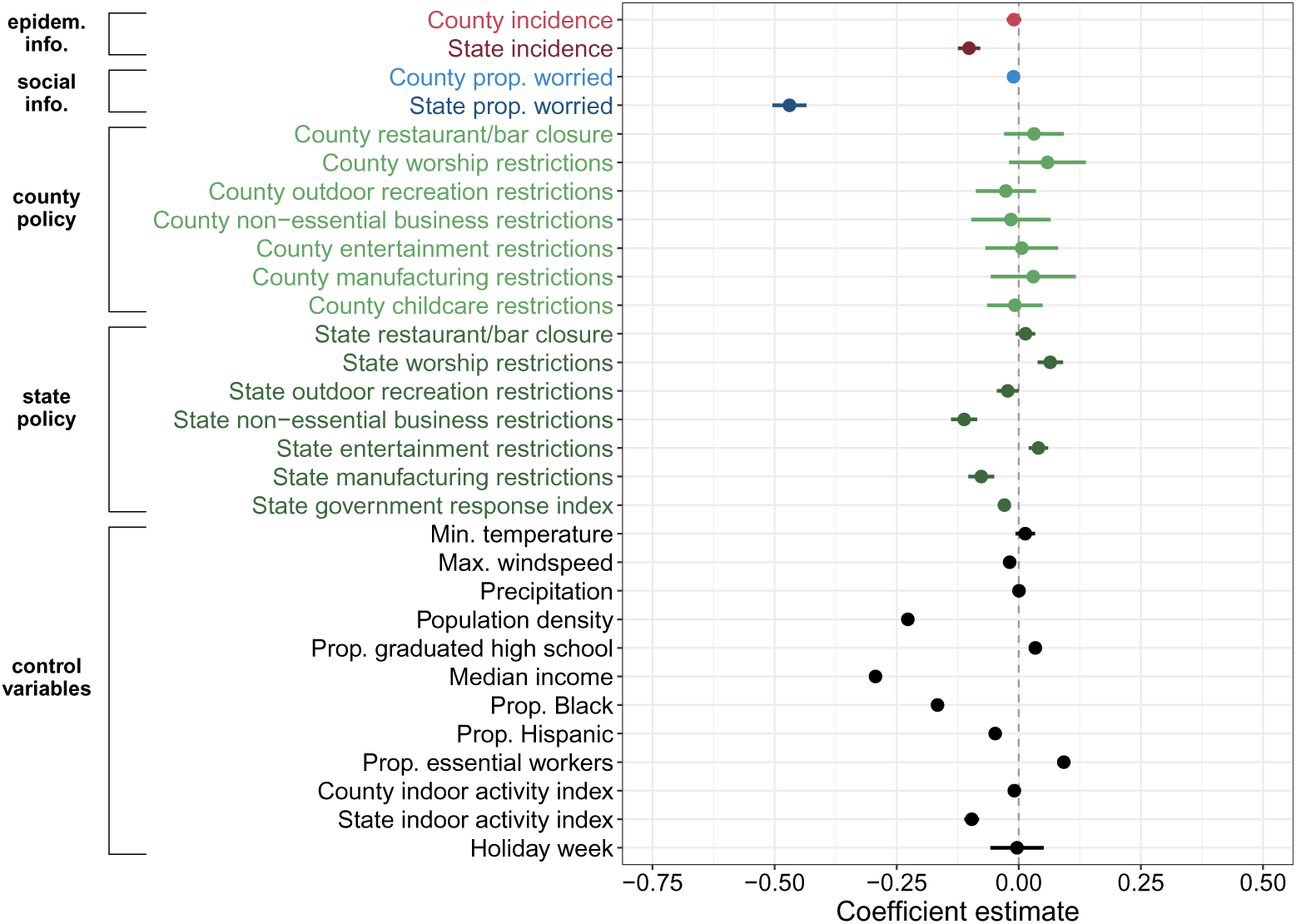
While epidemiological and social information are both strongly predictive of social distancing, state level variables are more predictive than county variables. Coefficient estimates and 95% credible intervals of the spatiotemporal regression predicting county-week mean number of non-household contacts per day, as a measure of social distancing. Positive coefficients indicate higher contact rates and lower social distancing. This analysis is performed for September 2020 through January 2021. n = 48,114 county-weeks.

These findings are robust to aggregation of epidemiological and social information over different time periods (Figures S11, S12), cross-validation (Figures S15, S16), and using incident deaths as a predictor of social distancing instead of cases (Figures S9, S10). However, increased state incidence is associated with decreased social distancing when mobility is used as the response instead of non-household contacts (Figures S7, S8). County epidemiological information is more predictive of behavior than county social information when state-level variables are excluded from the model (Figures S23, S24). When we correct for underreporting of cases, we observe similar trends albeit with higher uncertainty in our coefficient estimates to the point where corrected state case incidence is no longer associated with social distancing (Figures S13, S14).

### Behavior change is associated with physical and social interactions

We compared how policies, perceptions, and epidemiological information at county, state, and intermediate scales are associated with social distancing to identify the most predictive spatial scale. We find that state-level variables are more predictive of social distancing than county-level variables across covariates (Figure 1). Due to collinearity, we could not include national epidemiological or social information in the model, nor could we compare state and intermediate scale covariates within the same model. However, our results are robust when we restrict the analysis to larger sample size counties (Figures S18, S19, S20) and impose stricter collinearity constraints (Figures S21, S22).

At intermediate spatial scales, both linked county case incidence and proportion worried are significantly associated with the degree of social distancing in the focal county (Figure 2). Notably, linked county epidemiological and social information are more associated with behavior than focal county information, though the effects of policies in linked versus focal counties are neither consistently different, nor meaningfully predictive (Figure S3). In neighboring counties, social information is less predictive than epidemiological information, whereas social information is more predictive than cases in socially connected counties. This relationship holds under cross-validation (Figure S17). The two factors are nearly equally predictive in commuting counties. These results suggest that counties can influence each other through several pathways, including the spread of disease risk perceptions via social media interactions.

**Figure 2:**
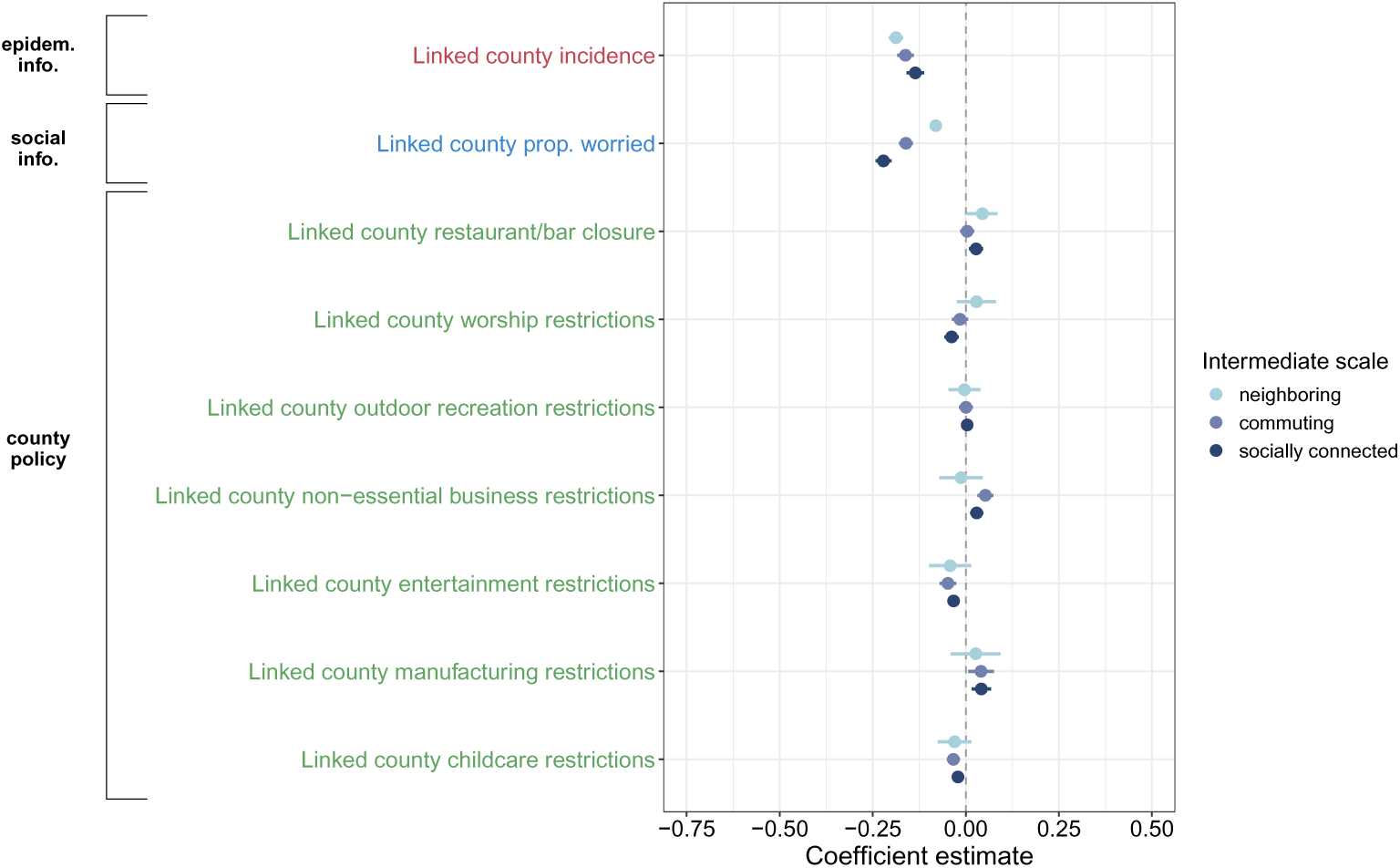
Interactions, whether physical or social, predict behavior change more than physical proximity. Coefficient estimates and 95% credible intervals of spatiotemporal regressions predicting county-week mean number of contacts per day as a measure of social distancing using intermediate spatial scales. This analysis is performed for September 2020 through January 2021. Neighboring counties share a border with the focal county. Commuting counties are connected to the focal county via commuting flows. Socially connected counties are connected to the focal county through friendships on Facebook. Positive coefficients indicate higher contact rates and lower social distancing. n = 41,040 county-weeks.

### Epidemiological information and social information are interchangeable in behavior-disease dynamics models

Although we find that social information is more predictive of social distancing than epidemiological information, many behavior-disease models assume that individuals respond to local epidemiological conditions. We test these common assumptions by fitting univariate regression models that predict social distancing behavior from either case incidence or survey-reported worry at different spatial scales, then assessing how these differing inputs affect epidemic dynamics in a transmission model. Social information and epidemiological information yield broadly consistent social distancing predictions across counties (Figures 3, S40, S43), with both deviating from observed patterns at the same time and in the same direction. This similarity translates to nearly indistinguishable epidemic dynamics in the transmission model (Figures 3, S41, S44), with the main exception being counties in the states of Wyoming, Idaho, Utah, and Nevada, where epidemiological information underpredicted peak size, while social information overpredicted it. Both epidemiologically and socially-informed behavior change models tend to more accurately predict epidemic peak time and size than models without behavior change (Figure S42). In general, these results support the continued use of epidemiologically-informed risk in coupled disease-behavior models and suggest that resources may be better spent on disease surveillance rather than on collecting social risk perception data.

**Figure 3:**
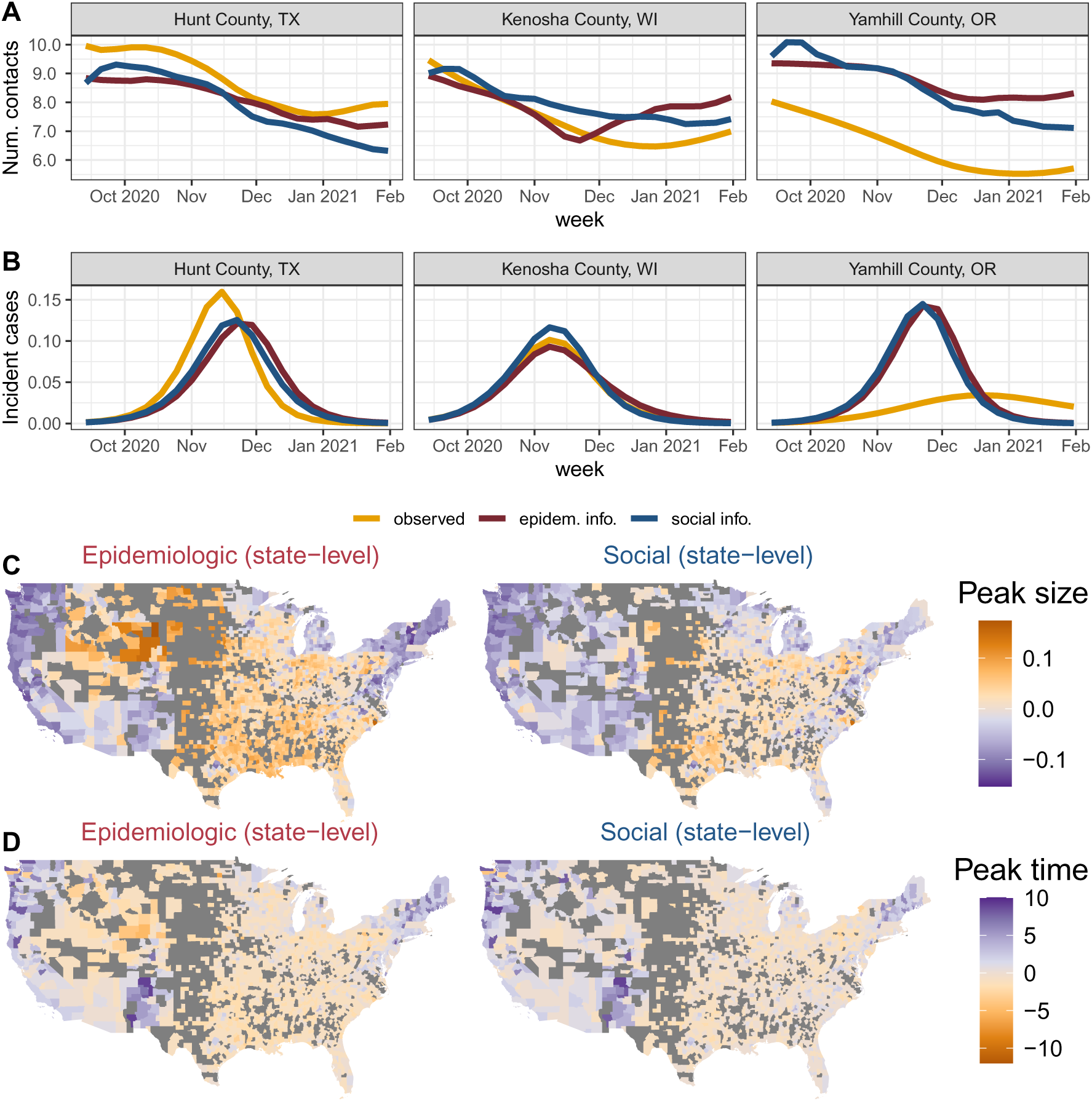
Social distancing driven by epidemiological versus social information produces similar disease outcomes across counties. (A) Three case study counties demonstrating the range in predicted social distancing using case incidence (objective) versus proportion worried (subjective) at the state level compared to the observed social distancing for winter 2020-21. Higher contact rates correspond to less social distancing. All three counties are considered metropolitan areas in terms of population density. (B) The repercussions of social distancing predictions for disease projections in the same three case study counties. We use deterministic models so there are no confidence intervals. (C, D) Epidemic repercussions across all counties for social distancing predictions based on state incidence or proportion worried. Peak size is the maximum proportion of the population infected at any one time (C) and peak time is the week in which the peak size occurs (D). For peak size, positive values (orange) indicate that predicted epidemics had a smaller peak incidence than observed, while negative values (purple) denote that predicted epidemics had a larger peak incidence than observed. For peak time, positive values (purple) denote predicted epidemics peaked earlier than observed and negative values (orange) indicate that predicted epidemics peaked later than observed.

Overall, we find that state social information and national case incidence are able to best explain the variation in social distancing patterns (adjusted-R^2^s above 0.4 for unworried individuals and above 0.3 for worried), though the residuals vary spatially. Thus, in general, higher scales of risk information are better at predicting behavior than local variables, which suggests that people are deriving their information from more “global” sources, rather than local community conditions. This result is troubling from a control perspective, as it means people may be taking precautions at suboptimal times relative to the disease incidence in their area. On the other hand, it could also be advantageous if people anticipate the need for social distancing by drawing on information from non-local contexts experiencing more critical conditions.

## Discussion

Human behavior is a critical component of disease transmission and remains challenging to predict. We retrospectively analyze the predictors of social distancing behavior during the COVID-19 pandemic and discuss the implications for future disease modeling efforts. We consider three possible categories of behavioral drivers (epidemiological information, social information, and policies) across five spatial scales (county, state, neighboring counties, commuting counties, and socially connected counties) and assess which is most predictive of social distancing behavior. We find that social information is a slightly stronger predictor of behavior than epidemiological information, and that state-enacted COVID-19 policies were more associated with behavior change than county-level policies. We additionally utilize an empirically-driven epidemiological model to evaluate how perception-driven behavior changes alter epidemic dynamics, compared to the typical behavior-disease model that relies on rational decision-making. We find that epidemiological information is a good substitute for social information, and that the spatial scale of predictor used to project behavior does not notably impact the results, despite epidemiological information and county-level variables being weaker predictors in the multivariate spatiotemporal regression model. These findings can facilitate the much-needed improvement of future disease models and interventions in the US.

Our finding that broader scale perceptions, data, and policies are more linked to human behavior has several consequences for public health. This result suggests that people may be getting pandemic-related information from less-localized sources, such as social media, television, and large news outlets, as opposed to local newspapers and interactions. Policy was primarily enacted at the state-level in the US, and state-level policies may have been better enforced and communicated than county-level policies. Furthermore, in some cases, county-level policies may have been in opposition with state policies or weakly enforced due to political differences at the county versus state level and state government preemption (legislation that prevents counties from implementing their own stricter policies) [51, 52]. Heterogeneity in county-level policies could have introduced confusion and allowed people to travel to a nearby county with different policies, resulting in different rates of social contact and disease spread. Therefore, policies might be most impactful, in terms of both recognition and implementation, if maintained by the state, or, at minimum, coordinated between adjacent counties to prevent policy-escape between counties [53]. Local risk-motivated behavior change can facilitate more spatiotemporally targeted transmission reduction than global risk-motivated behavior change. Therefore, investing in local media coverage of risk and enacting more policy agility could be effective. Nevertheless, mesoscale information appears to be associated with shifts in behavior in our data, suggesting that disease models that use mesoscale rather than microscale perception may better capture the link between disease and behavior.

Social information was the most important predictor of changes in social distancing across spatial scales. This finding is supported by other studies on risk perception and health behaviors both for childhood vaccination [54] and for COVID-19 [55]. While there may be an interaction between policies and risk perception [13], we aimed to account for this relationship by including both variables in our model. Still, it remains possible that other unmeasured policies, recommendations, or declarations could have influenced risk perception, meaning this perception may not be entirely self-derived. This distinction matters because data-driven risk results in behavior change in areas where transmission is occurring and is thus more effective, whereas socially-informed risk can result in behavioral changes that are disconnected from measured disease dynamics, making these behavioral changes less impactful. Thus, the mechanism that is most predictive of behavior change (social information) is actually the less effective strategy. Interventions that bridge the gap between perceived and true risk, like interventions that increase awareness about local disease conditions, could be a productive public health step [24]. Our findings align with those from other countries; for example, in France, both epidemiological and social context are predictive of mitigation behavior [56], and in Germany, broader scale policies were more associated with behavior change than local ones [57]. Because risk perception data are difficult to collect, we explored how using epidemiological information, i.e., recent case incidence, would alter predicted behavioral and epidemic dynamics. We find that using epidemiological information to predict behavior and disease does not substantially worsen projections, indicating that social information data do not need to be collected to reasonably predict human behavior. This does not necessarily imply a rational risk response, since epidemiological and social information may simply converge on similar perceived risk and behavioral outcomes at the scales we examined. Finally, because our analysis omits some of the least populated counties in the US due to missing data, future work should confirm these findings in more rural parts of the country.

We also evaluated how connected counties could influence social behavior. This analysis revealed that case incidence and risk perception in linked counties is significantly associated with social distancing behavior, regardless of how the counties are connected. The effect of linked county case incidence on focal county social distancing was similar across types of county connections, while the association between social information and social distancing was the strongest for socially connected counties, followed by commuting counties. The effect of policies in linked counties on focal county social distancing was not meaningful. These findings suggest that concerns about COVID-19 may have been disproportionately shared through technology between socially connected counties which aligns with the ease of sharing and spreading opinions on social networking platforms [58, 59]. Our dataset capturing social connection relies on Facebook friendship networks which may not be a representative source for COVID-19 information exchange given that Facebook users skew younger and higher income [60]. Nevertheless, the fact that non-adjacent counties can influence each other could provide an avenue through which interventions could be implemented to calibrate perceived risk with absolute risk. Conversely, there is the potential for spread of misinformation through these social networking platforms. Further work will be needed to disentangle the role of information versus misinformation spread related to behavior through social networking platforms.

In an effort to mimic the assumptions made by classical disease-behavior models, we construct our models so that each county has the same relationship between epidemiological or social information and change in social distancing, i.e., the same risk tolerance. We find that this decision leads to poor model fit, overlooks spatial heterogeneity in behavior change, and, as a result, produces inaccurate disease projections. Considering this type of spatial heterogeneity in epidemiological models is paramount to improving their accuracy and may require additional data collection to understand which counties behave similarly and can be treated homogeneously [33].

Our findings further highlight the importance of other factors related to social distancing behavior that should be taken into account in future work. Warmer temperatures are associated with less social distancing; people are more likely to go places and participate in leisure activities when the weather is warmer [61] and may have shifted social engagements to outdoor settings. Indeed, we found that an increase in outdoor mobility is associated with decreased social distancing. Higher population density was associated with more social distancing; those in more urban areas were likely more adherent to COVID-19 recommendations [62, 63]. Higher household income was also associated with more social distancing, likely due to socioeconomic differences in essential workers: high income individuals could likely telework, whereas lower income workers were more likely to be classified as essential [64]. The main model shows that a higher proportion of the population that is Black or Hispanic is associated with more social distancing; this could be due to differences in underlying health conditions and healthcare access that disproportionately impact Black and Hispanic communities, leading them to be more concerned about infection and reduce their contacts [65]. Further work on racial differences in the ability to social distance is necessary to better understand this difference.

Overall, our work demonstrates the associations of policies and perceptions with mitigation behavior. We have empirically identified which factors altered behavior during the COVID-19 pandemic and the spatial scale of information to which people are most responsive. These findings underscore the importance of coordinating public health policies across geographical scales and highlight that localized disease information is not readily available or relied upon. We have demonstrated the downstream effects of the observed behavior changes on transmission dynamics and delineated the consequences of these findings for coupled disease-behavior models. Our results show that broader-scale information, whether epidemiological or social, is more strongly linked to behavior change than local information. Future work should investigate whether this response reflects rational risk assessment, and should further explore heterogeneity in risk response across individuals and populations.

## Data availability

Individual survey responses on social distancing and concern about infection cannot be shared by the authors, but researchers can refer to https://cmu-delphi.github.io/delphi-epidata/symptom-survey/data-access.html if they would like to enter an agreement for data usage with CMU Delphi. Aggregated data for all variables to reproduce main and supplementary analyses are available at https://github.com/bansallab/policy_perception. Spatiotemporal regression numerical results are available in Supplementary Data files 1-3. Case incidence and death data were obtained from the New York Times (https://github.com/nytimes/covid-19-data/tree/master/rolling-averages). Policy data were obtained from the US Department of Health and Human Services (https://healthdata.gov/dataset/COVID-19-State-and-County-Policy-Orders/gyqz-9u7n/about_data).

## Code availability

Code to reproduce main and supplementary analyses is available at https://github.com/bansallab/policy_perception [66]. Analyses were run using R version 4.5.2.

## Authors’ contributions

CMW conceived the study, performed analyses, interpreted findings, and drafted and edited the manuscript. JCT performed analyses, interpreted findings, and drafted and edited the manuscript. VC interpreted the findings and edited the manuscript. SB conceived and supervised the study, interpreted the findings, and edited the manuscript. CMW, JCT, and SB accessed and verified the data. All authors were responsible for the decision to submit the manuscript for publication.

## Acknowledgments

We thank Giulia Pullano for sharing estimates of case underreporting in the US for the study period and Alexes Merritt for summarizing the differences between US policy datasets.

## Supplementary Materials

**Table S1:** Variance Inflation Factors (VIF) for each variable in the main model.

| Variable | VIF |
| --- | --- |
| County incidence | 4.89 |
| State incidence | 7.49 |
| County prop. worried | 1.82 |
| State prop. worried | 5.95 |
| County restaurant/bar closure | 3.65 |
| County worship restrictions | 2.62 |
| County outdoor recreation restrictions | 3.05 |
| County non-essential business restrictions | 5.68 |
| County entertainment restrictions | 3.24 |
| County manufacturing restrictions | 2.38 |
| County childcare restrictions | 2.44 |
| State restaurant/bar closure | 2.22 |
| State worship restrictions | 1.86 |
| State outdoor recreation restrictions | 2.84 |
| State non-essential business restrictions | 3.40 |
| State entertainment restrictions | 2.58 |
| State manufacturing restrictions | 1.38 |
| State government response index | 1.90 |
| Min. temperature | 2.92 |
| Max. windspeed | 1.18 |
| Precipitation | 1.08 |
| Population density | 1.20 |
| Prop. graduated high school | 1.16 |
| Median income | 1.25 |
| Prop. Black | 1.17 |
| Prop. Hispanic | 1.08 |
| Prop. essential workers | 1.05 |
| County indoor activity index | 1.92 |
| State indoor activity index | 3.43 |
| Holiday week | 1.07 |

**Table S2:** Variance Inflation Factors (VIF) for each variable in the neighboring county model.

| Variable | VIF |
| --- | --- |
| Focal county incidence | 5.98 |
| Neighboring county incidence | 6.97 |
| Focal county prop. worried | 1.67 |
| Neighboring county prop. worried | 2.53 |
| Focal county restaurant/bar closure | 4.50 |
| Neighboring county restaurant/bar closure | 3.87 |
| Focal county worship restrictions | 3.19 |
| Neighboring county worship restrictions | 3.44 |
| Focal county outdoor recreation restrictions | 3.71 |
| Neighboring county outdoor recreation restrictions | 3.39 |
| Focal county non-essential business restrictions | 7.03 |
| Neighboring county non-essential business restrictions | 6.55 |
| Focal county entertainment restrictions | 4.28 |
| Neighboring county entertainment restrictions | 3.89 |
| Focal county manufacturing restrictions | 3.30 |
| Neighboring county manufacturing restrictions | 3.71 |
| Focal county childcare restrictions | 3.30 |
| Neighboring country childcare restrictions | 3.38 |
| Min. temperature | 2.25 |
| Max. windspeed | 1.16 |
| Precipitation | 1.07 |
| Population density | 1.20 |
| Prop. graduated high school | 1.16 |
| Median income | 1.25 |
| Prop. Black | 1.17 |
| Prop. Hispanic | 1.08 |
| Prop. essential workers | 1.05 |
| Focal county indoor activity index | 1.41 |
| Holiday week | 1.07 |

**Table S3:** Variance Inflation Factors (VIF) for each variable in the commuting county model.

| Variable | VIF |
| --- | --- |
| Focal county incidence | 4.57 |
| Commuting county incidence | 7.72 |
| Focal county prop. worried | 1.74 |
| Commuting county prop. worried | 3.86 |
| Focal county restaurant/bar closure | 3.90 |
| Commuting county restaurant/bar closure | 2.15 |
| Focal county worship restrictions | 2.69 |
| Commuting county worship restrictions | 1.68 |
| Focal county outdoor recreation restrictions | 3.18 |
| Commuting county outdoor recreation restrictions | 2.35 |
| Focal county non-essential business restrictions | 5.86 |
| Commuting county non-essential business restrictions | 3.19 |
| Focal county entertainment restrictions | 3.49 |
| Commuting county entertainment restrictions | 1.80 |
| Focal county manufacturing restrictions | 2.65 |
| Commuting county manufacturing restrictions | 1.67 |
| Focal county childcare restrictions | 2.60 |
| Commuting county childcare restrictions | 1.55 |
| Min. temperature | 2.52 |
| Max. windspeed | 1.17 |
| Precipitation | 1.08 |
| Population density | 1.20 |
| Prop. graduated high school | 1.16 |
| Median income | 1.24 |
| Prop. Black | 1.17 |
| Prop. Hispanic | 1.08 |
| Prop. essential workers | 1.05 |
| Focal county indoor activity index | 1.41 |
| Holiday week | 1.08 |

**Table S4:** Variance Inflation Factors (VIF) for each variable in the socially connected county model.

| Variable | VIF |
| --- | --- |
| Focal county incidence | 3.45 |
| Socially connected county incidence | 7.13 |
| Focal county prop. worried | 1.77 |
| Socially connected county prop. worried | 4.68 |
| Focal county restaurant/bar closure | 3.82 |
| Socially connected county restaurant/bar closure | 4.24 |
| Focal county worship restrictions | 2.67 |
| Socially connected county worship restrictions | 3.40 |
| Focal county outdoor recreation restrictions | 3.09 |
| Socially connected county outdoor recreation restrictions | 2.03 |
| Focal county non-essential business restrictions | 5.74 |
| Socially connected county non-essential business restrictions | 3.17 |
| Focal county entertainment restrictions | 3.37 |
| Socially connected county entertainment restrictions | 1.44 |
| Focal county manufacturing restrictions | 2.55 |
| Socially connected county manufacturing restrictions | 1.27 |
| Focal county childcare restrictions | 2.53 |
| Socially connected county childcare restrictions | 1.42 |
| Min. temperature | 2.92 |
| Max. windspeed | 1.19 |
| Precipitation | 1.08 |
| Population density | 1.20 |
| Prop. graduated high school | 1.16 |
| Median income | 1.25 |
| Prop. Black | 1.17 |
| Prop. Hispanic | 1.08 |
| Prop. essential workers | 1.05 |
| Focal county indoor activity index | 1.44 |
| Holiday week | 1.15 |

**Table S5:** Based on R^2^, univariate models state-level variables explain more variation in social distancing than county-level variables. Social information is measured using the right-aligned 3 week rolling average proportion of CTIS respondents reporting that they are somewhat or very worried about someone in their immediate family becoming seriously ill from COVID-19. Epidemiological information is based on the right-aligned 3 week rolling average of new COVID-19 cases. Data are pooled across all counties for the period from September 6, 2020 through January 31, 2021.

| Predictor scale | Epidemiological information | Social information |
| --- | --- | --- |
| County | 0.287 | 0.167 |
| State | 0.357 | 0.358 |

**Figure S1:**
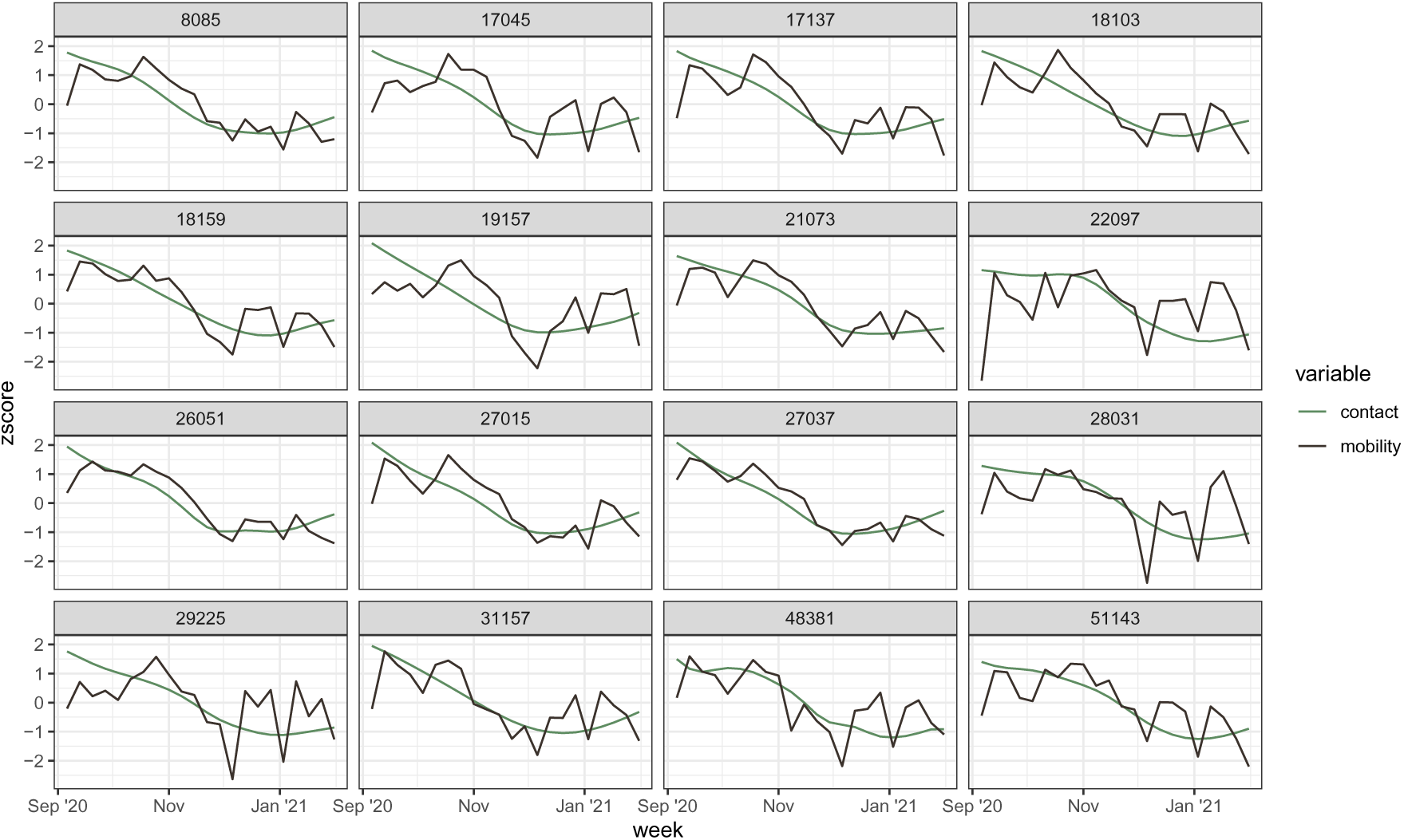
Contact versus mobility time series. The two indicators of social distancing generally follow the same temporal trend across a random sample of US counties. We perform sensitivity analyses using mobility data instead of contact as the response in our regression models to demonstrate the robustness of our results.

**Figure S2:**
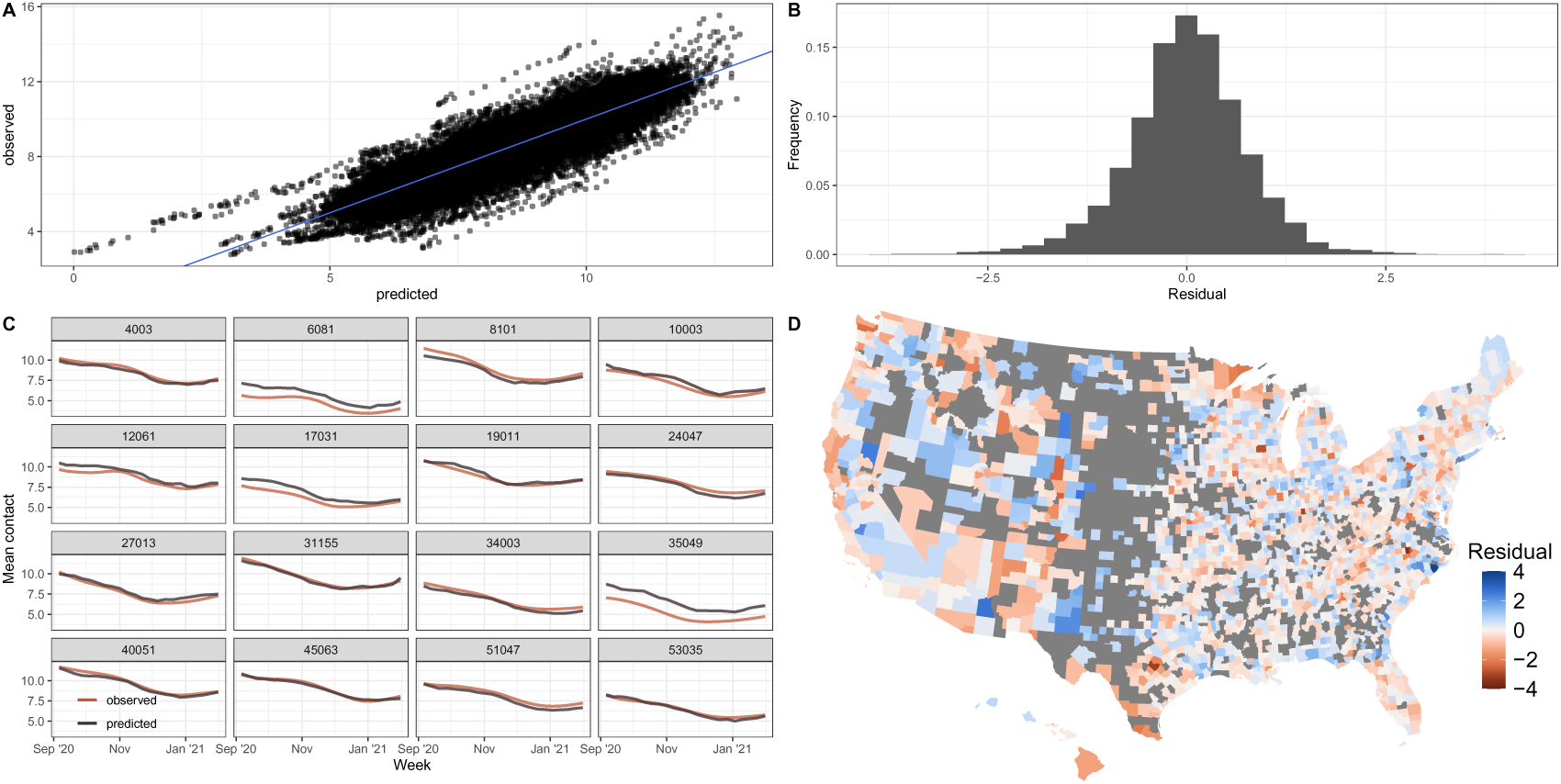
Model diagnostics for main regression. (A) Observed vs predicted contact rates, as a measure of social distancing. Blue line represents y = x. Predicted contact rates are positively associated with observed rates which indicates a good model fit. (B) Distribution of residuals. Distribution is centered around zero which indicates good model fit. (C) Observed vs predicted time series. Predicted time series generally match observed time series which indicates good model fit. (D) Spatial distribution of residuals. There is no clear spatial autocorrelation of residuals, indicating that we are not missing key predictors in our model.

**Figure S3:**
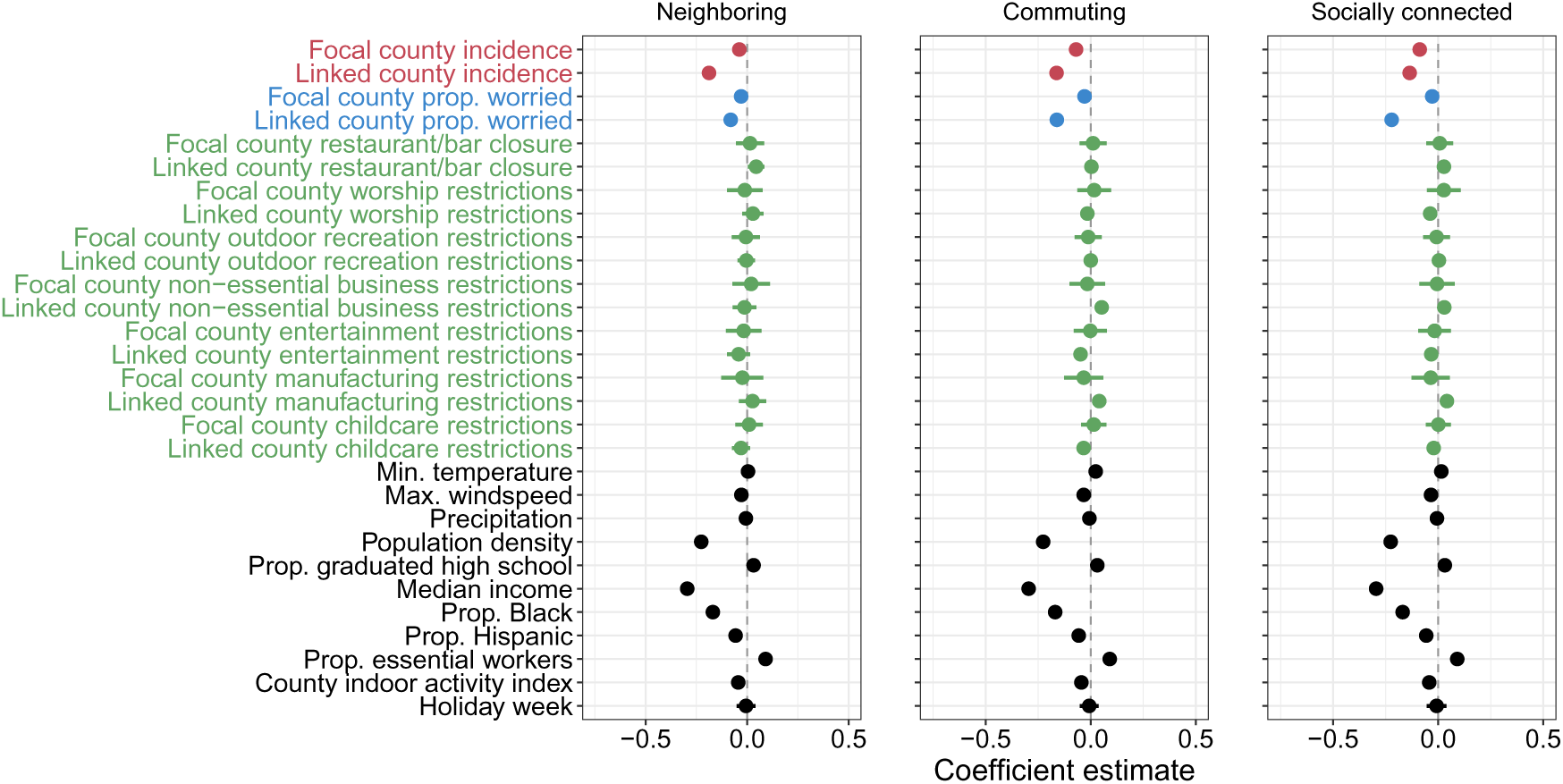
Coefficient estimates and 95% credible intervals of spatiotemporal regressions predicting county-week mean number of contacts per day using intermediate spatial scales. This analysis is performed for September 2020 through Jan 2021. This figure is for the same model as in the main text (Figure 2) but shows coefficients for all covariates in the models, not just the linked county covariates (as in the main figure). Neighboring counties share a border with the focal county. Commuting counties are connected to the focal county via commuting flows. Socially connected counties are connected to the focal county through friendships on Facebook. Focal county non-essential business restrictions were dropped from the models due to collinearity.

**Figure S4:**
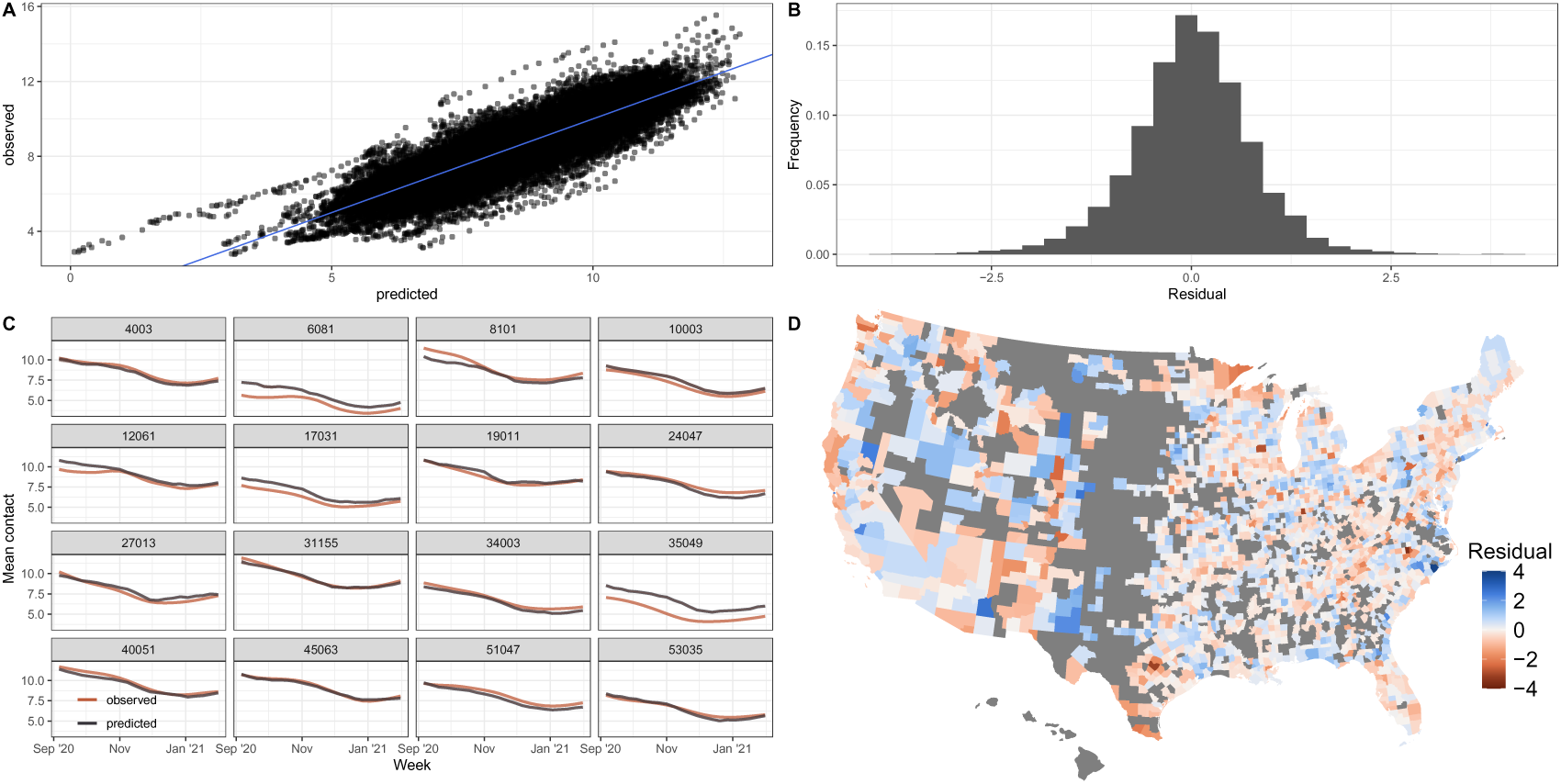
Model diagnostics for neighboring county regression. (A) Observed vs predicted contact rates, as a measure of social distancing. Blue line represents y = x. Predicted contact rates are positively associated with observed rates which indicates a good model fit. (B) Distribution of residuals. Distribution is centered around zero which indicates good model fit. (C) Observed vs predicted time series. Predicted time series generally match observed time series which indicates good model fit. (D) Spatial distribution of residuals. There is no clear spatial autocorrelation of residuals, indicating that we are not missing key predictors in our model.

**Figure S5:**
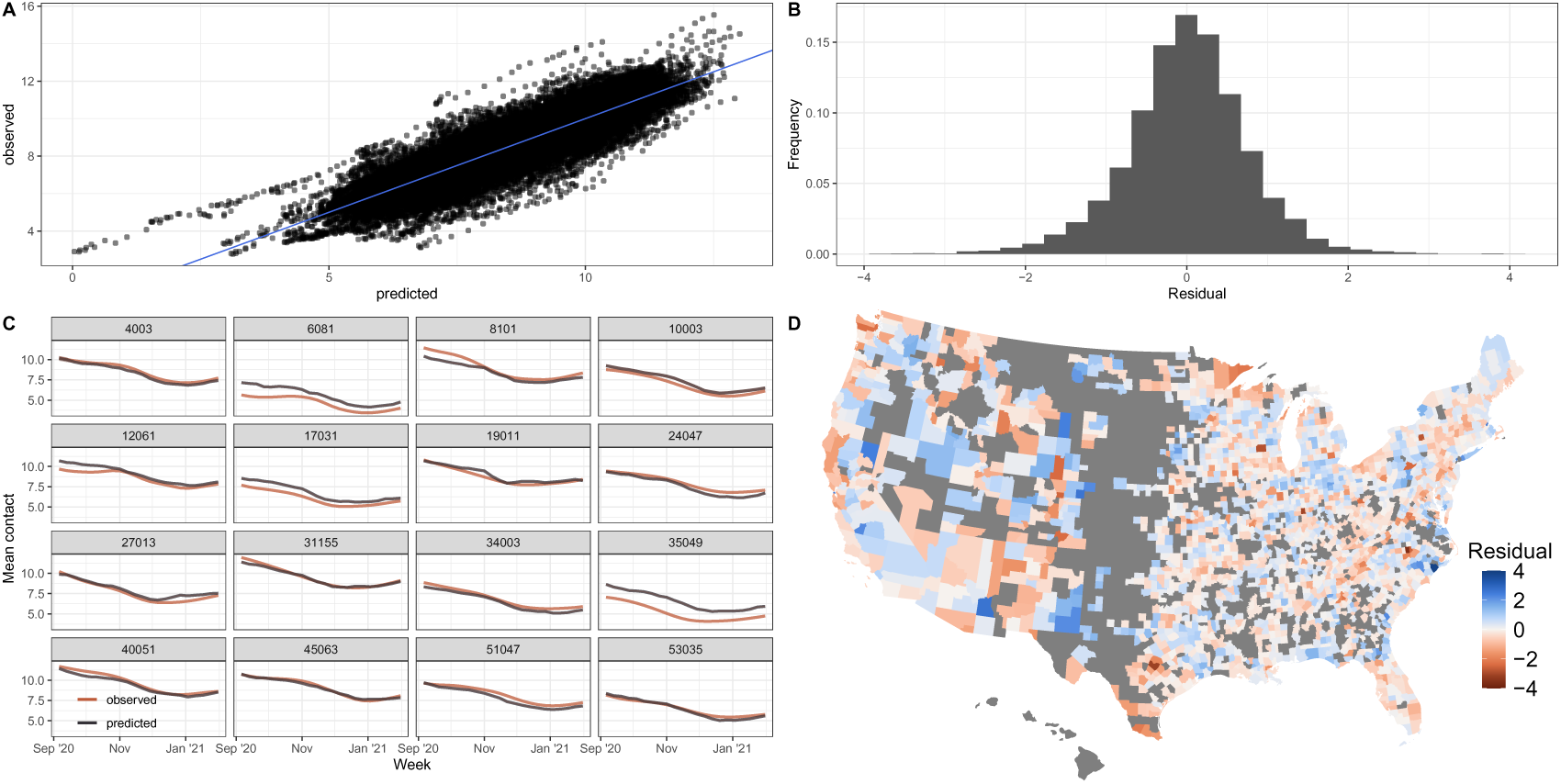
Model diagnostics for commuting county regression. (A) Observed vs predicted contact rates, as a measure of social distancing. Blue line represents y = x. Predicted contact rates are positively associated with observed rates which indicates a good model fit. (B) Distribution of residuals. Distribution is centered around zero which indicates good model fit. (C) Observed vs predicted time series. Predicted time series generally match observed time series which indicates good model fit. (D) Spatial distribution of residuals. There is no clear spatial autocorrelation of residuals, indicating that we are not missing key predictors in our model.

**Figure S6:**
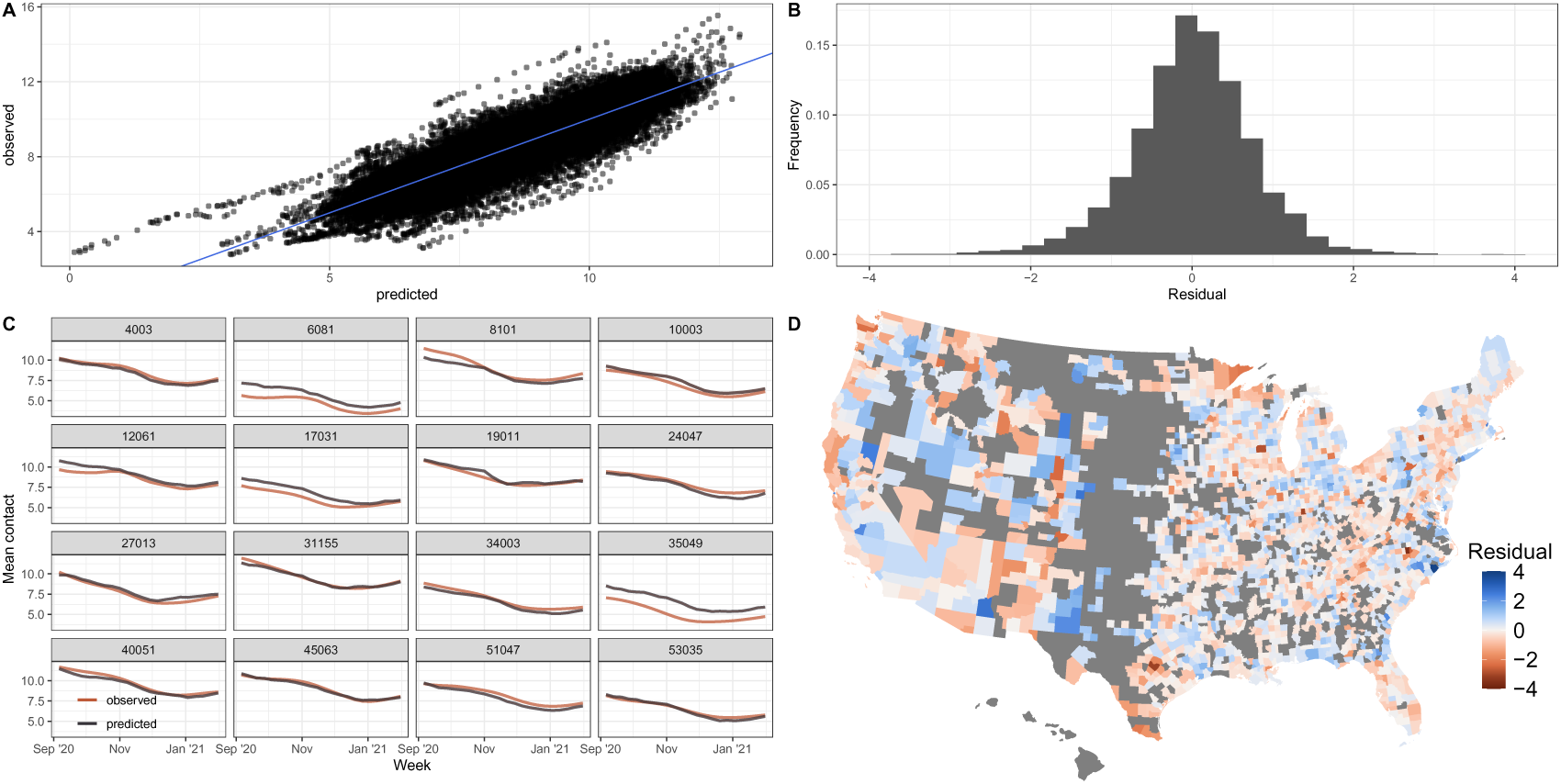
Model diagnostics for socially connected county regression. (A) Observed vs predicted contact rates, as a measure of social distancing. Blue line represents y = x. Predicted contact rates are positively associated with observed rates which indicates a good model fit. (B) Distribution of residuals. Distribution is centered around zero which indicates good model fit. (C) Observed vs predicted time series. Predicted time series generally match observed time series which indicates good model fit. (D) Spatial distribution of residuals. There is no clear spatial autocorrelation of residuals, indicating that we are not missing key predictors in our model.

**Figure S7:**
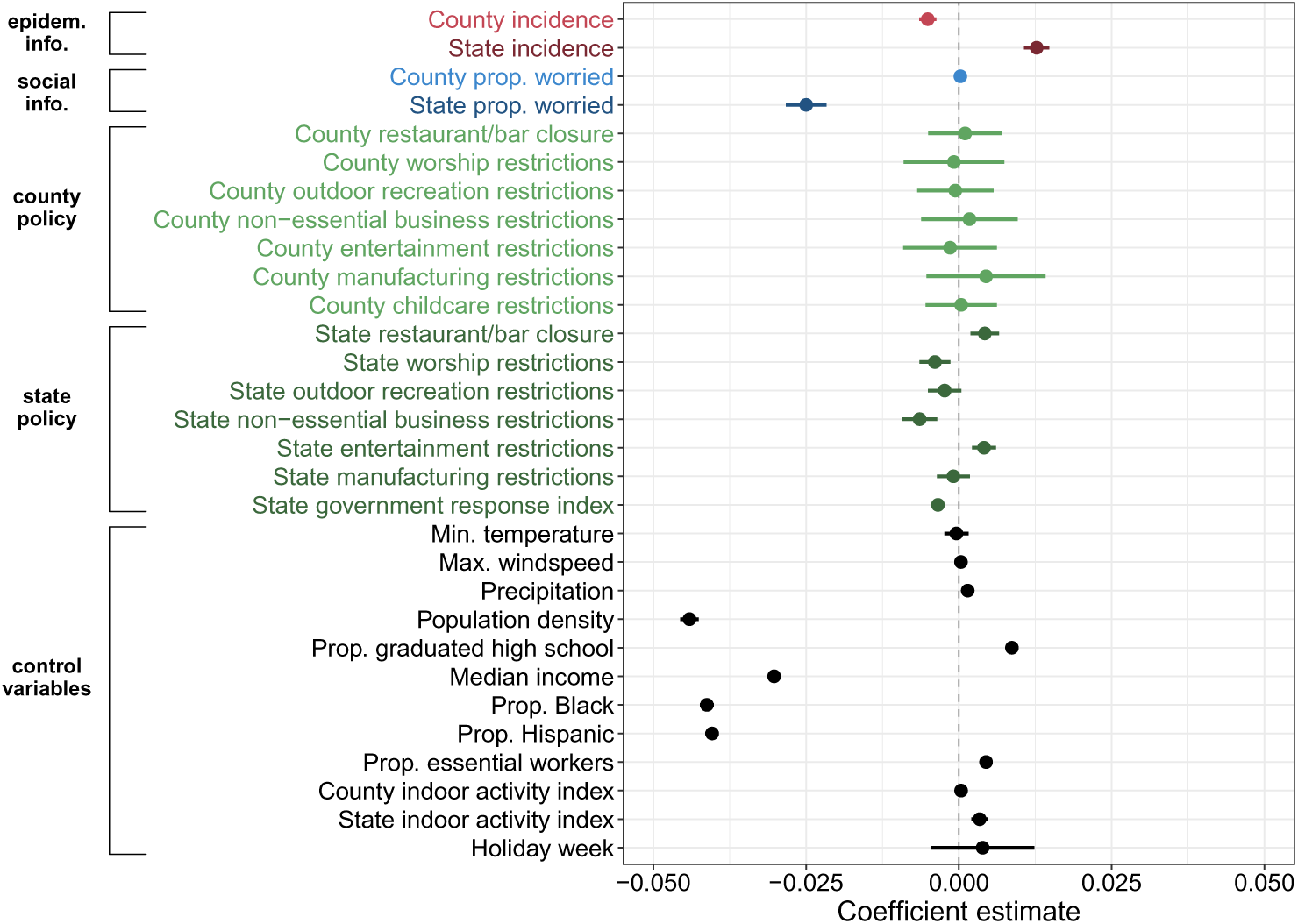
Main regression with mobility as the response instead of contact. Coefficient estimates and 95% credible intervals of the spatiotemporal regression predicting county-week mean mobility, as a measure of social distancing. Positive coefficients indicate higher mobility and lower social distancing. This analysis is performed for September 2020 through January 2021. Findings are generally consistent with those found using contact data as the response except that increased state incidence is associated with increased mobility, i.e., decreased social distancing.

**Figure S8:**
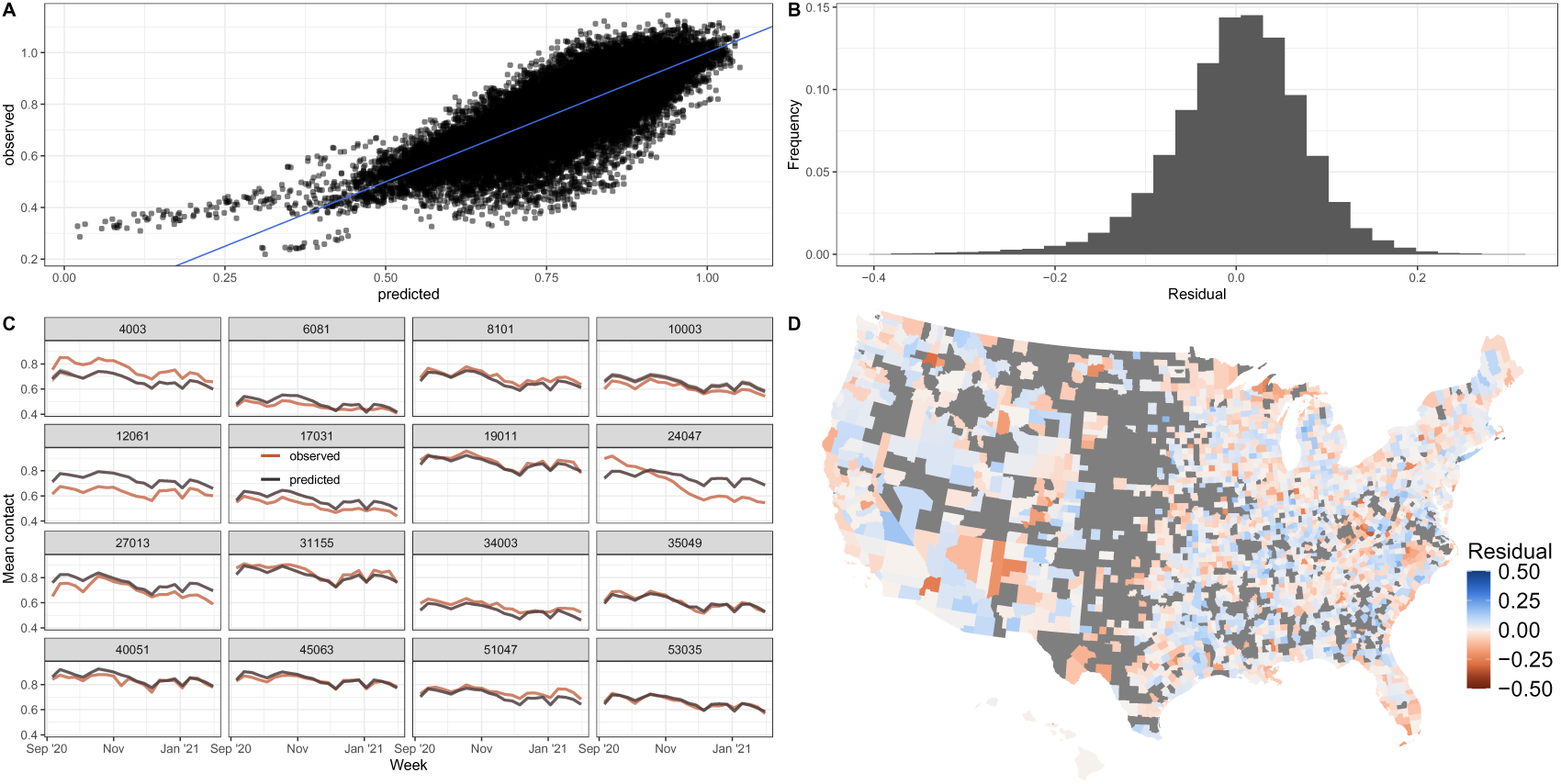
Main regression with mobility as the response instead of contact model diagnostics. (A) Observed vs predicted mobility, as a measure of social distancing. Blue line represents y = x. Predicted mobility is positively associated with observed mobility which indicates a good model fit. (B) Distribution of residuals. Distribution is centered around zero which indicates good model fit. (C) Observed vs predicted time series. Predicted time series generally match observed time series which indicates good model fit. (D) Spatial distribution of residuals. There is no clear spatial autocorrelation of residuals, indicating that we are not missing key predictors in our model.

**Figure S9:**
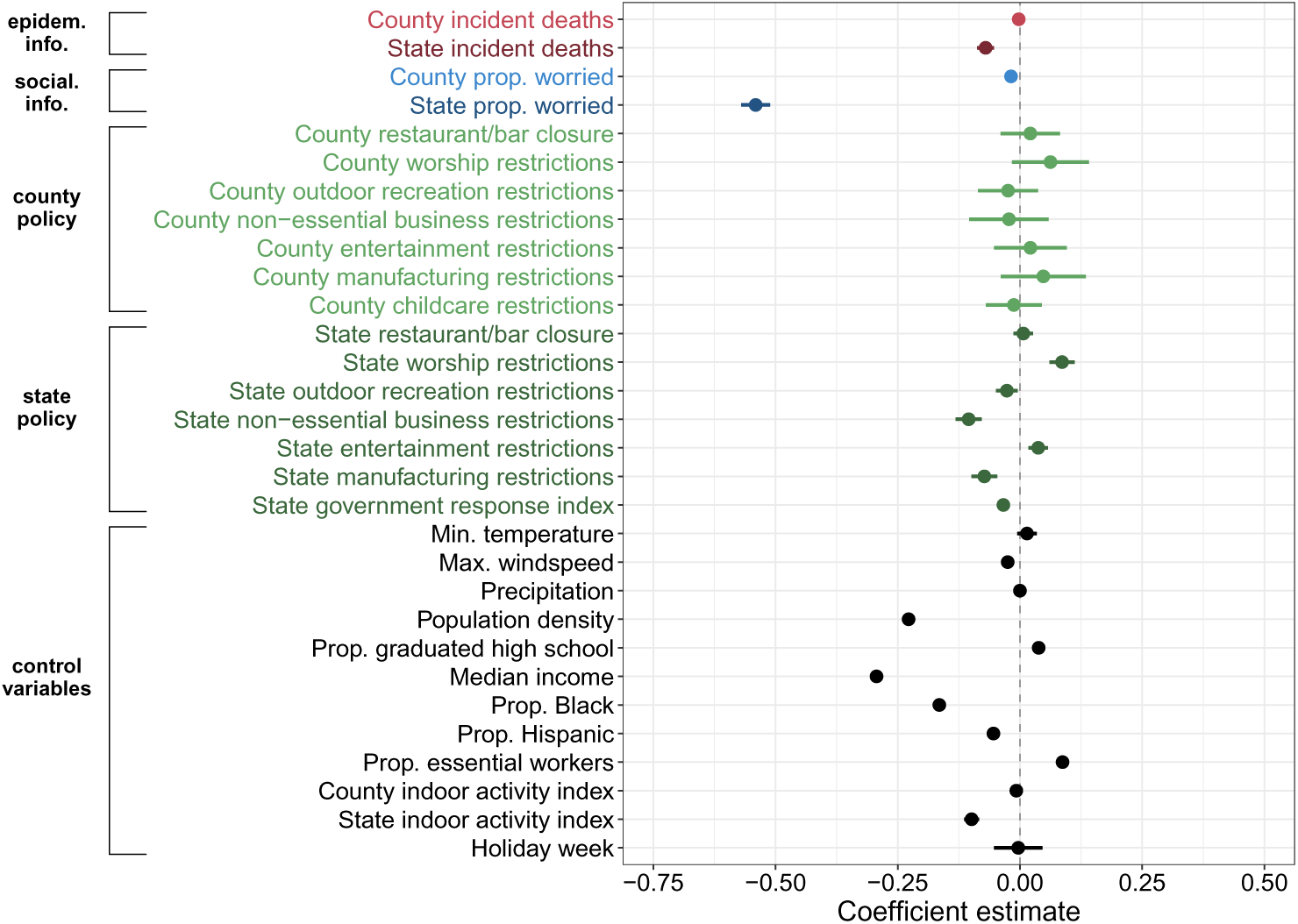
Main regression with incident deaths (4-week rolling average at county level, 3-week at state level as this combination yielded the lowest DIC) instead of incident cases as indicators of epidemiological information. Coefficient estimates and 95% credible intervals of the spatiotemporal regression predicting county-week mean number of non-household contacts per day, as a measure of social distancing. Positive coefficients indicate higher contact rates and lower social distancing. This analysis is performed for September 2020 through January 2021. Findings are consistent with those found using COVID-19 case incidence as a predictor.

**Figure S10:**
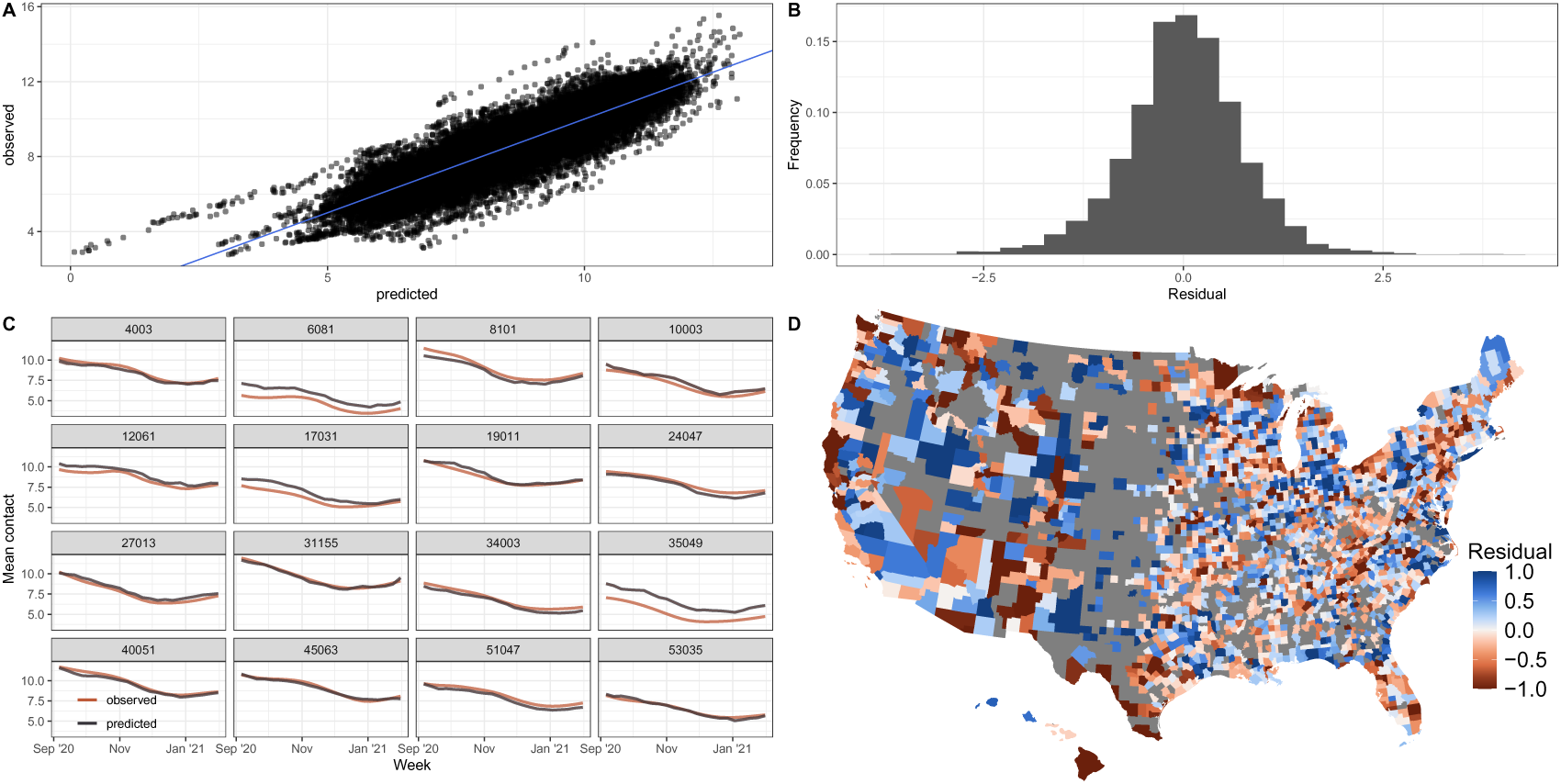
Model diagnostics for main regression with incident deaths (4-week rolling average at county level, 3 week at state level, as this combination yielded the lowest DIC) instead of incident cases as indicators of epidemiological information. (A) Observed vs predicted contact rates, as a measure of social distancing. Blue line represents y = x. Predicted contact rates are positively associated with observed rates which indicates a good model fit. (B) Distribution of residuals. Distribution is centered around zero which indicates good model fit. (C) Observed vs predicted time series. Predicted time series generally match observed time series which indicates good model fit. (D) Spatial distribution of residuals. There is no clear spatial autocorrelation of residuals, indicating that we are not missing key predictors in our model.

**Figure S11:**
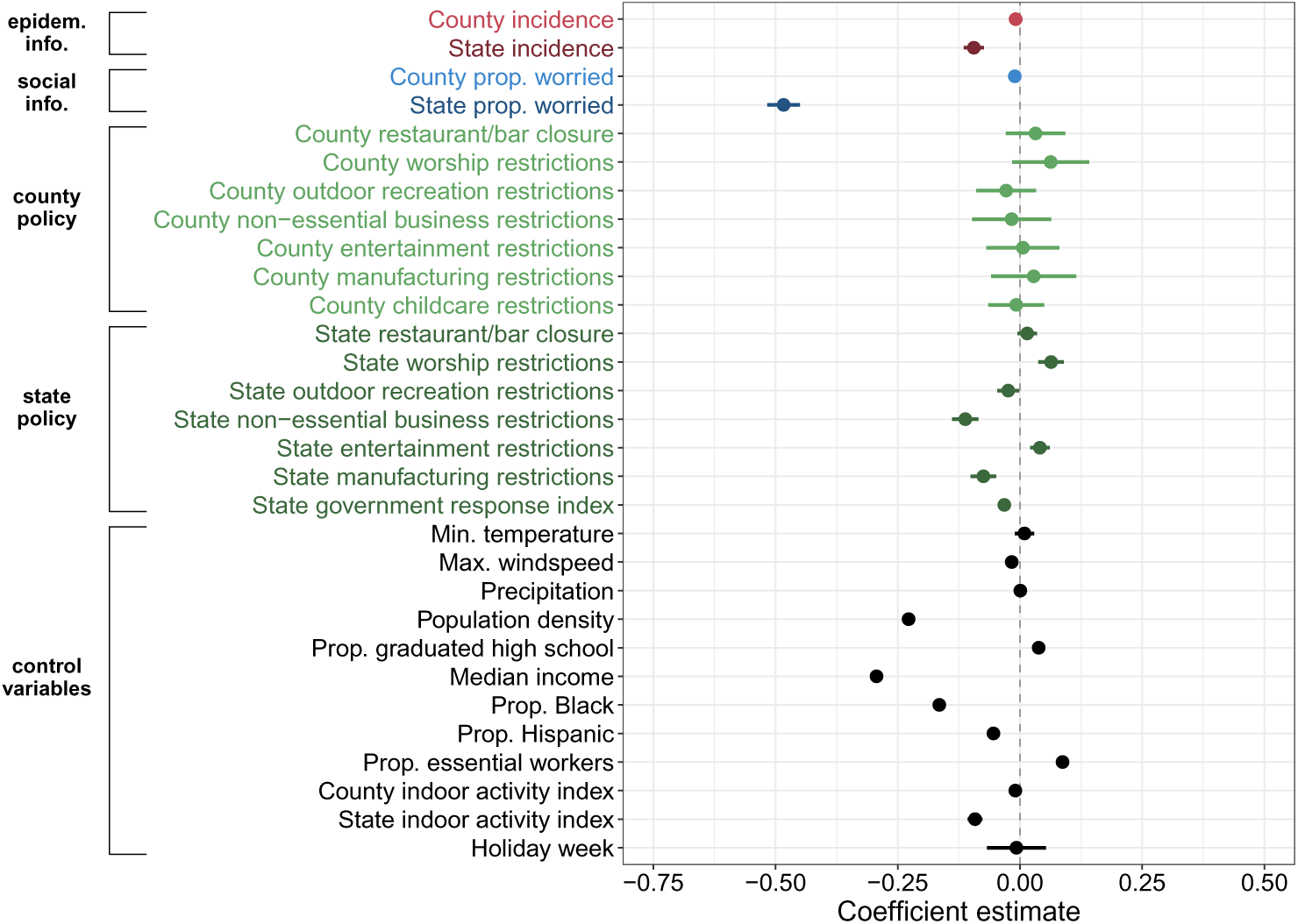
Main regression with worry as a 3 week rolling average, cases as 2 week rolling average. Coefficient estimates and 95% credible intervals of the spatiotemporal regression predicting county-week mean number of non-household contacts per day, as a measure of social distancing. Positive coefficients indicate higher contact rates and lower social distancing. This analysis is performed for September 2020 through January 2021. Findings are consistent with those found by smoothing disease and worry data over longer time periods.

**Figure S12:**
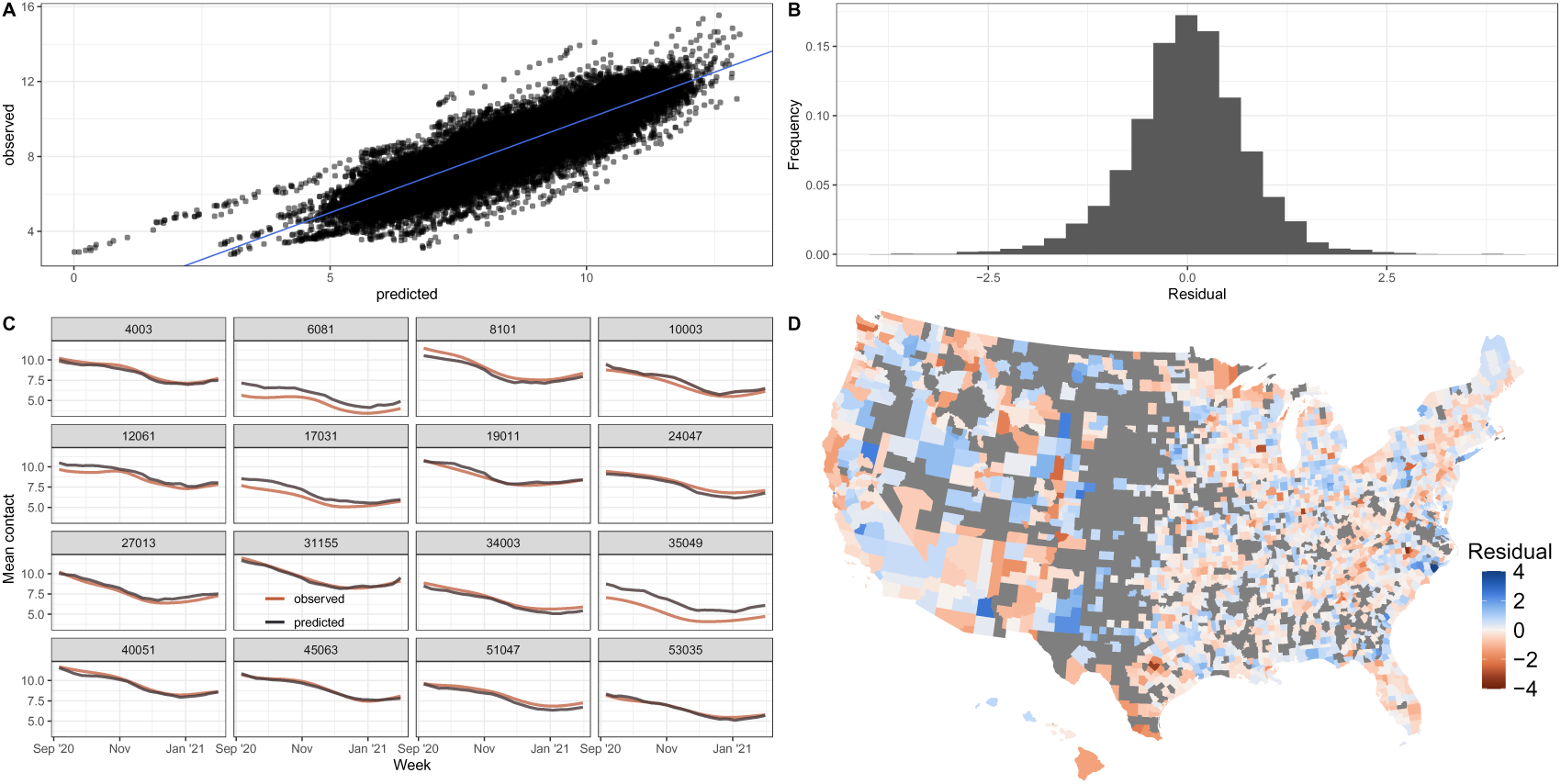
Diagnostics for main regression with worry as a 3 week rolling average, cases as 2 week rolling average. (A) Observed vs predicted contact rates, as a measure of social distancing. Blue line represents y = x. Predicted contact rates are positively associated with observed rates which indicates a good model fit. (B) Distribution of residuals. Distribution is centered around zero which indicates good model fit. (C) Observed vs predicted time series. Predicted time series generally match observed time series which indicates good model fit. (D) Spatial distribution of residuals. There is no clear spatial autocorrelation of residuals, indicating that we are not missing key predictors in our model.

**Figure S13:**
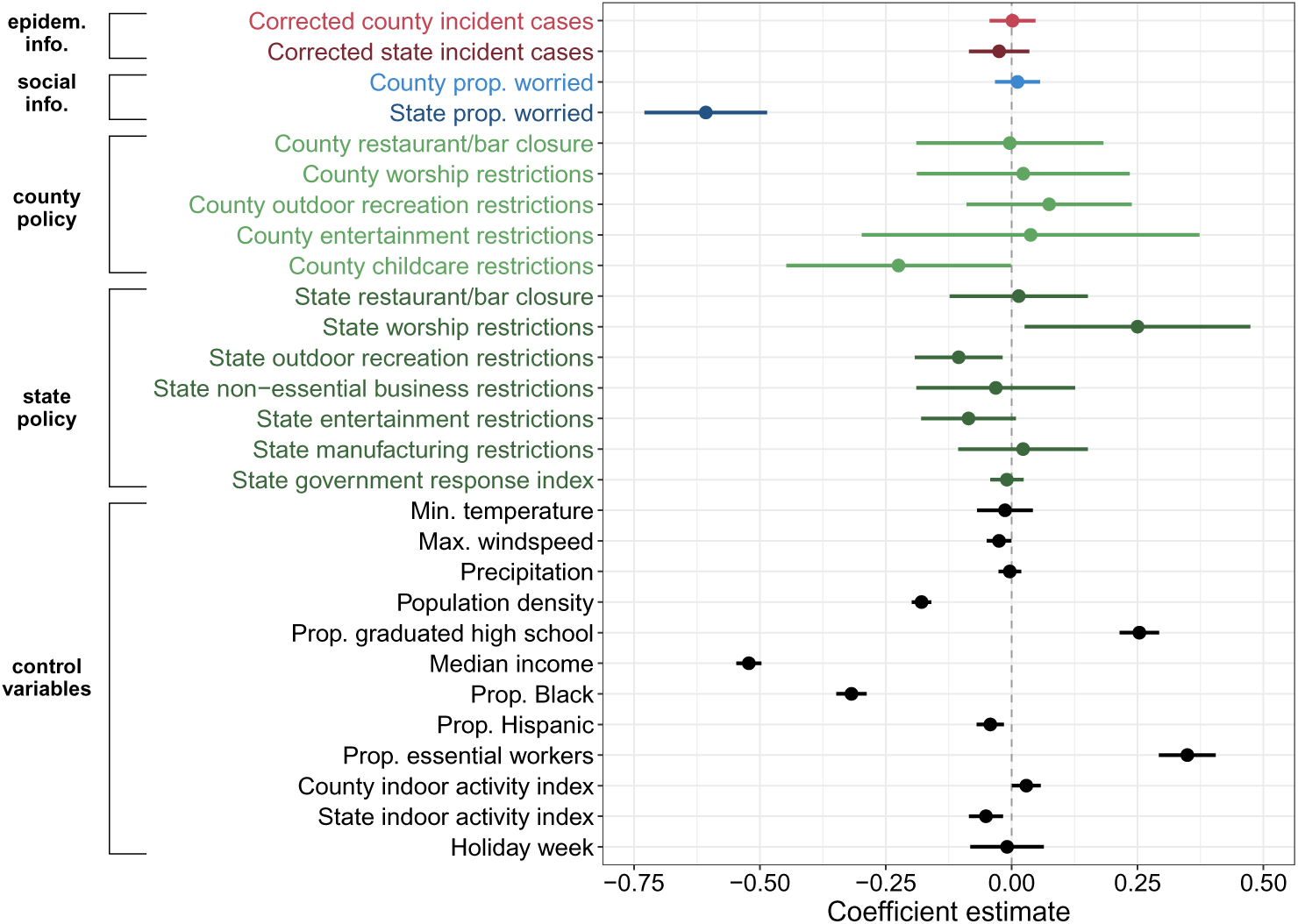
Main regression with incident cases corrected for underreporting as indicators of epidemiological risk. Worry is 3 week rolling average, corrected cases aren’t rolled, as this choice yielded the lowest DIC. This analysis is limited to 239 counties for which we have estimates of underreporting rates. There are no changes in county manufacturing policies in these counties during the study period, as a result these covariates have been dropped. County non-essential business restrictions covariates were dropped due to collinearity. Coefficient estimates and 95% credible intervals of the spatiotemporal regression predicting county-week mean number of non-household contacts per day, as a measure of social distancing. Positive coefficients indicate higher contact rates and lower social distancing. This analysis is performed for September 2020 through January 2021. Findings are consistent with those found using uncorrected case incidence as a predictor.

**Figure S14:**
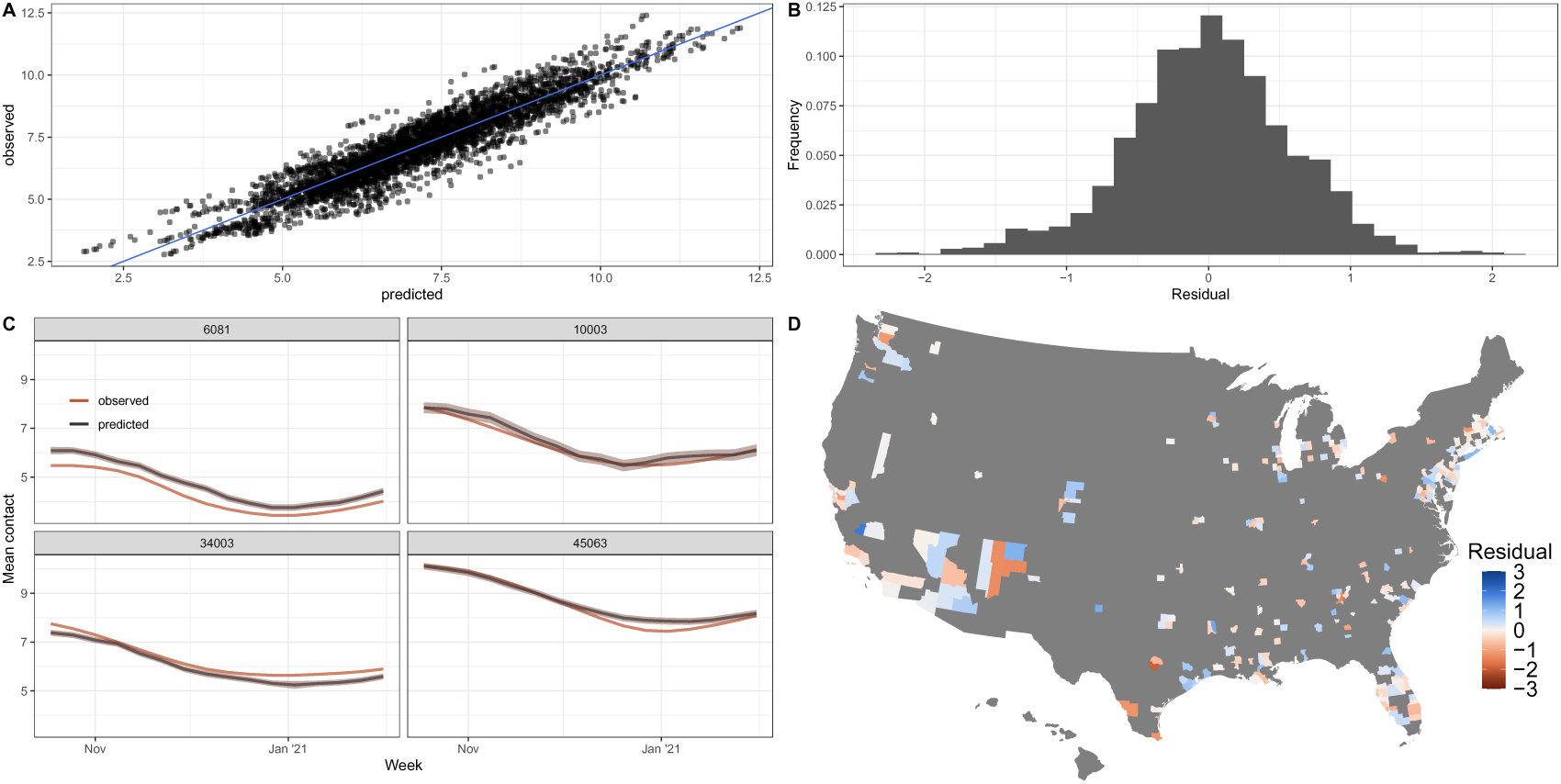
Diagnostics for main regression with incident cases corrected for underreporting as indicators of epidemiological information. Worry is 3 week rolling average, corrected cases aren’t rolled, as this choice yielded the lowest DIC. This analysis is limited to 239 counties for which we have estimates of underreporting rates. (A) Observed vs predicted contact rates, as a measure of social distancing. Blue line represents y = x. Predicted contact rates are positively associated with observed rates which indicates a good model fit. (B) Distribution of residuals. Distribution is centered around zero which indicates good model fit. (C) Observed vs predicted time series. Predicted time series generally match observed time series which indicates good model fit. (D) Spatial distribution of residuals. There is no clear spatial autocorrelation of residuals, indicating that we are not missing key predictors in our model.

**Figure S15:**
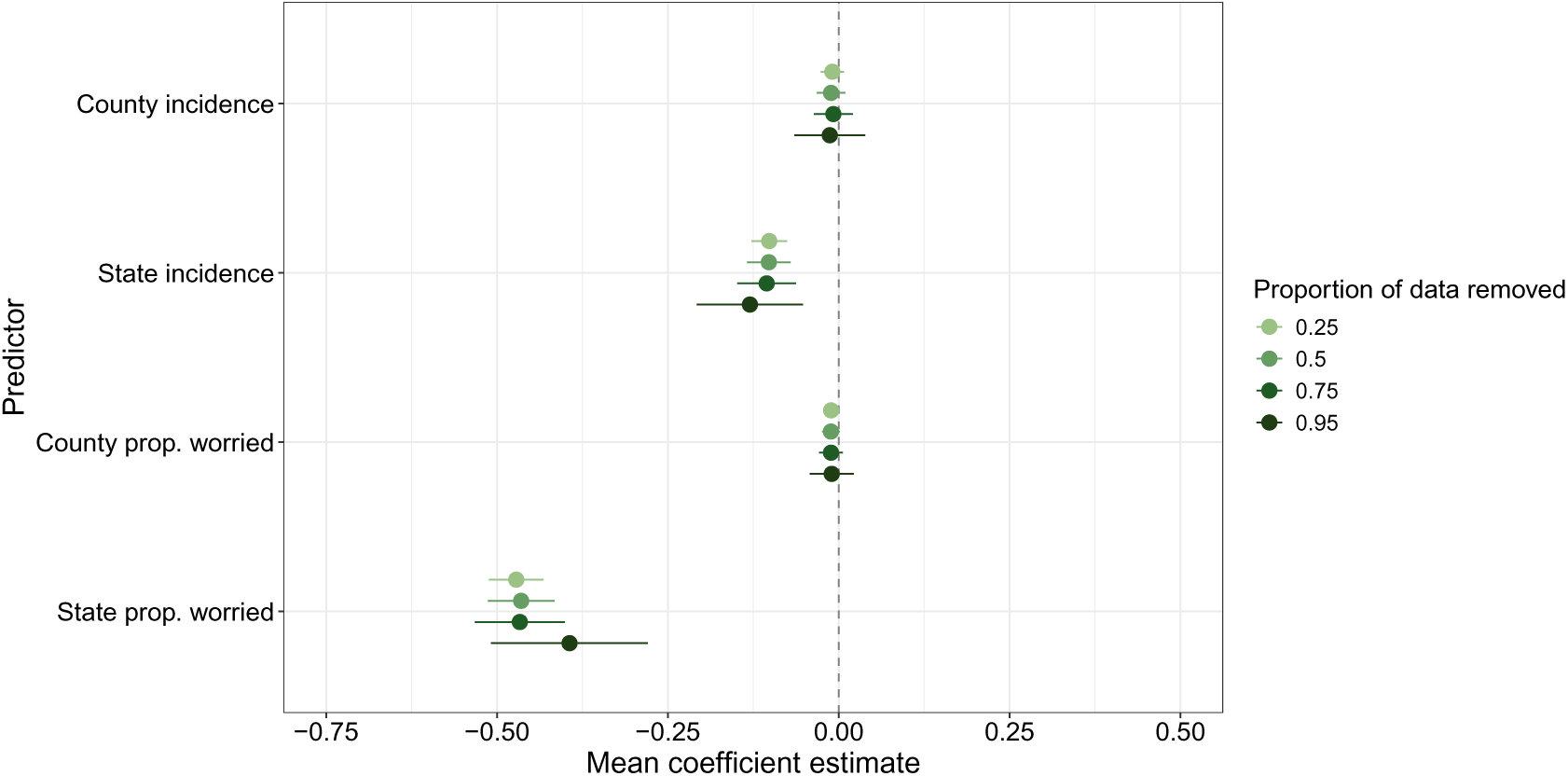
Cross validation of main regression results. A proportion of the counties in the data were removed at random, and the model was re-fit to the remaining data to estimate coefficients. We repeated this process ten times for each omission proportion. Note that all policy and control variables in the main model are included in this analysis but omitted from the plot. Points represent the mean coefficient estimate across the ten replicates, while ranges represent mean lower and upper 95% credible intervals across the ten replicates. Trends are consistent with those in the main text, where state variables are more associated with social distancing than county variables, and social information is more associated than epidemiological information.

**Figure S16:**
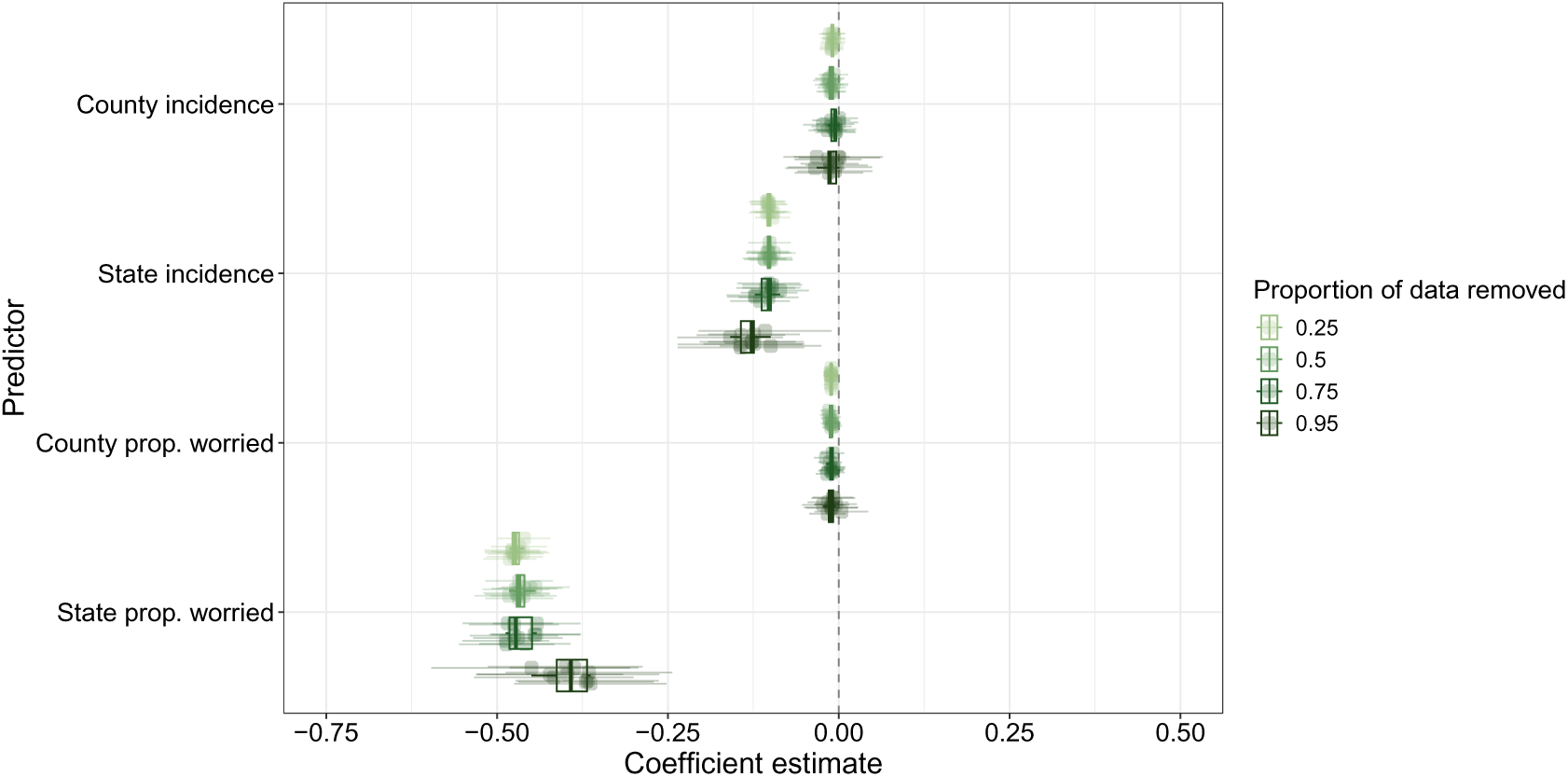
Cross validation of main regression results. A proportion of the counties in the data were removed at random, and the model was re-fit to the remaining data to estimate coefficients. We repeated this process ten times for each omission proportion. Note that all policy and control variables are included in this analysis but omitted from the plot. Points represent coefficient estimates and ranges represent lower and upper 95% credible intervals for each of the ten replicates. Boxplots summarize the mean coefficient estimate across replicates. Trends are consistent with those in the main text, where state variables are more associated with social distancing than county variables, and social information is more associated than epidemiological information.

**Figure S17:**
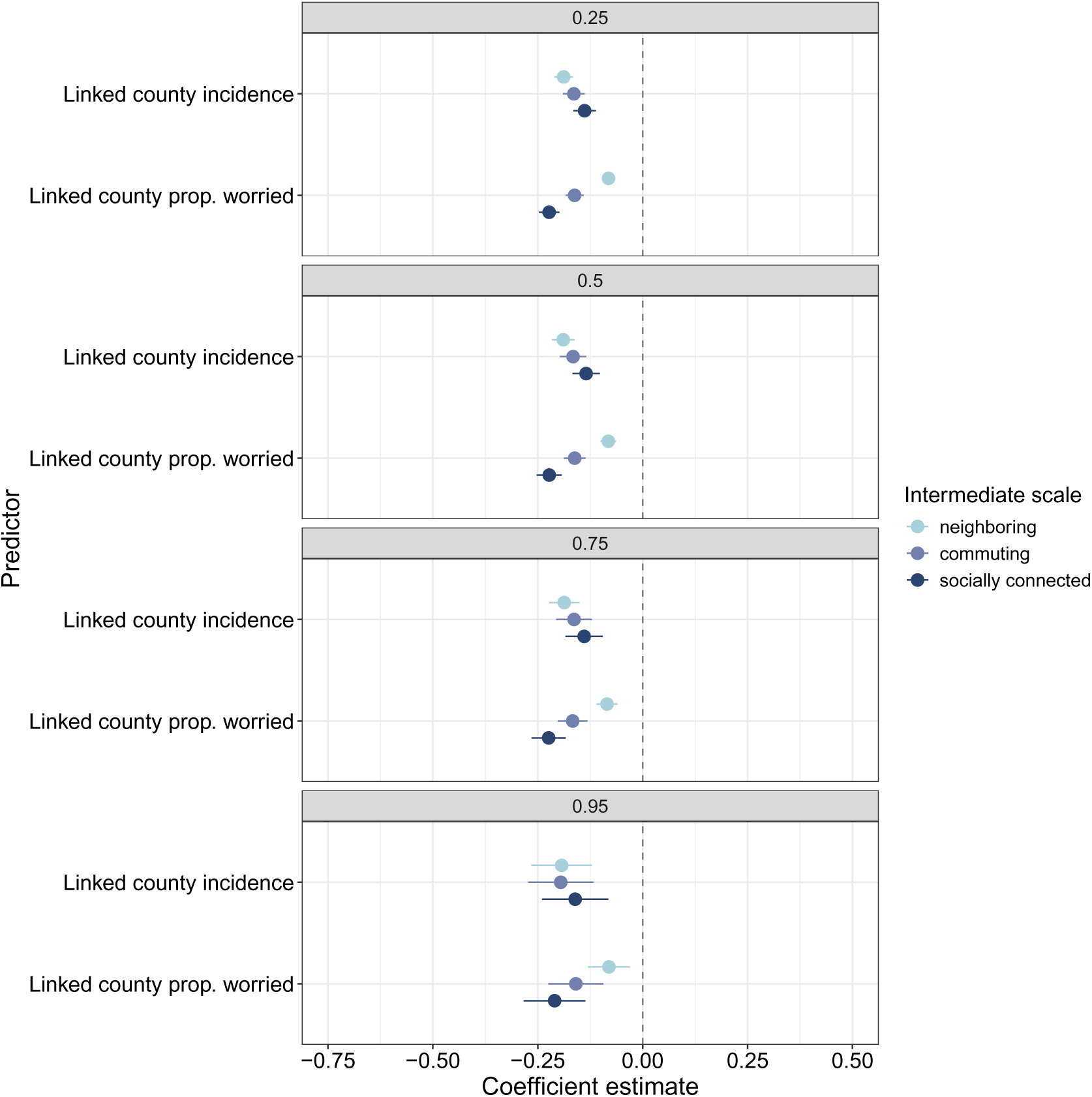
Cross validation of linked county regression results. A proportion of the counties in the data were removed at random, and the model was re-fit to the remaining data to estimate coefficients. We repeated this process ten times for each omission proportion. Note that all policy and control variables are included in this analysis but omitted from the plot. Points represent the mean coefficient estimate across the ten replicates, while ranges represent mean lower and upper 95% credible intervals across the ten replicates. Each facet is the proportion of the counties omitted. Trends are generally consistent with those in the main text.

**Figure S18:**
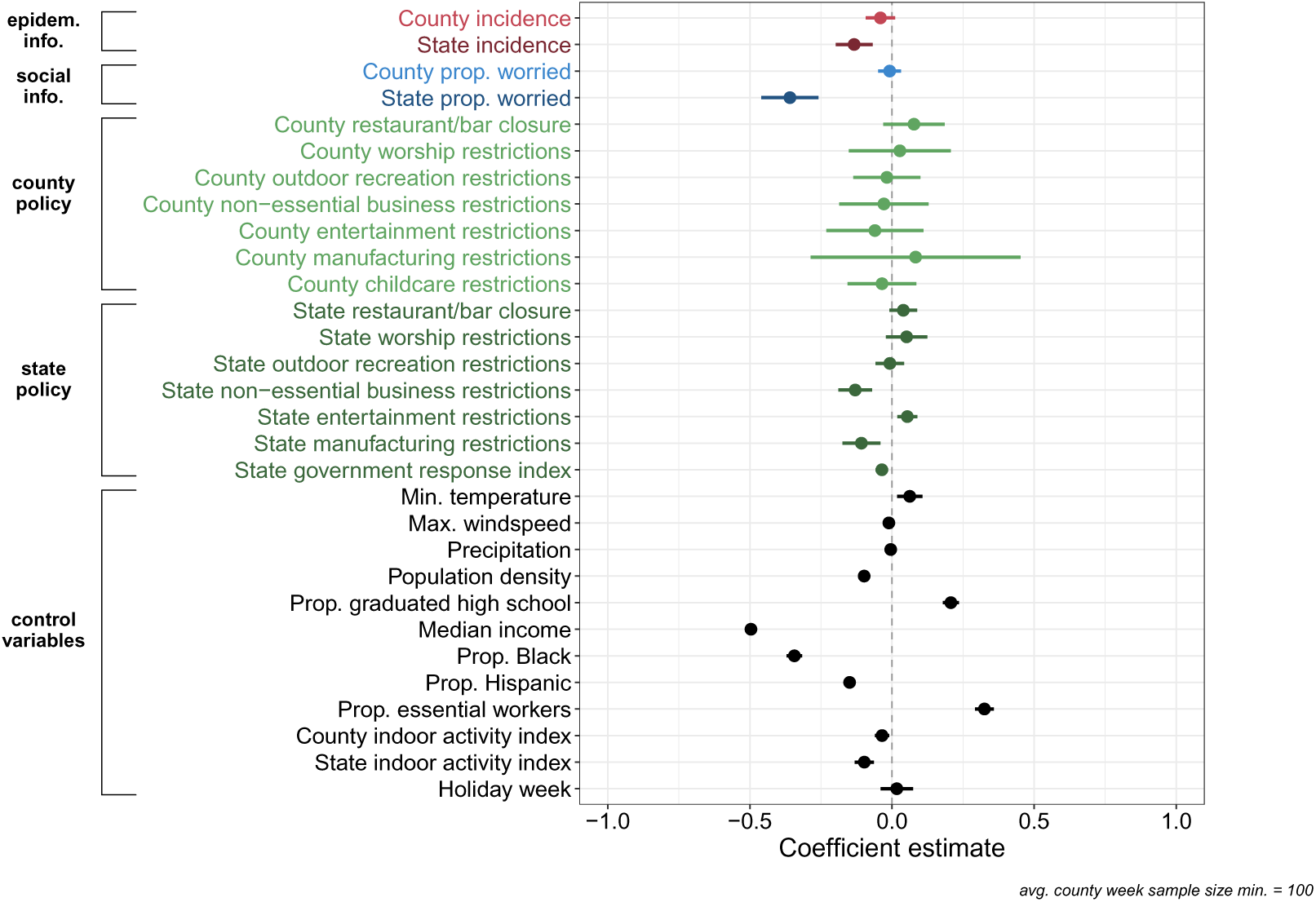
Main regression fit only to counties with an average weekly sample size greater than 100 to test sensitivity of county proportion worried coefficient estimates to sample size. Coefficient estimates and 95% credible intervals of the spatiotemporal regression predicting county-week mean number of non-household contacts per day, as a measure of social distancing. Positive coefficients indicate higher contact rates and lower social distancing. This analysis is performed for September 2020 through January 2021. Even after removing low sample size counties, county proportion worried is not predictive of county social distancing behavior.

**Figure S19:**
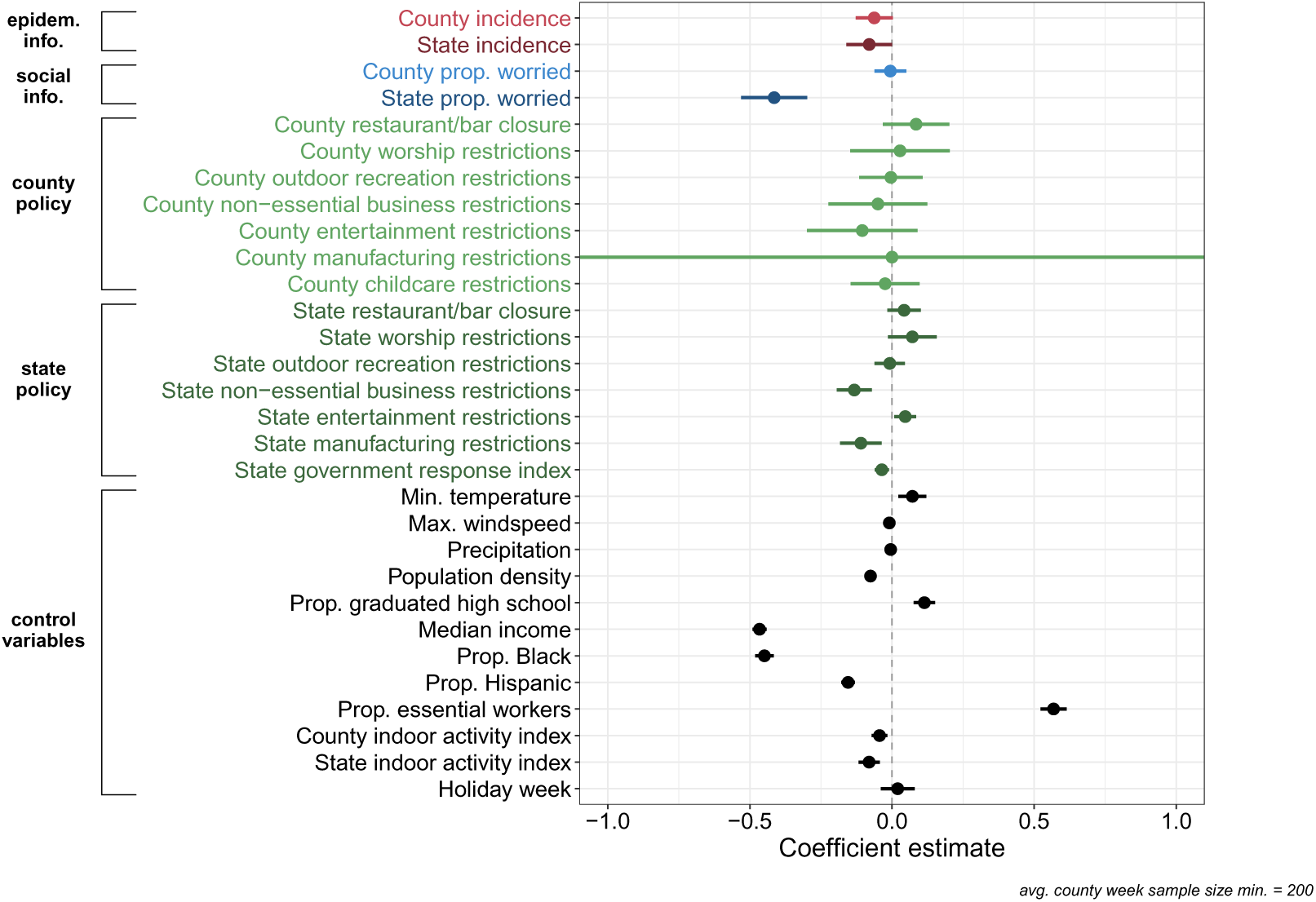
Main regression fit only to counties with an average weekly sample size greater than 200 to test sensitivity of county proportion worried coefficient estimates to sample size. Coefficient estimates and 95% credible intervals of the spatiotemporal regression predicting county-week mean number of non-household contacts per day, as a measure of social distancing. Positive coefficients indicate higher contact rates and lower social distancing. This analysis is performed for September 2020 through January 2021. Even after removing low sample size counties, county proportion worried is not predictive of county social distancing behavior.

**Figure S20:**
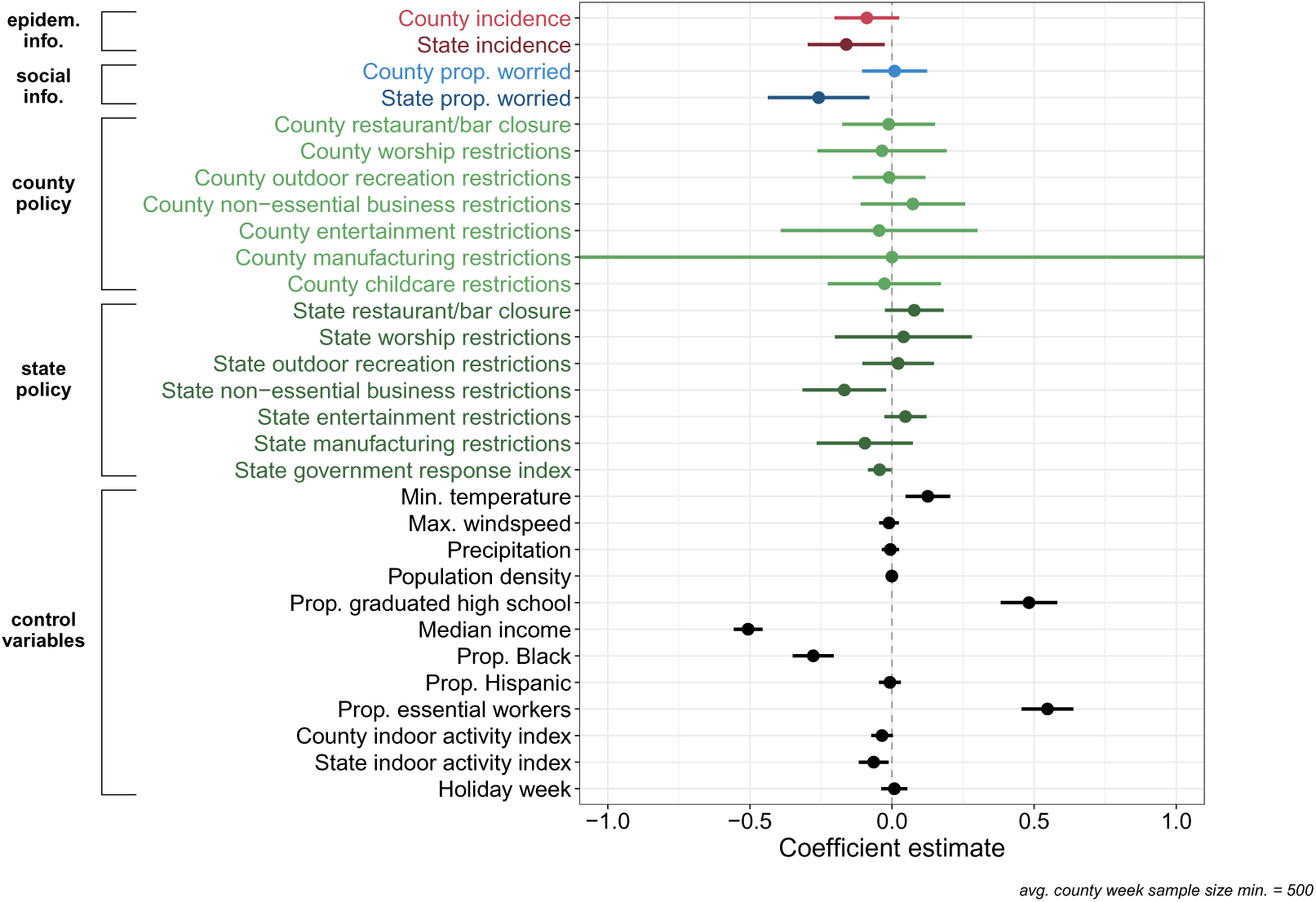
Main regression fit only to counties with an average weekly sample size greater than 500 to test sensitivity of county proportion worried coefficient estimates to sample size. Coefficient estimates and 95% credible intervals of the spatiotemporal regression predicting county-week mean number of non-household contacts per day, as a measure of social distancing. Positive coefficients indicate higher contact rates and lower social distancing. This analysis is performed for September 2020 through January 2021. Even after removing low sample size counties, county proportion worried is not predictive of county social distancing behavior.

**Figure S21:**
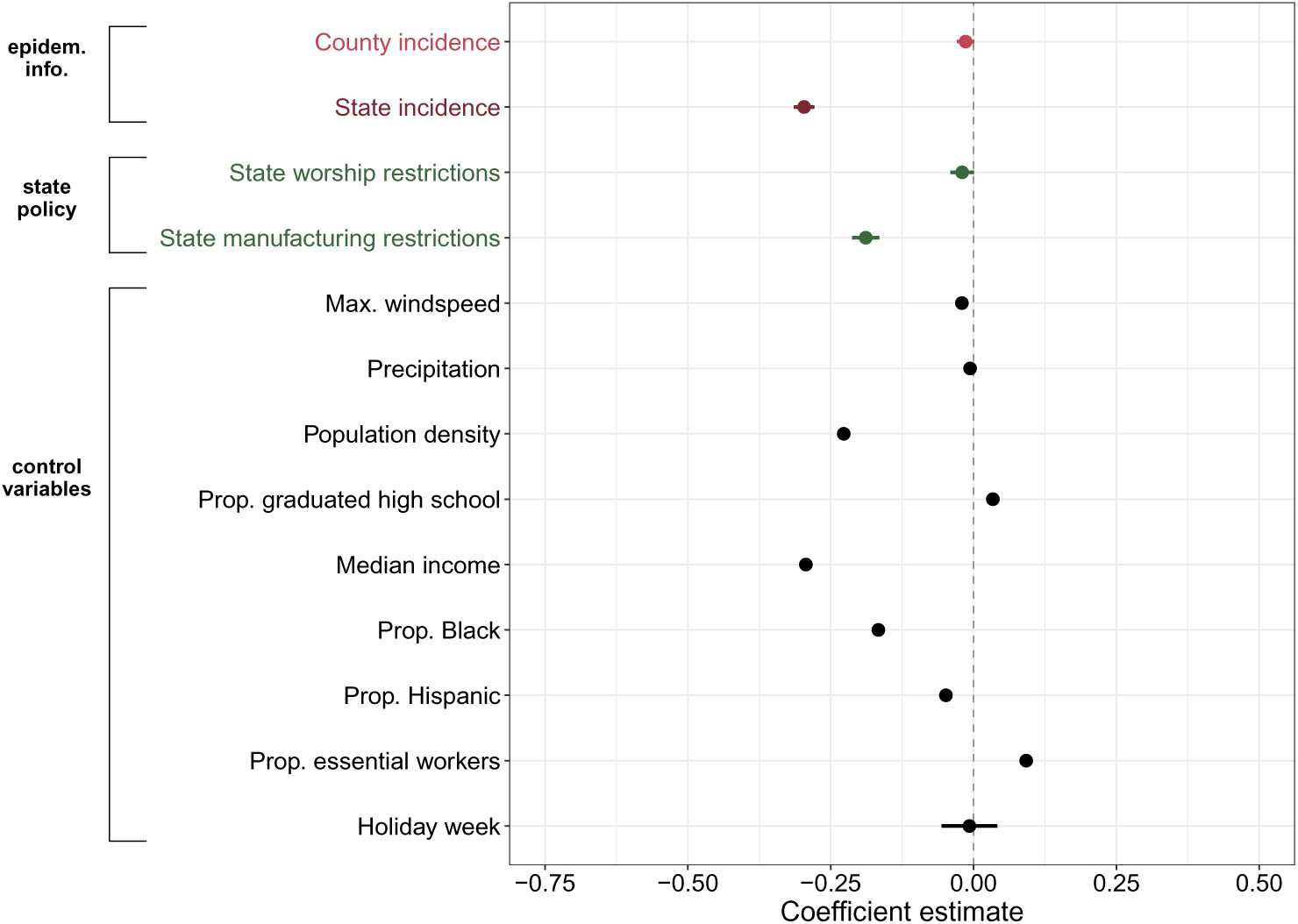
Main regression model using VIF threshold of 5 and excluding social information variables. State case incidence data remains more predictive than county under stronger multicollinearity constraints.

**Figure S22:**
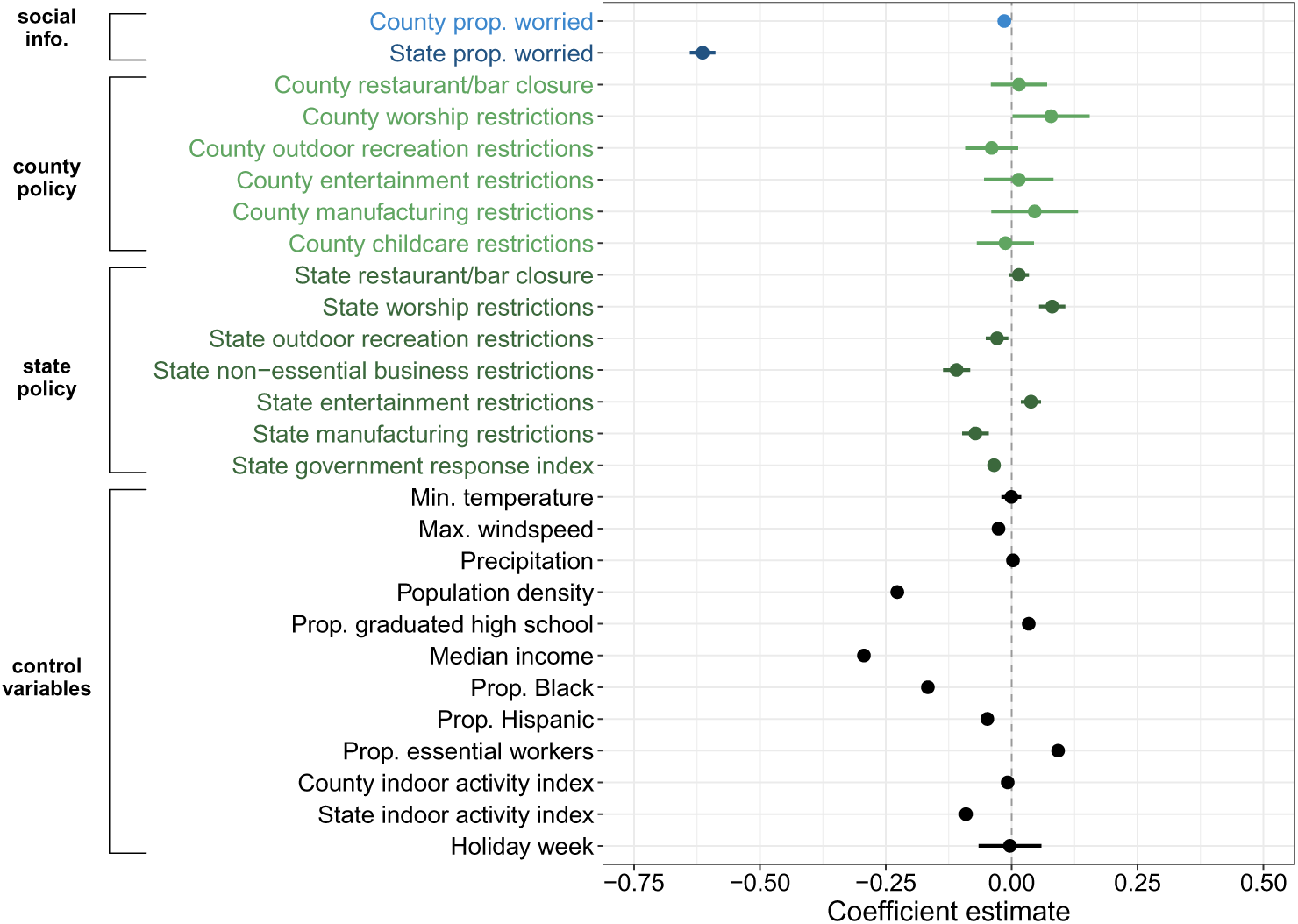
Main regression model using VIF threshold of 5 and excluding epidemiological information variables. State proportion worried data remains more predictive than county under stronger multicollinearity constraints.

**Figure S23:**
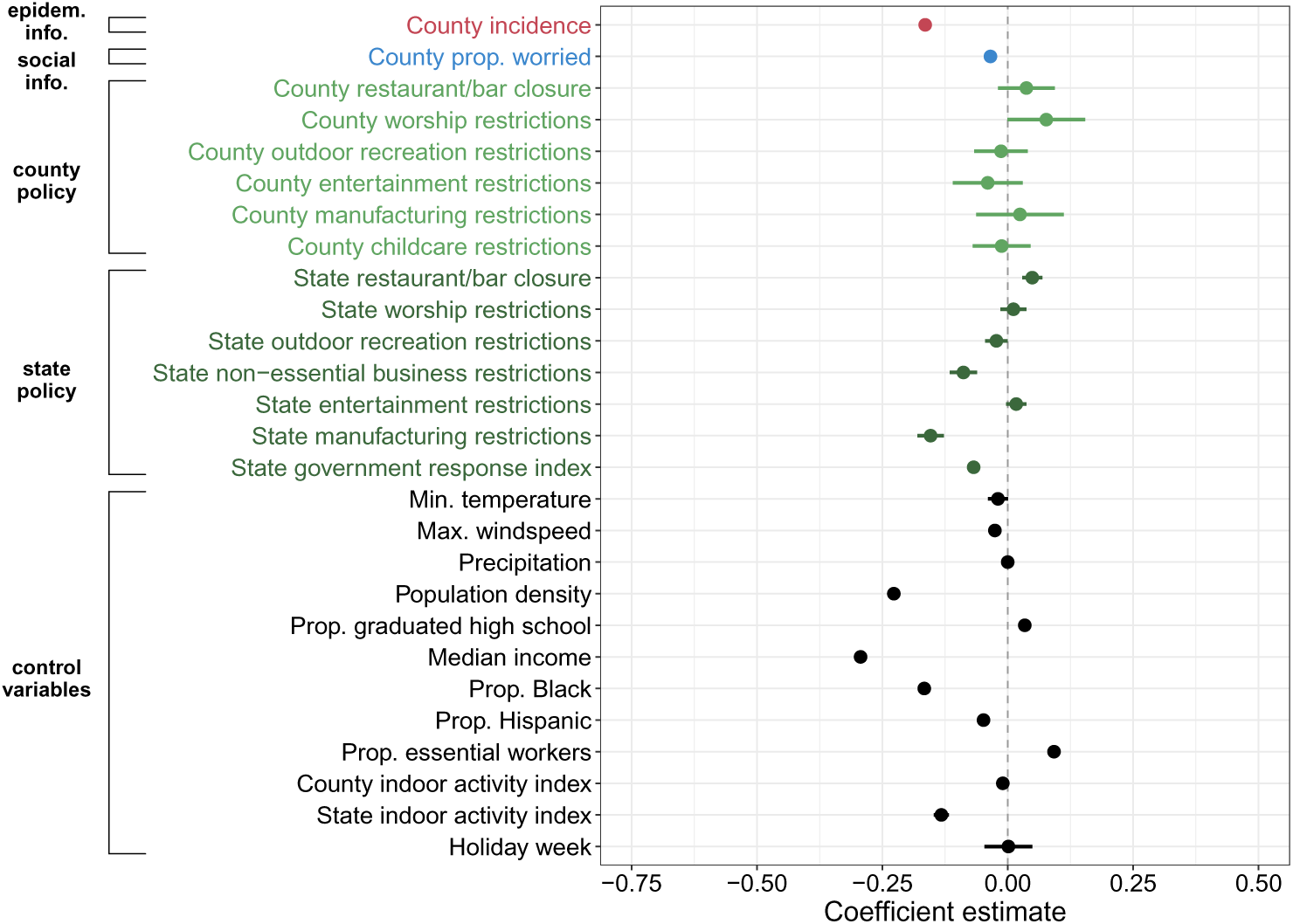
Main regression model using VIF threshold of 5 and excluding state-level epidemiological and social information variables. At the county scale, incidence is more predictive of behavior than proportion worried.

**Figure S24:**
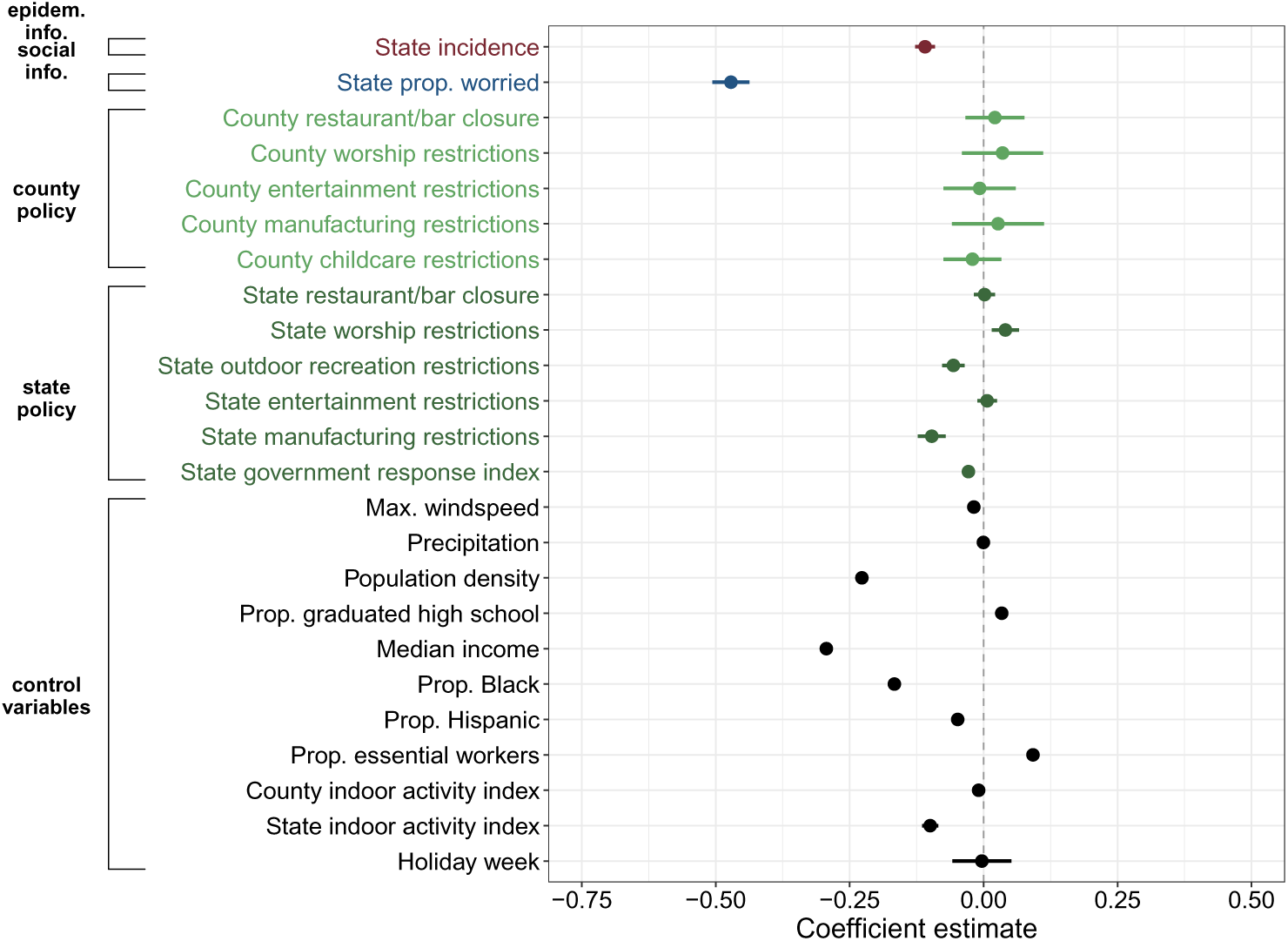
Main regression model using VIF threshold of 5 and excluding county-level epidemiological and social information variables. State social information remains more predictive than state epidemiological information under stronger multicollinearity constraints.

**Figure S25:**
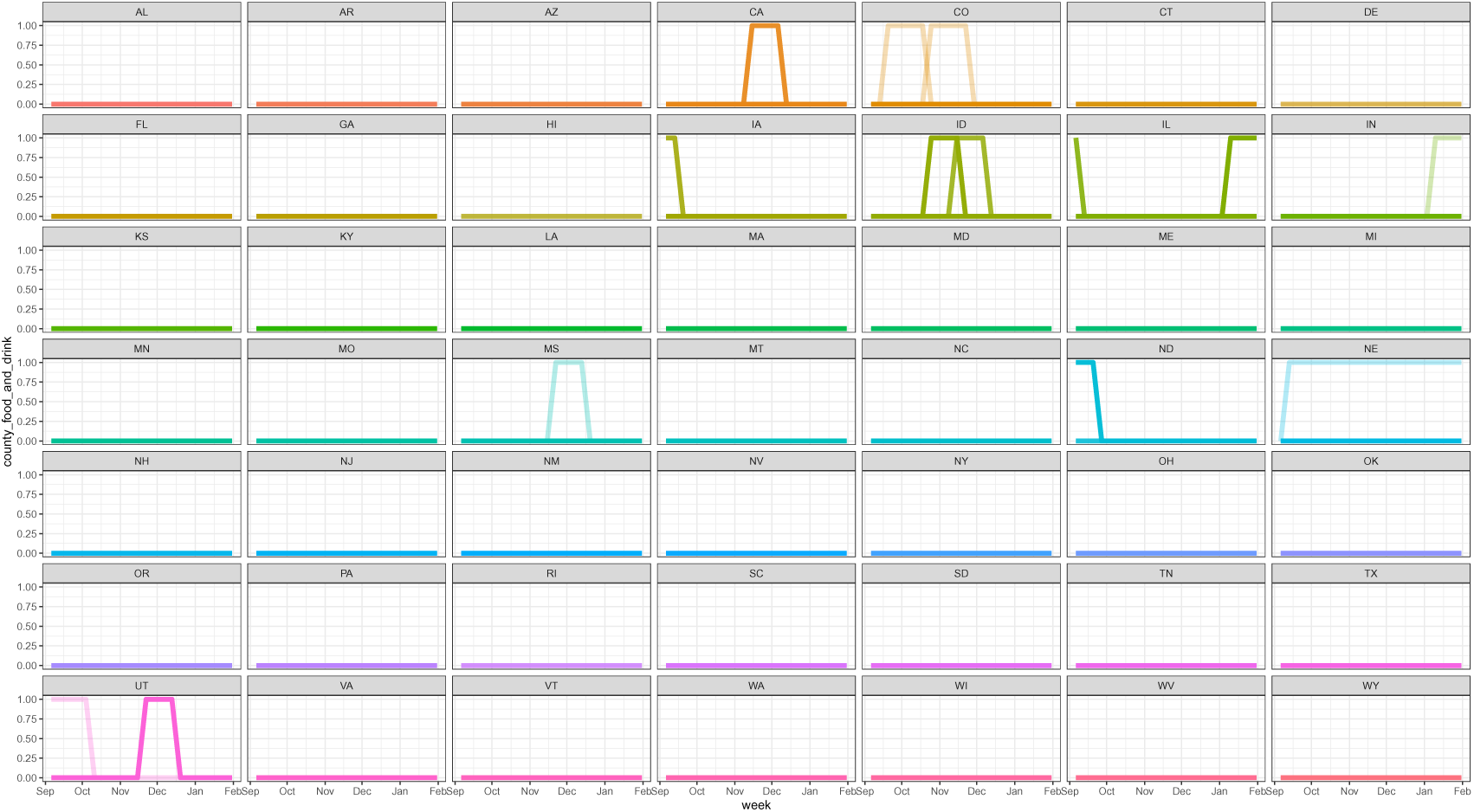
County restaurant/bar closure data. Each line represents a county and each color a state. Values of 1 indicate a policy in place, while 0 indicates there was no policy in place.

**Figure S26:**
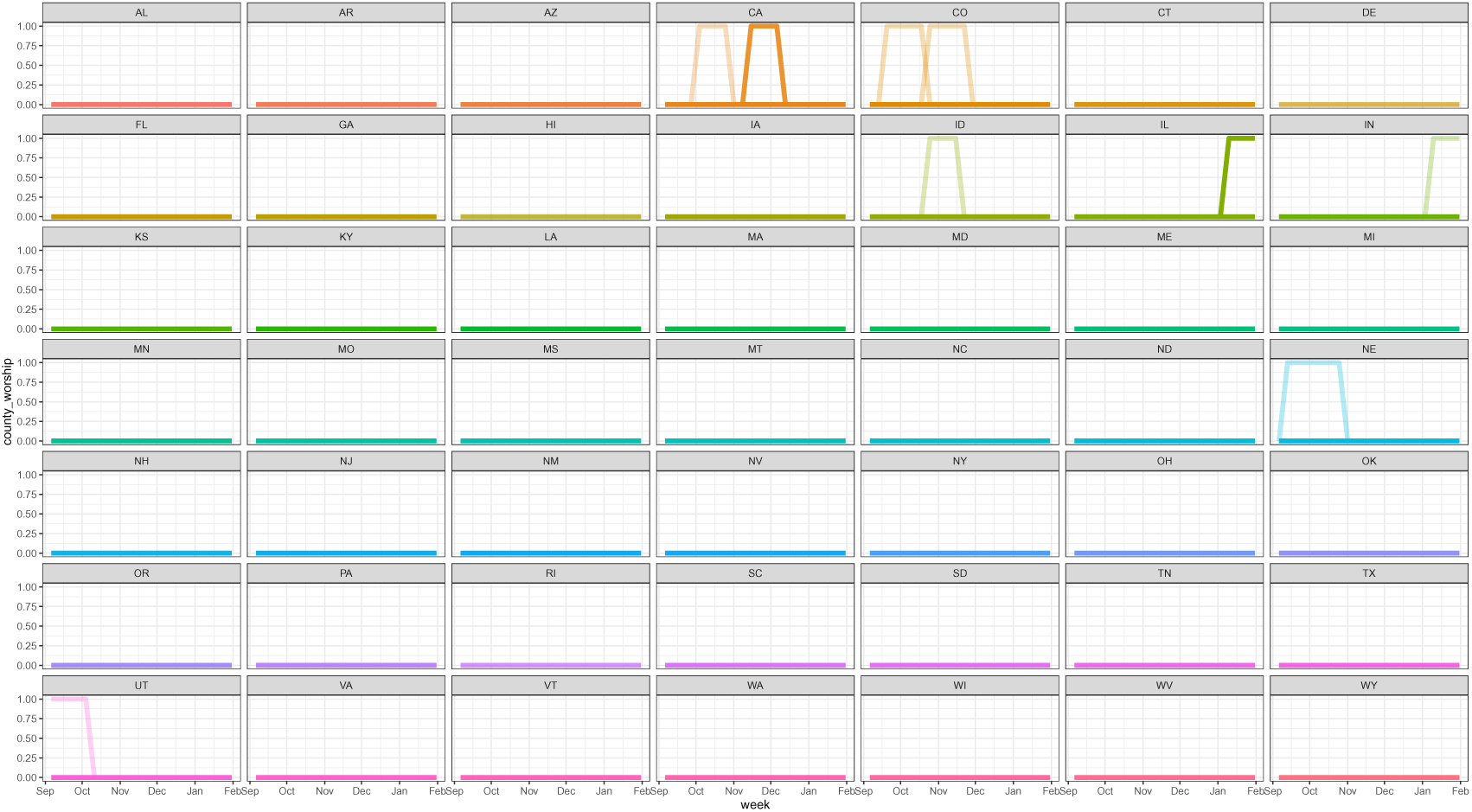
County worship restrictions data. Each line represents a county and each color a state. Values of 1 indicate a policy in place, while 0 indicates there was no policy in place.

**Figure S27:**
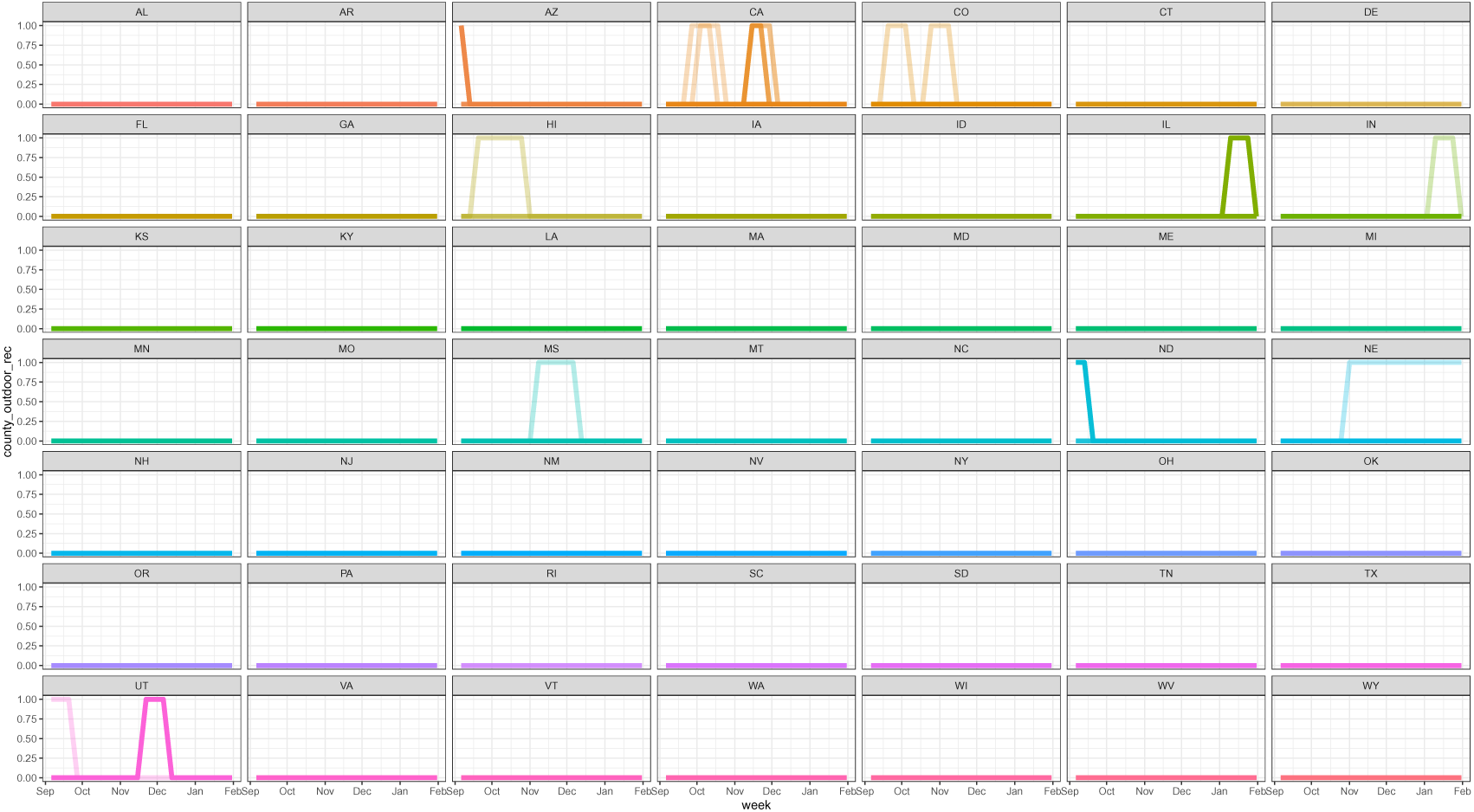
County outdoor recreation restrictions data. Each line represents a county and each color a state. Values of 1 indicate a policy in place, while 0 indicates there was no policy in place.

**Figure S28:**
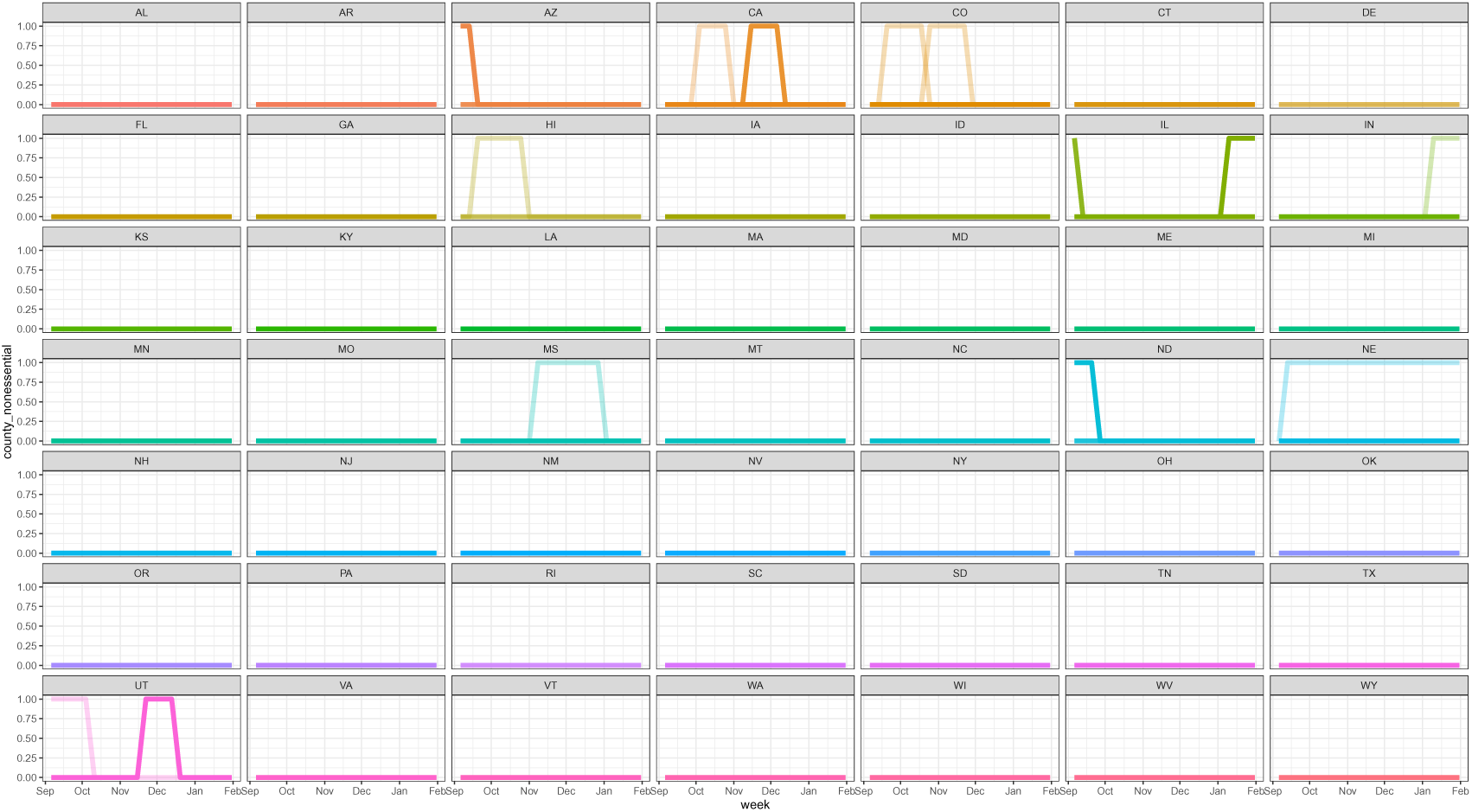
County nonessential business restrictions data. Each line represents a county and each color a state. Values of 1 indicate a policy in place, while 0 indicates there was no policy in place.

**Figure S29:**
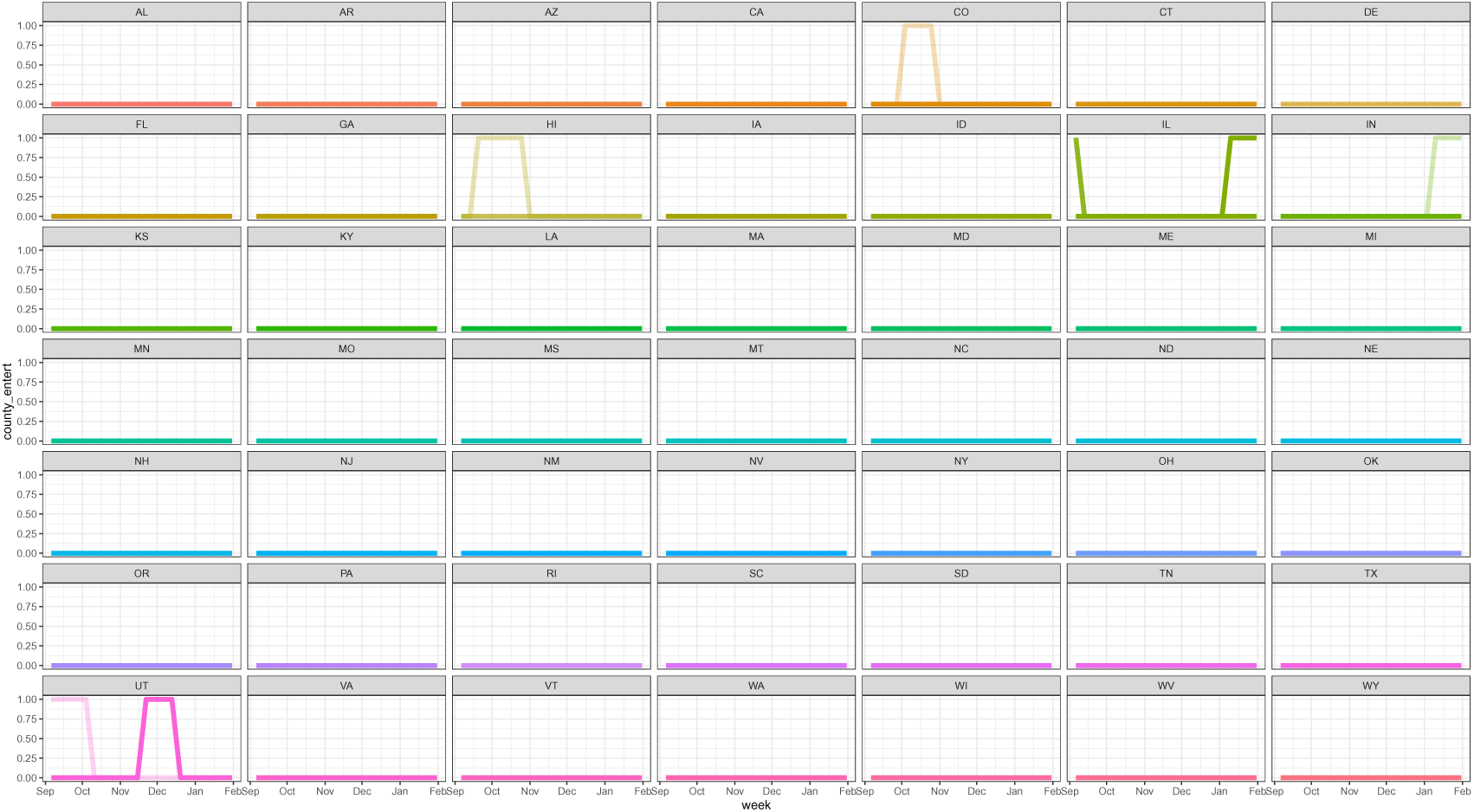
County entertainment restrictions data. Each line represents a county and each color a state. Values of 1 indicate a policy in place, while 0 indicates there was no policy in place.

**Figure S30:**
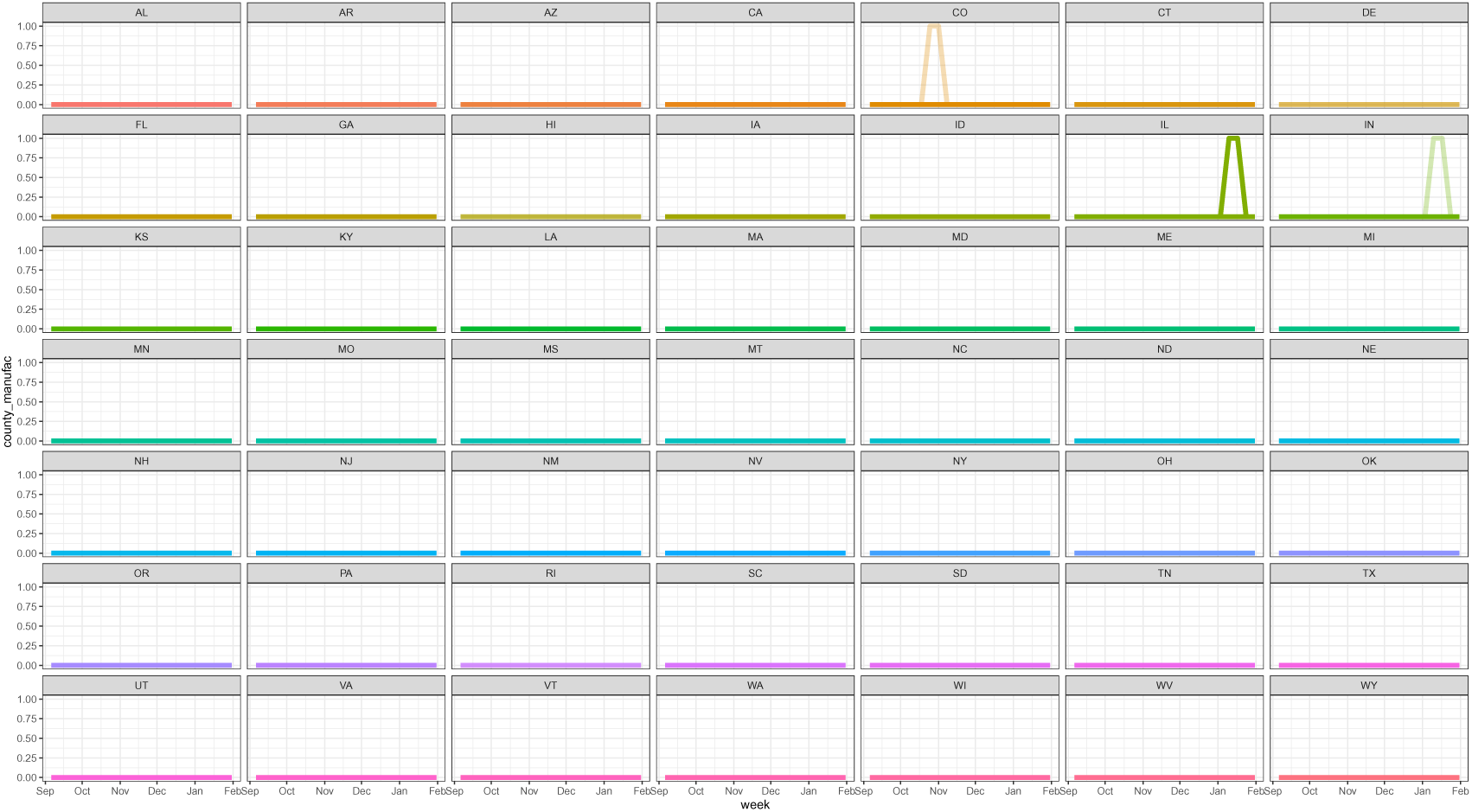
County manufacturing restrictions data. Each line represents a county and each color a state. Values of 1 indicate a policy in place, while 0 indicates there was no policy in place.

**Figure S31:**
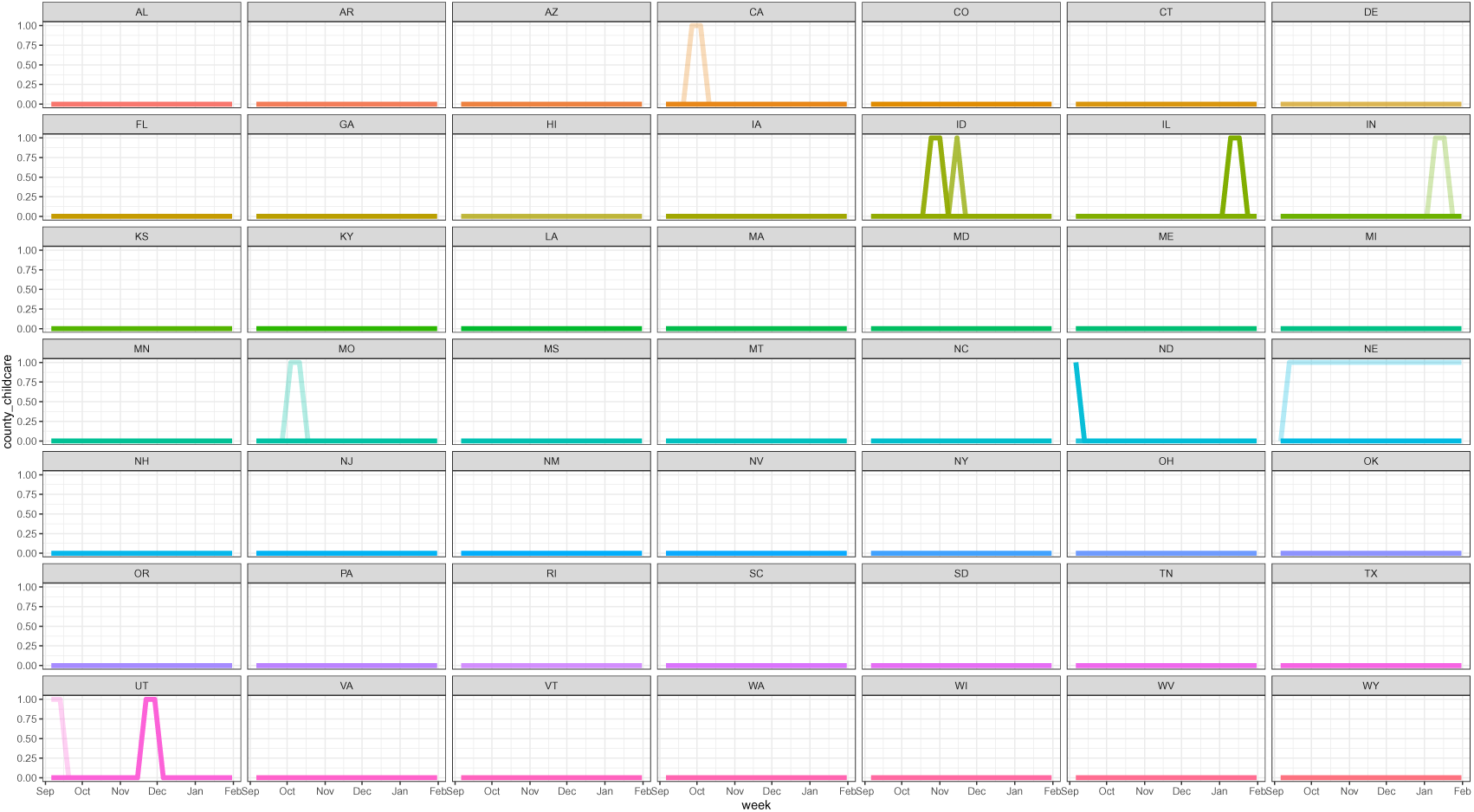
County childcare restrictions data. Each line represents a county and each color a state. Values of 1 indicate a policy in place, while 0 indicates there was no policy in place.

**Figure S32:**
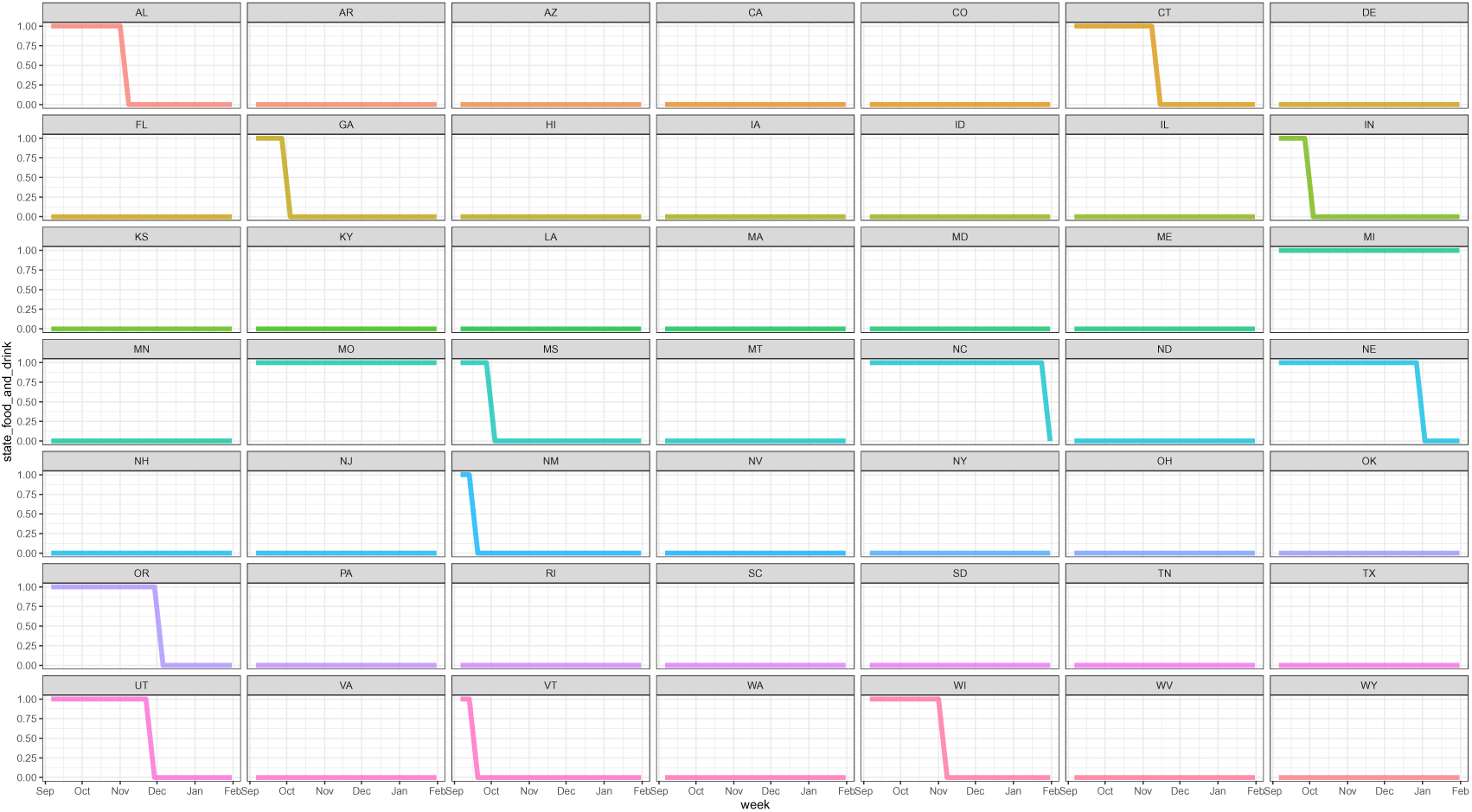
State restaurant/bar closure data. Each line and color represents a state. Values of 1 indicate a policy in place, while 0 indicates there was no policy in place.

**Figure S33:**
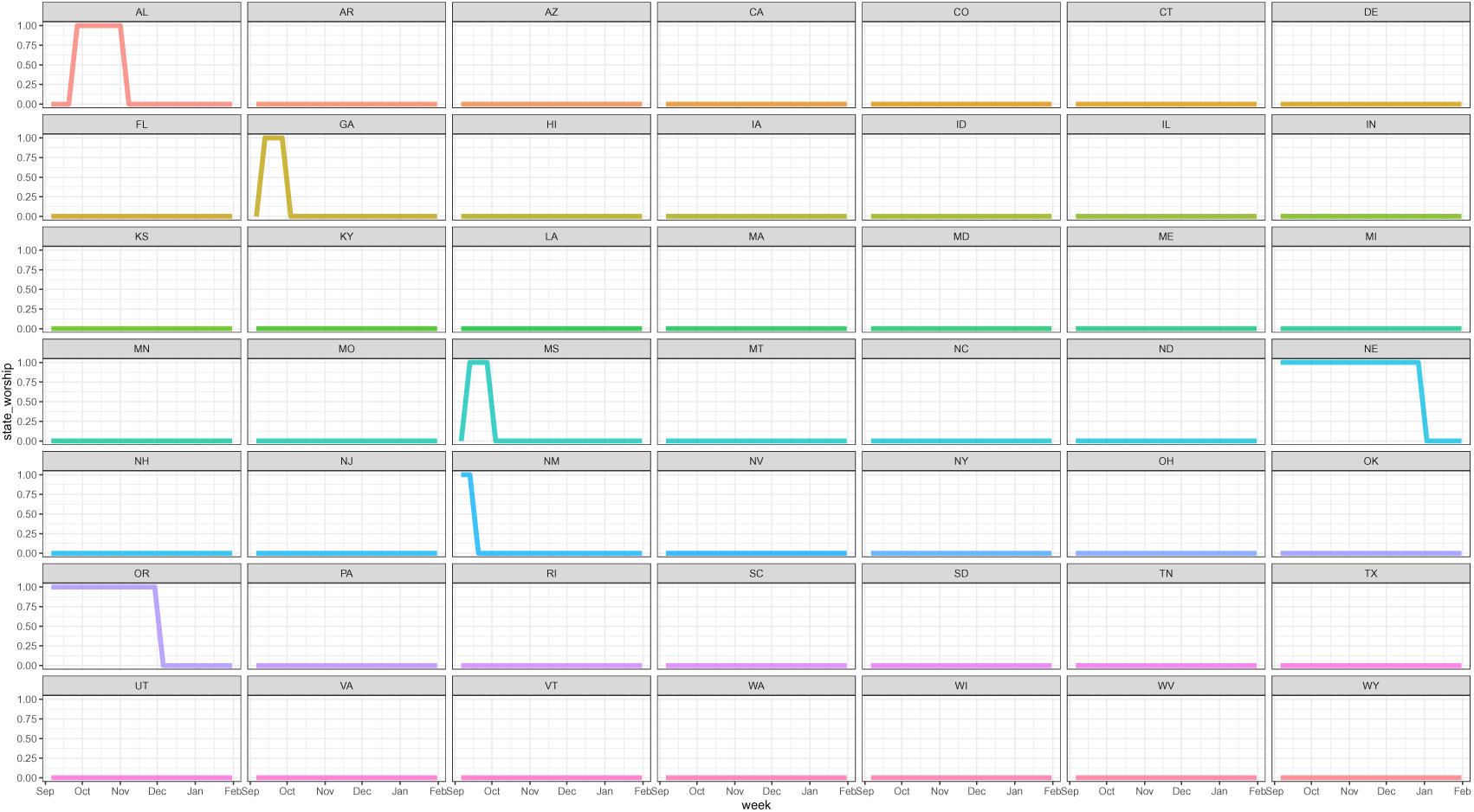
State worship restrictions data. Each line and color represents a state. Values of 1 indicate a policy in place, while 0 indicates there was no policy in place.

**Figure S34:**
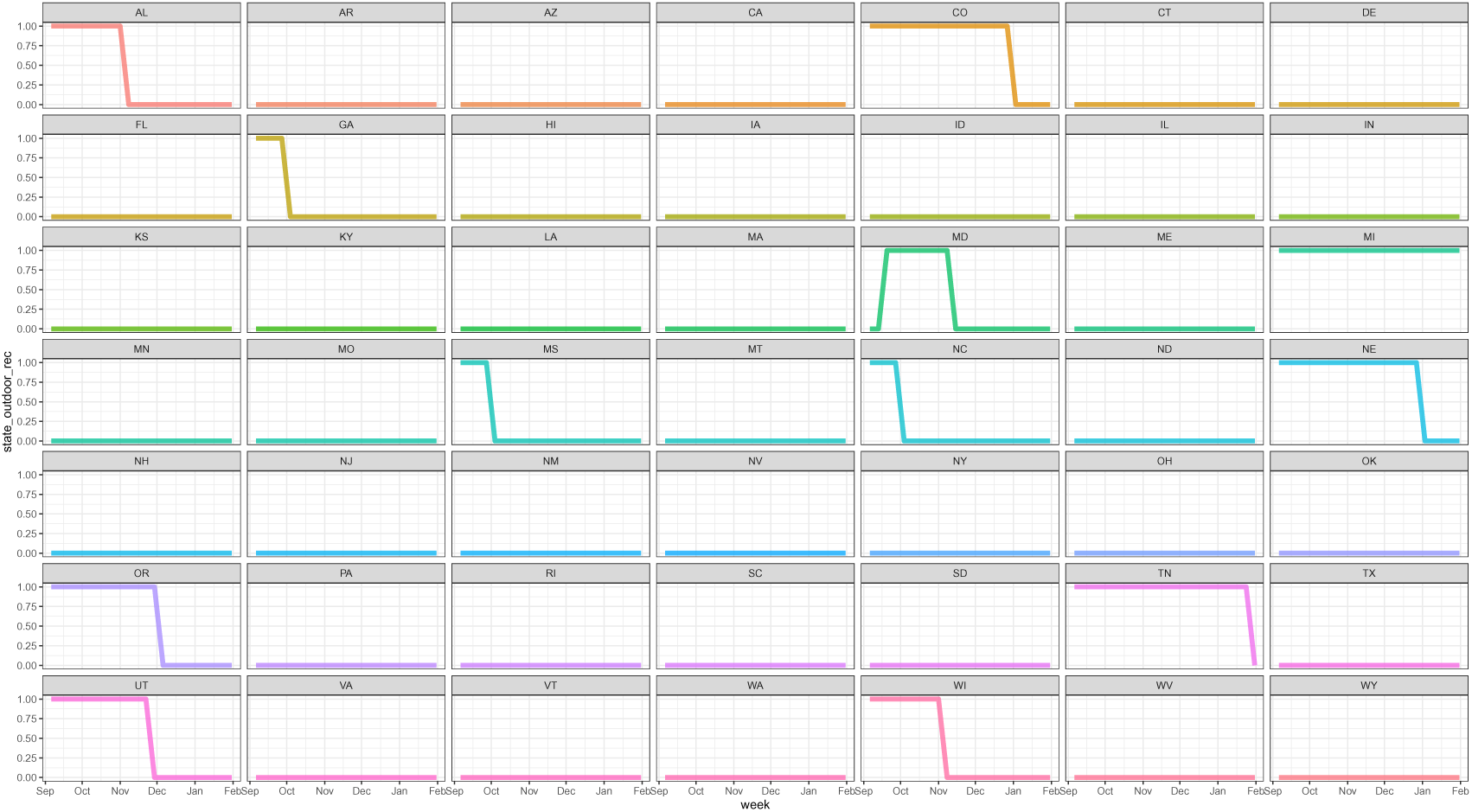
State outdoor recreation restrictions data. Each line and color represents a state. Values of 1 indicate a policy in place, while 0 indicates there was no policy in place.

**Figure S35:**
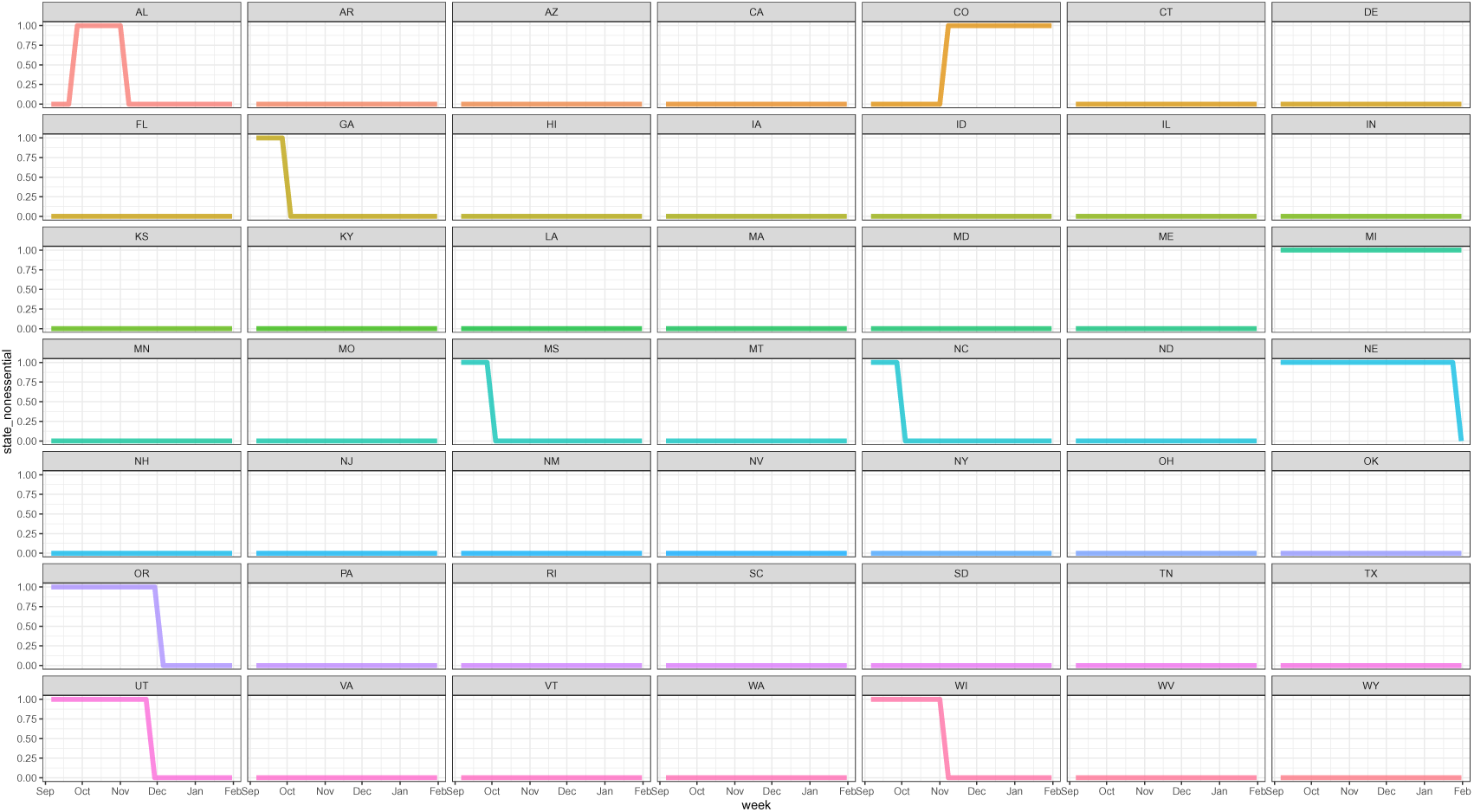
State nonessential business restrictions data. Each line and color represents a state. Values of 1 indicate a policy in place, while 0 indicates there was no policy in place.

**Figure S36:**
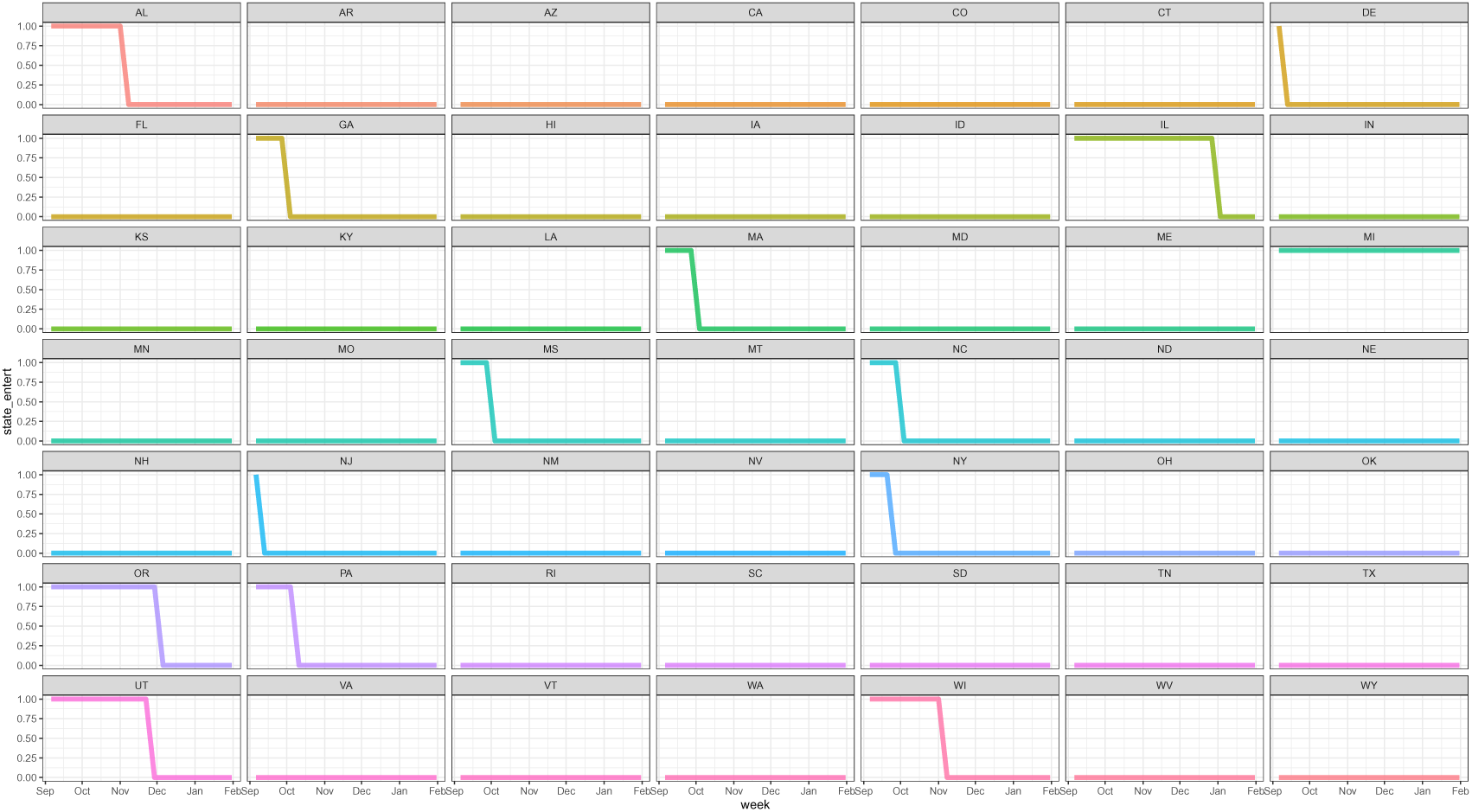
State entertainment restrictions data. Each line and color represents a state. Values of 1 indicate a policy in place, while 0 indicates there was no policy in place.

**Figure S37:**
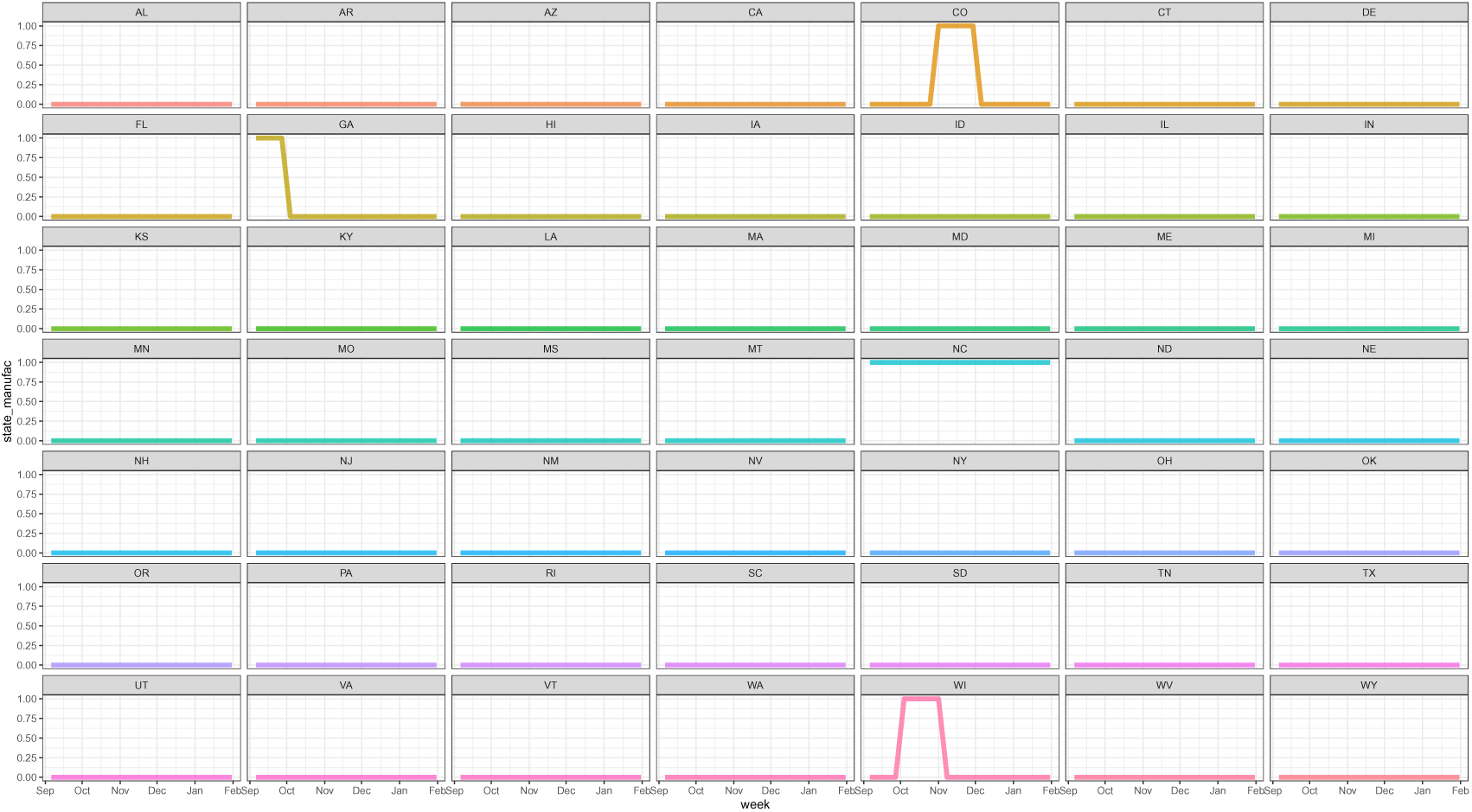
State manufacturing restrictions data. Each line and color represents a state. Values of 1 indicate a policy in place, while 0 indicates there was no policy in place.

**Figure S38:**
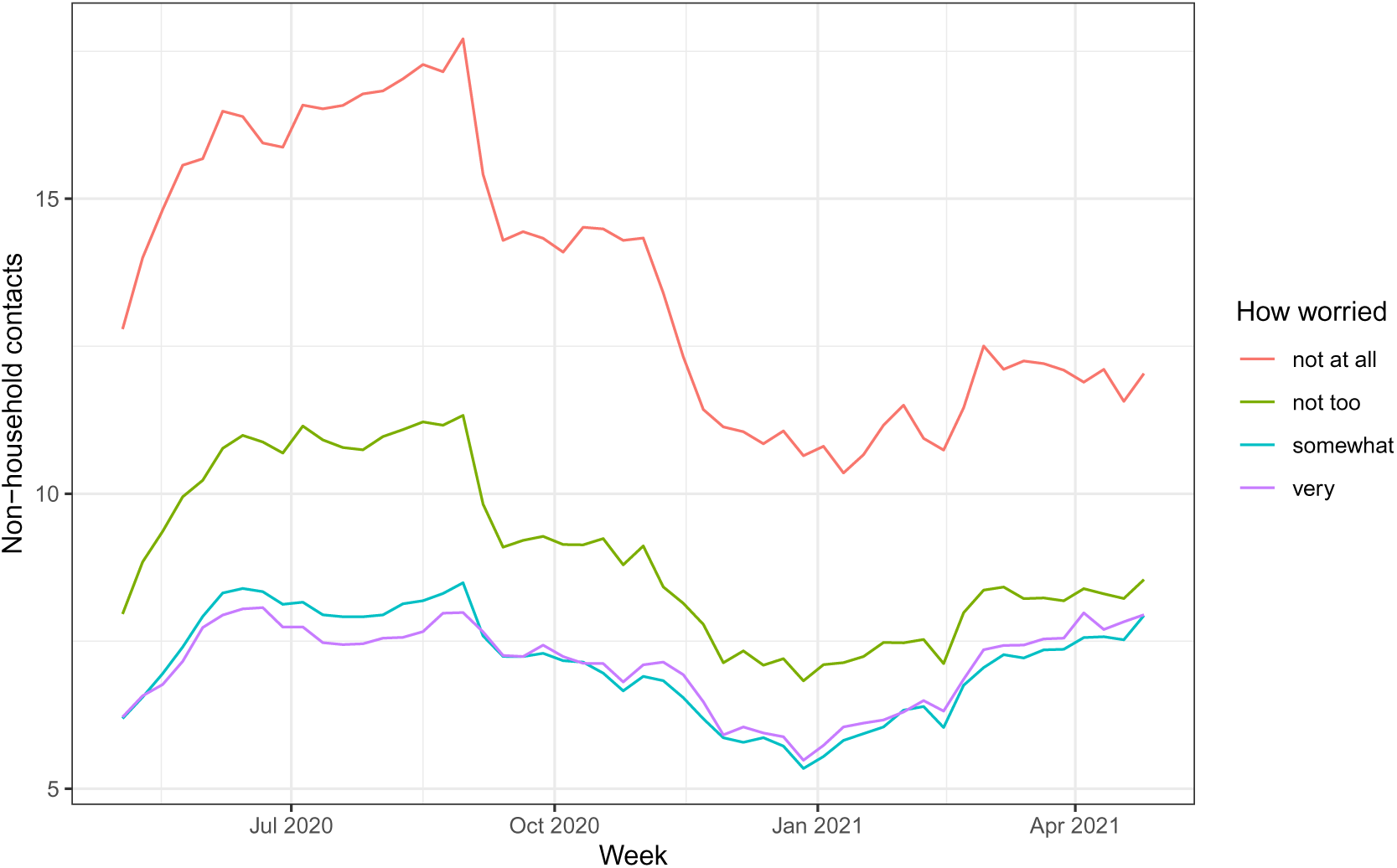
Weekly mean of daily non-household contacts by worry status. We classify both very and somewhat worried people as “worried” as they have similar contact rates over our study period.

**Figure S39:**
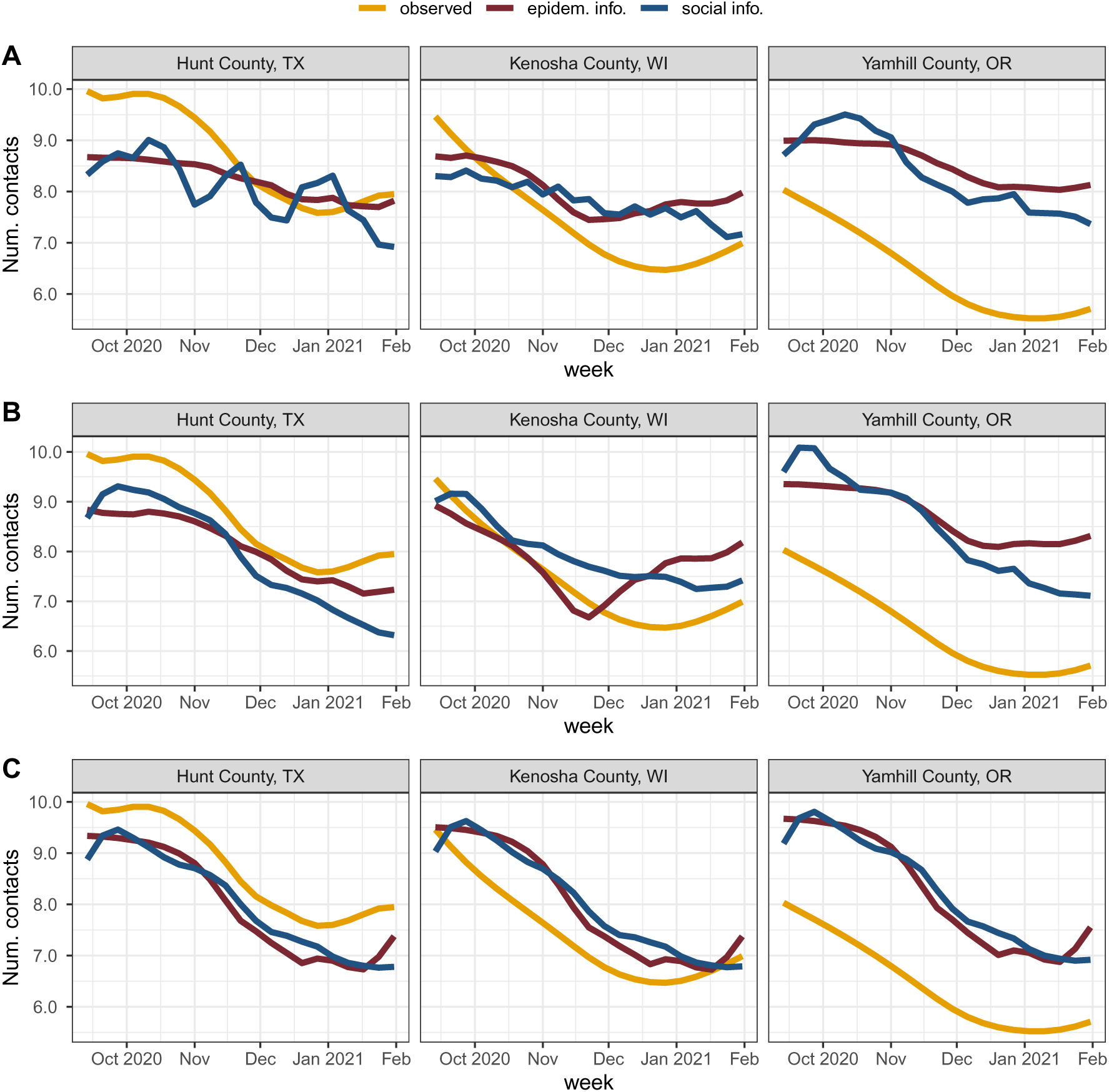
Contact predictions for three case study counties shown in main text using predictors at different spatial scales. (A) county predictors, (B) state predictors, (C) national predictors. Contact predictions from epidemiological and social information tend to be similar, with more divergence at smaller spatial scales in these case studies.

**Figure S40:**
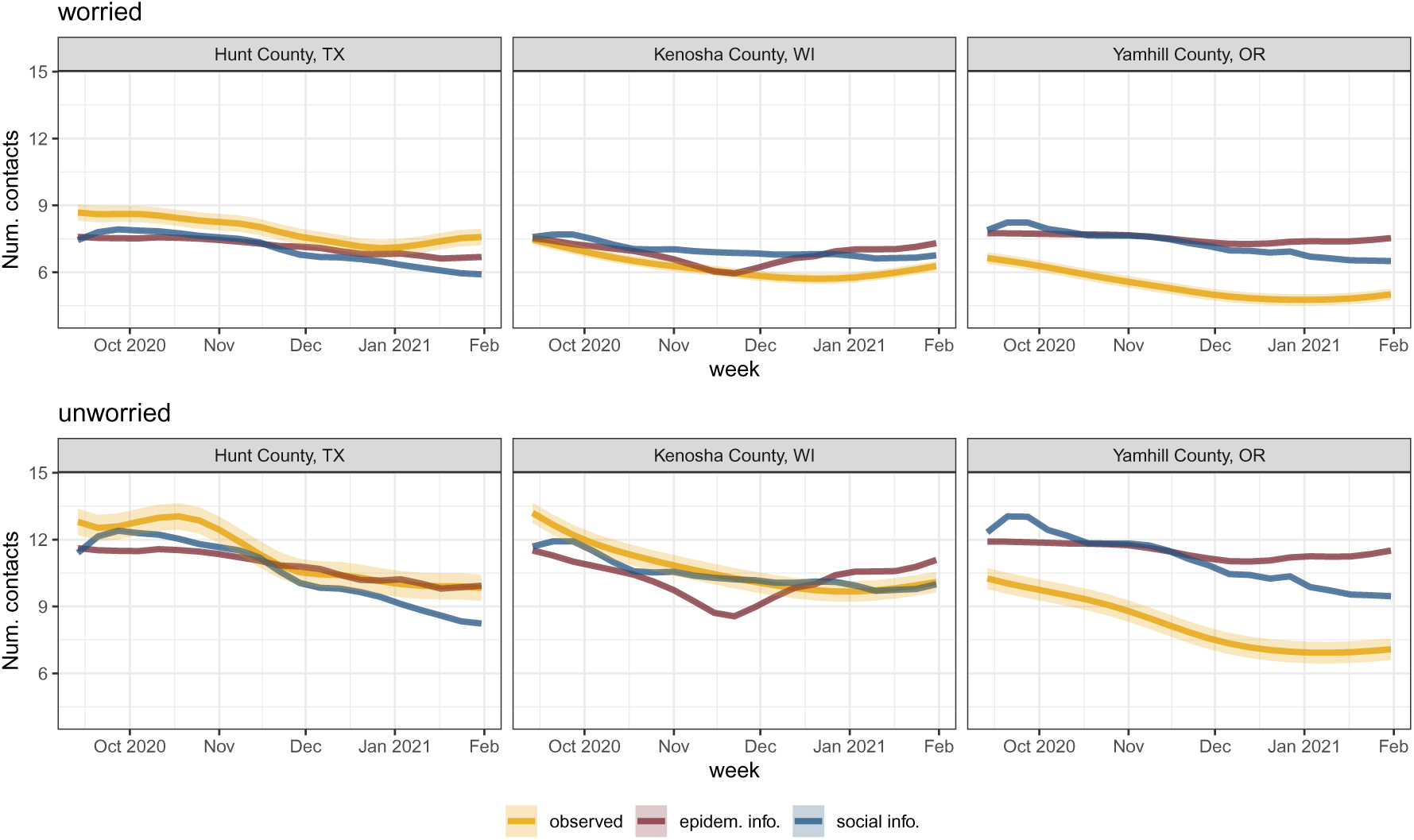
Contact predictions for three case study counties shown in the main text disaggregated by worry status and with uncertainty (one standard error). Errors for contact estimates from epidemiological and social information are very small.

**Figure S41:**
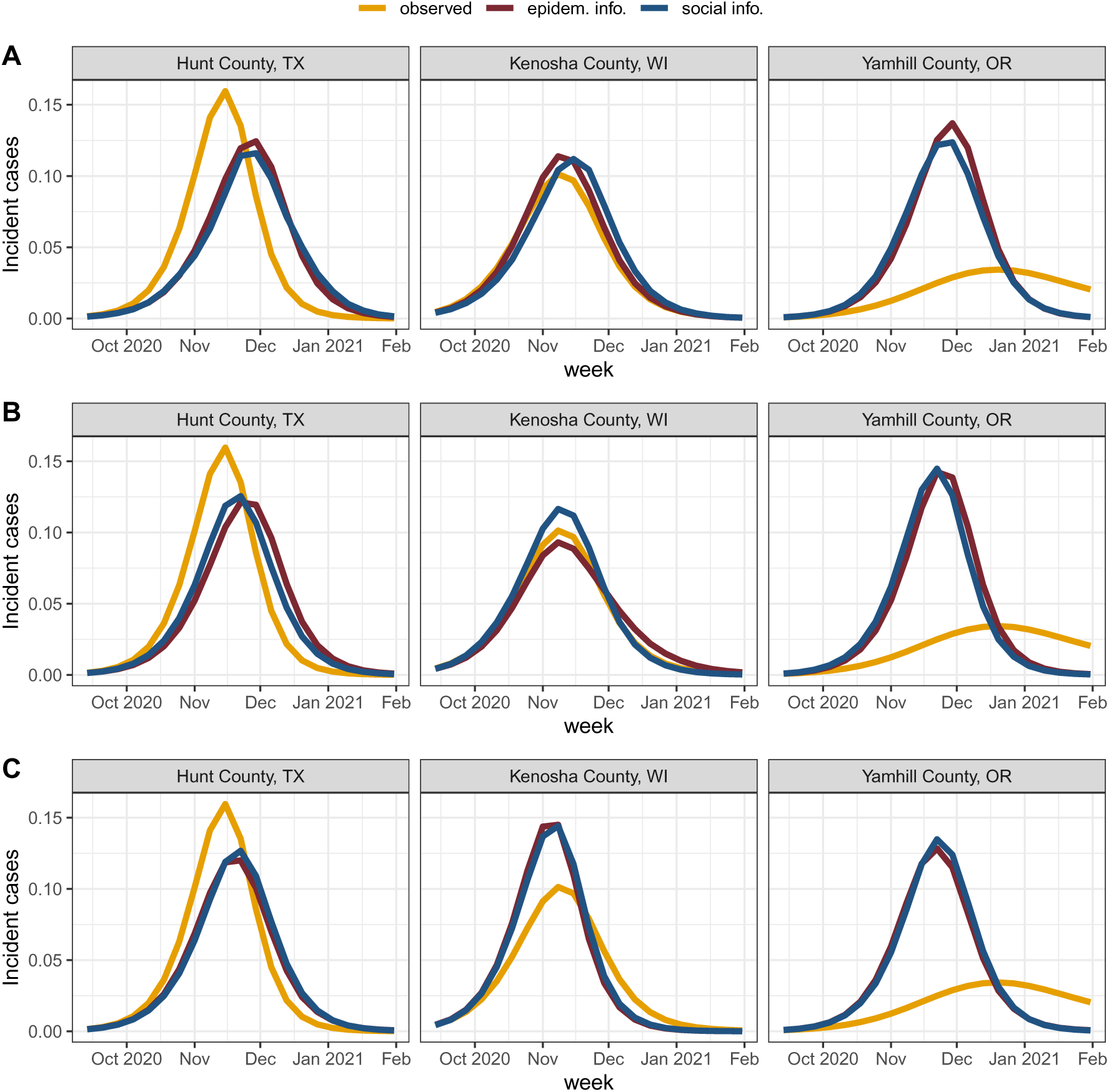
Disease predictions for three case study counties shown in main text using contact predicted based on epidemiological and social information at different spatial scales. (A) county predictors, (B) state predictors, (C) national predictors. Disease predictions from epidemiological and social information-based contact predictions tend to be similar.

**Figure S42:**
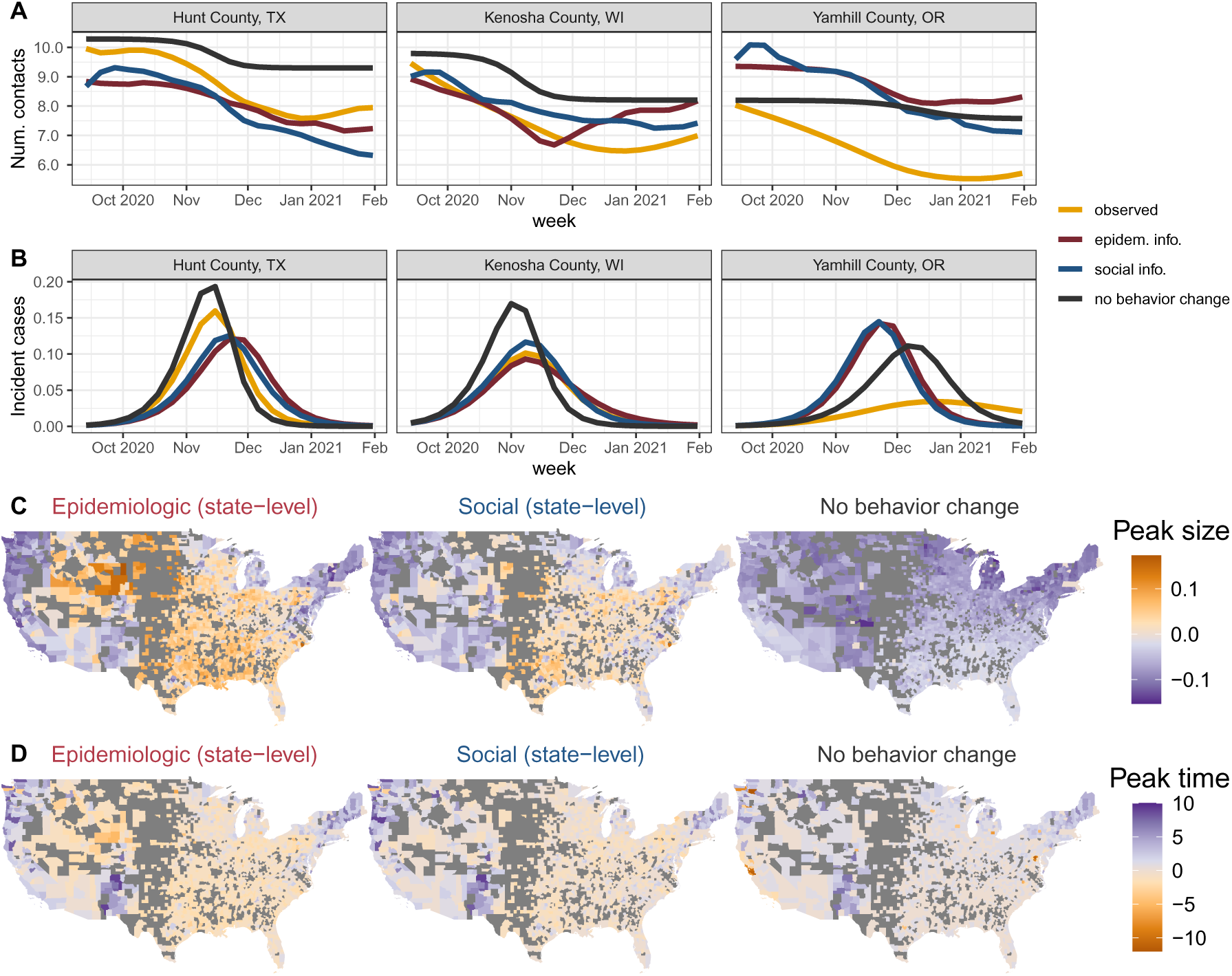
Behavior change reduces epidemic peak size and delays epidemic peak time. (A) Three case study counties demonstrating the range in predicted social distancing using case incidence (objective) versus proportion worried (subjective) at the state level compared to the observed social distancing for winter 2020-21. The no behavior change scenario fixes all individuals at the contact rate observed for their worry group (worried or unworried) in the week of September 6, 2020. Contact still changes over time in this simulation as high degree individuals are preferentially depleted. Higher contact rates correspond to less social distancing. All three counties are considered metropolitan areas in terms of population density. (B) The repercussions of social distancing predictions for disease projections in the same three case study counties. (C, D) Epidemic repercussions across all counties for social distancing predictions based on state incidence, proportion worried, or no behavior change. Peak size is the maximum proportion of the population infected at any one time (C) and peak time is the week in which the peak size occurs (D). For peak size, positive values (orange) indicate that predicted epidemics had a smaller peak incidence than observed, while negative values (purple) denote that predicted epidemics had a larger peak incidence than observed. For peak time, positive values (purple) denote predicted epidemics peaked earlier than observed and negative values (orange) indicate that predicted epidemics peaked later than observed.

**Figure S43:**
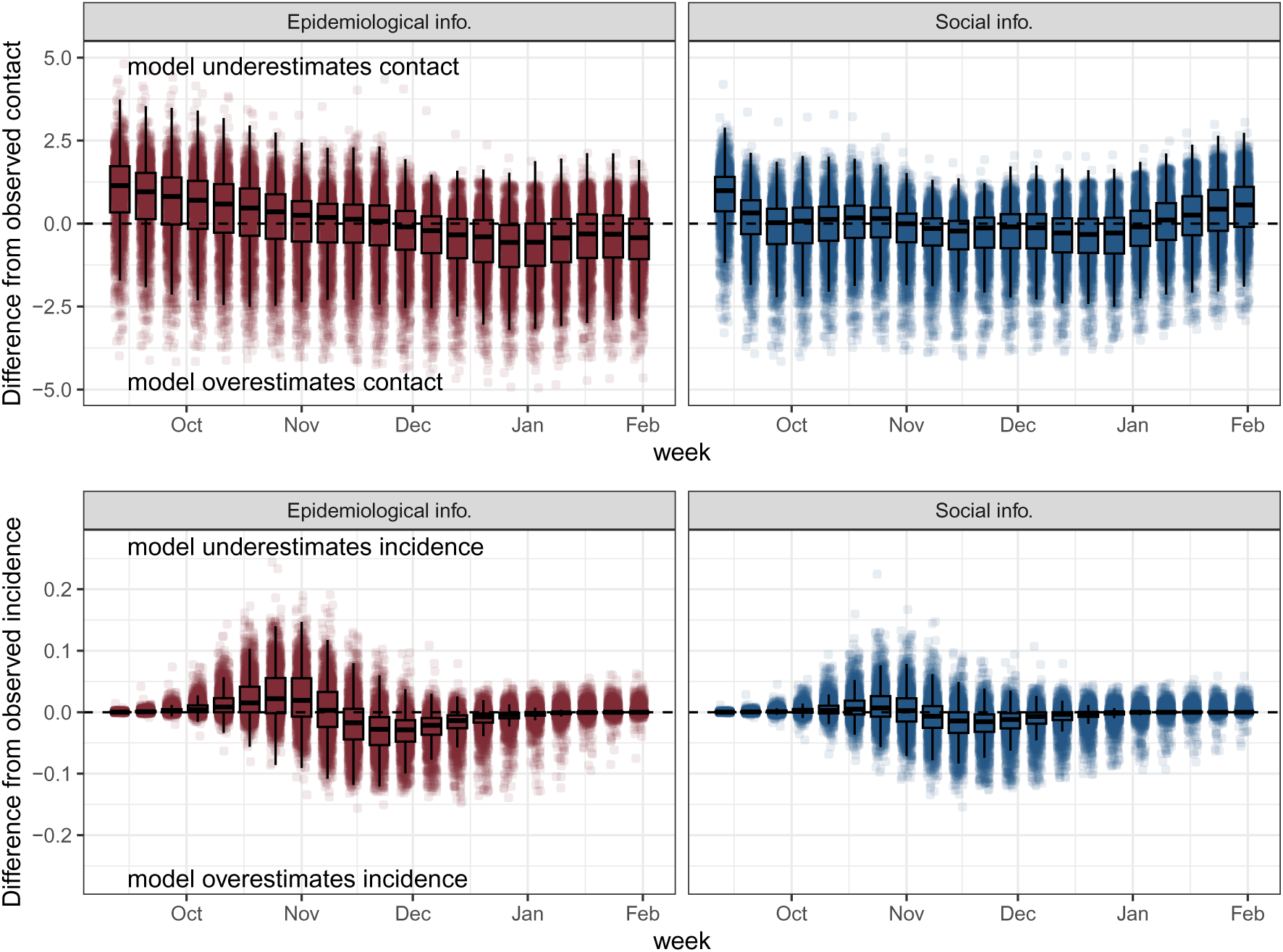
Contact and incidence residuals when using predicted contact based on epidemiological or social information from September 2020 through January 2021. Across counties, epidemiological information-based behavior change underestimates contact rates in late fall 2020 leading to under-estimates of disease incidence, while social information-based estimates are consistently close to observed contact and predicted disease incidence across the study period.

**Figure S44:**
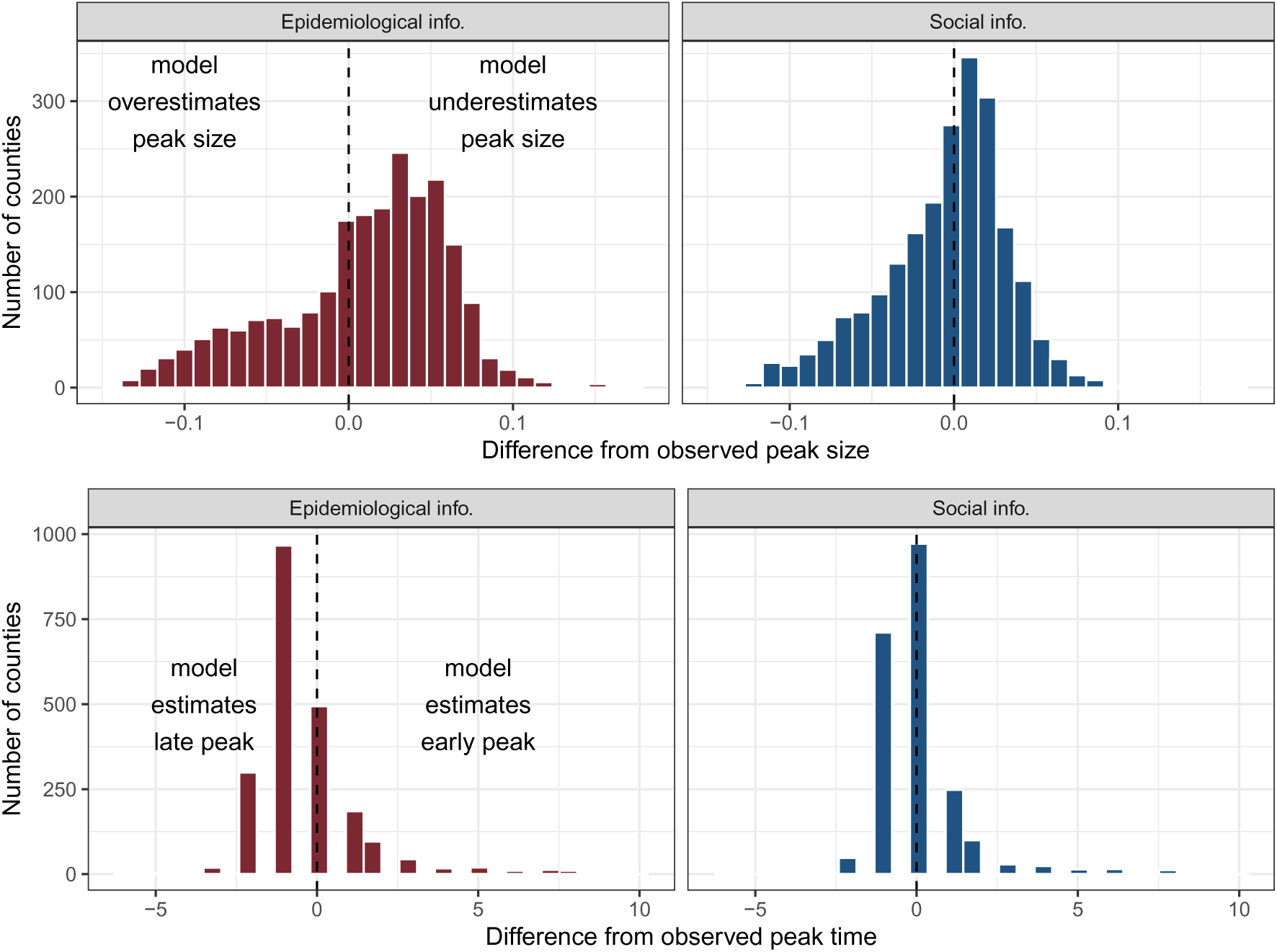
Peak time and size residuals when using predicted contact based on epidemiological or social information. Epidemiological information-based models are more likely to underestimate peak size and estimate a later peak than is observed; social information-based models better predict contact rates and disease incidence.

